# A framework for reconstructing SARS-CoV-2 transmission dynamics using excess mortality data

**DOI:** 10.1101/2021.10.04.21264540

**Authors:** Mahan Ghafari, Oliver J Watson, Ariel Karlinsky, Luca Ferretti, Aris Katzourakis

## Abstract

Detailed reconstruction of the SARS-CoV-2 transmission dynamics and assessment of its burden in several parts of the world has still remained largely unknown due to the scarcity of epidemiological analyses and limited testing capacities of different countries to identify cases and deaths attributable to COVID-19 [1-4]. Understanding the true burden of the Iranian COVID-19 epidemic is subject to similar challenges with limited clinical and epidemiological studies at the subnational level [5-9]. To address this, we develop a new quantitative framework that enables us to fully reconstruct the transmission dynamics across the country and assess the level of under-reporting in infections and deaths using province-level, age-stratified all-cause mortality data. We show that excess mortality aligns with seroprevalence estimates in each province and subsequently estimate that as of 2021-10-22, only 48% (95% confidence interval: 43-55%) of COVID-19 deaths in Iran have been reported. We find that in the most affected provinces such as East Azerbaijan, Qazvin, and Qom approximately 0.4% of the population have died of COVID-19 so far. We also find significant heterogeneity in the estimated attack rates across the country with 11 provinces reaching close to or higher than 100% attack rates. Despite a relatively young age structure in Iran, our analysis reveals that the infection fatality rate in most provinces is comparable to high-income countries with a larger percentage of older adults, suggesting that limited access to medical services, coupled with undercounting of COVID-19-related deaths, can have a significant impact on accurate estimation of COVID-19 fatalities. Our estimation of high attack rates in provinces with largely unmitigated epidemics whereby, on average, between 10% to 25% individuals have been infected with COVID-19 at least twice over the course of 20 months also suggests that, despite several waves of infection, herd immunity through natural infection has not been achieved in the population.

---

Assessing the true burden of COVID-19 requires a comprehensive surveillance system to monitor outbreaks and a large capacity for diagnostic testing. However, this is particularly challenging for many low- and middle-income countries due to limited diagnostic capacities and access to healthcare [10, 1]. An alternative approach to measuring the burden of COVID-19 is to monitor changes in all-cause mortality trends with respect to baseline levels based on historic trends (i.e., measuring excess mortality). While excess mortality during the current pandemic may not exactly match the true number of COVID-19-related deaths due to various reasons such as disruptions in treatment for other fatal diseases and socio-economic disparities in health care in different regions of a country, several works have shown that a significant portion of excess deaths in many countries are directly attributable to COVID-19 [1, 5, 12-15]. This enables us to use excess mortality as a proxy for estimating the number of infections and fatalities from COVID-19 and correct for under-reporting of cases and deaths that may arise due to limited testing from suspected cases and uncertainty in the number of fatalities attributable to COVID-19 [16, 7].

Iran was among the first countries outside mainland China to report a large outbreak of SARS-CoV-2 in February and March 2020 and also acted as a major source for its spread in several countries in the Middle East and elsewhere [18]. It was also one of the first countries to experience a second wave of infection after the relaxation of non-pharmaceutical interventions (NPIs) in July 2020 [6, 9]. Since then, the country experienced three additional waves, two of which were driven by the Alpha and Delta variants of SARS-CoV-2, in May and September 2021. Despite a recent increase in the number of daily administered vaccine doses, Iran had a very slow start on its immunisation programme with only ∼3% of the population being fully vaccinated by the end of July 2021, during the surge in cases with the Delta variant [20]. Because the Ministry of Health and Medical Education (MoHME) of Iran stopped releasing province-level data on the number of confirmed COVID-19 cases and deaths from 2020-03-22, the transmission dynamics across the country have remained largely unknown. One way to estimate the number of infections and account for under-reporting of cases is by converting deaths attributable to COVID-19 to infections using a population-weighted infection fatality ratio (IFR) estimate (i.e., infections = deaths / IFR), taking into account the demographics of the population of interest [21]. However, such estimation of attack rates can be inaccurate or misleading if it does not take into account the impact of limited access to health care in municipalities with low socioeconomic status, uneven adoption of pharmaceutical and non-pharmaceutical intervention in different areas, and the increased chance of re-infection over time. Neglecting to account for these factors can result in an overestimate of SARS-CoV-2 attack rates and underestimation its potential for continued transmission, with immediate implications for assessments of the potential burden of SARS-CoV-2 variants of concern such as Omicron in a population. In this work, we use the newly updated province-level age-stratified weekly all-cause mortality data from the National Organization for Civil Registration (NOCR) of Iran and develop a mathematical framework to fully reconstruct the Iranian epidemic with the aim to estimate the attack rate, number of daily hospital admissions, deaths, and re-infection rates of SARS-CoV-2 across the country.

## Results

We obtain age-stratified weekly all-cause mortality data from NOCR and apply a statistical model to calculate the weekly excess mortality for all 31 provinces and across 17 age-groups during the Iranian epidemic (see Methods section). **Figure 1A** shows 5 distinct peaks in excess mortality with a temporal trend that is strongly associated with the nationwide reported COVID-19 deaths over time. Several provinces show significant levels of excess mortality from the first week of February 2020 suggesting that the Iranian epidemic likely started at least a month before that (**Figure 1B**). This is in agreement with previous phylogenetic and epidemiological studies suggesting the epidemic started in late December 2019 to early January 2020 [6, 22]. Furthermore, during the first two waves of the Iranian epidemic, the excess mortality was roughly 2.5 times higher than reported COVID-19 fatalities. However, this dropped to 2.1 at the later stages suggesting that the testing capacity of the country to record infections and deaths gradually improved over time, but not to the extent to fully capture the majority of COVID-19-related deaths. This is also in agreement with statements from the Iranian health officials suggesting that Iran’s COVID-19-related deaths could be twice the official numbers and also previous studies showing elevated levels of under-reporting of COVID-19 fatalities in Iran, particularly in the early stages of the pandemic [6, 7]. We also find that the cumulative nationwide excess deaths by 2021-10-22 is 257,950 (95% CI: 221,530 - 278,440) which is roughly twice the 125,000 reported COVID-19 deaths at the time.

**Figure 1:**
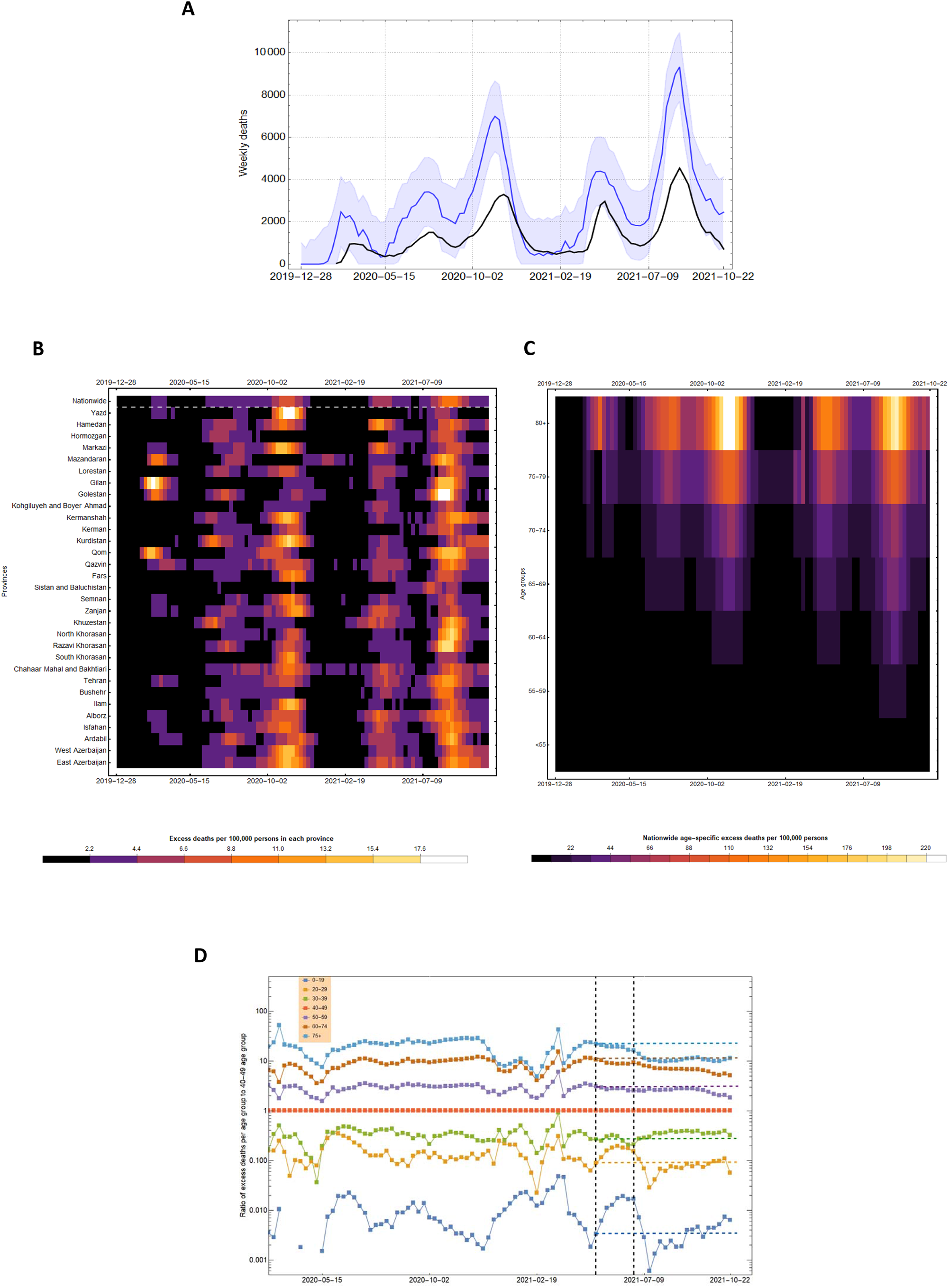
**(A)** One-month average of weekly excess mortality (blue) and reported COVID-19 deaths (black) in Iran from 2019-12-28 to 2021-10-22. Shaded area shows the 95% forecast interval in weekly excess mortality. **(B)** One-month average of weekly excess deaths per 100,000 persons per province over time. **(C)** Nationwide excess deaths per 100,000 persons per age group over time. **(D)** Ratio of weekly excess per age-group to the 40-49 age-group. Vertical dashed lines show the start of the immunisation programme for individuals aged above 75 on 2021-05-18 and above 60 on 2021-07-14. The horizontal dashed lines show the ratio of excess deaths at the start of the immunisation programme for individuals aged above 75.

**Figure 1B** shows the intensity of excess mortality as quantified by the number of excess deaths per 100,000 persons in each province over time. During the first wave in spring 2020, only a few provinces showed significant levels of excess mortality with Gilan, Qom, Mazandaran, and Golestan among the hardest-hit provinces. This is likely because these provinces usually attract a large number of tourists and pilgrims around the New Year’s holiday in February/March which led to large superspreading events and uncontrolled outbreaks due to limited NPIs before the national lockdown on 2020-03-05 [6]. As the epidemic progressed, the timing of the peak excess mortality between different provinces were more simultaneous (**Figure 1B**; **Figure S1**). Most notably, during the third wave in October/November 2020 and the fifth wave in August/September 2021 with the Delta variant of SARS-CoV-2, almost all provinces experienced significant number of fatalities around the same time.

We also carry out a similar analysis to estimate the temporal pattern of COVID-19 fatalities for different age-groups (**Figure 1C**). We find that the majority of excess deaths are concentrated in the older age categories (>55) and that the third and fifth epidemic waves had the largest impact on younger age groups. **Figure 1D** shows the per capita ratio of excess mortality per age group with respect to the 40–49 age group. While we see a gradual increase in the ratio of deaths in the older age-groups during the first three waves, there is a two-fold drop in the 60-74 and 75+ age-groups before the start of the fifth wave. This is likely due to a higher vaccination coverage in these age-groups. While the exact number of vaccinated individuals per age-group in Iran is not known, given that the immunisation programme for individuals aged >75 started in May 2021 (first phase priority groups for vaccination in Iran are frontline healthcare workers as well as those aged above 80; see [20] for more information), and that Iran fully vaccinated roughly 5 times the population size of individuals aged 75 and above by 2021-09-07, it is likely that the drop in excess mortality in the older age-groups before and during the fifth wave is due to their high vaccination coverage.

Assuming that lack of testing is the primary reason behind the discrepancy between excess mortality and official COVID-19 deaths [1, 23, 24], we attribute the mean excess mortality during the pandemic period to COVID-19. This enables us to reconstruct the SARS-CoV-2 transmission dynamics across the country and infer the population fatality rate (PFR), IFR, and attack rate per province (see Methods section). We first estimate the national and provincial attack rates of SARS-CoV-2 under two different set of assumptions of the IFR by age relationship and select the better-fitting model by comparing the modelled seroprevalence estimates to data (**Figure S2; Data S1-S6**). We find that the model with a higher IFR per age-group (and an overall lower estimated attack rate) that explicitly accounts for an increase in IFR when hospital capacity is reached (see Methods section) provides a better fit to seroprevalence data. With a median value of approximately 0.50% (0.32-0.64%), our inferred province-level IFR is roughly twice as high as the population-weighted IFR, which is calculated using age-specific IFR estimates derived from serological studies in predominantly high-income countries (**Table 1**) [21]. This discrepancy is expected because disparities in healthcare access can be offset by the younger population age structure of many developing countries resulting in population IFRs that are similar to high-income countries [12, 25, 26]. **Figure 2A** and **Figure 2B** show a nearly 75% attack rate of SARS-CoV-2 across the country with nearly 15% of all infections due to reinfections by 2021-10-22. We also find that the age-distribution of the modelled COVID-19 deaths are well-aligned with the pattern of excess mortality per age-group and both follow a clear log-linear relationship with respect to age, a signature characteristic of COVID-19 fatalities [27], suggesting that the model successfully reconstructs age-dependent contact patterns for disease spread in Iran (**Figure 2C**). There is also a strong association between the modelled and observed daily hospital admissions over time and across provinces, further indicating that the majority of excess deaths during the pandemic period are directly attributable to COVID-19 (**Figure S3**). There is, however, a notably lower number of modelled daily hospital admissions compared to observed hospitalisations in some provinces (e.g., South Khorasan, Lorestan, and Yazd) during the last wave in comparison to earlier waves. To explore these patterns, we use a mixed-effects model to examine the relationship between the number of hospitalisations inferred from our model fits and the observed confirmed COVID-19 hospital admissions over time and across epidemic waves while controlling for province-specific variation (see Methods section). Using this framework, we show that there was no significant decrease in the ratio of model inferred hospitalisations to observed hospitalisations during the last wave compared to previous waves, and that the majority of variation in hospitalisation patterns observed is explained by province-level random effects, most likely due to specific changes in treatment seeking or the age demographics being most affected in later waves (**Figure S4**). However, a significant decrease in the ratio of model inferred hospitalisations to observed hospitalisations was observed since the start of the pandemic, which suggests that case fatality rates from hospitalised infections have decreased during this period, likely due to a combination of improved case management, increased number of infections resulting from reinfections, and higher vaccination coverage.

**Table 1:** The overall excess mortality, PFR, estimated IFR (estimated from the transmission model), population-weighted IFR (based on age distribution of each province), and percentage of the population ever exposed to SARS-CoV-2 in each province as of 2021-10-22.

| Province | Excess deaths | % PFR | % Estimated IFR | % Population - weighted IFR* | % Exposed |
| --- | --- | --- | --- | --- | --- |
| East Azerbaijan | 17,035 (8,887 - 28,237) | 0.438 (0.228 - 0.726) | 0.50 (0.41 - 0.62) | 0.30 (0.28 - 0.32) | 84.92 (69.06 - 103.05) |
| West Azerbaijan | 11,609 (6,289 - 19,204) | 0.346 (0.187 - 0.573) | 0.47 (0.36 - 0.57) | 0.24 (0.22 - 0.25) | 75.66 (60.25 - 98.53) |
| Ardabil | 4,538 (1,974 - 8,053) | 0.361 (0.157 - 0.641) | 0.38 (0.31 - 0.46) | 0.26 (0.25 - 0.28) | 92.55 (76.19 - 112.14) |
| Isfahan | 18,281 (9,612 - 30,412) | 0.355 (0.187 - 0.591) | 0.52 (0.41 - 0.64) | 0.29 (0.27 - 0.31) | 65.90 (51.80 - 81.46) |
| Alborz | 11,068 (5,477 - 18,219) | 0.388 (0.192 - 0.639) | 0.49 (0.38 - 0.61) | 0.24 (0.22 - 0.25) | 64.09 (51.07 - 81.97) |
| Ilam | 1,719 (611 - 3,743) | 0.294 (0.104 - 0.640) | 0.47 (0.38 - 0.56) | 0.23 (0.22 - 0.25) | 61.35 (51.01 - 73.74) |
| Bushehr | 3,023 (1,241 - 5,649) | 0.244 (0.100 - 0.45) | 0.43 (0.31 - 0.55) | 0.18 (0.17 - 0.19) | 61.29 (44.63 - 82.10) |
| Tehran | 49,320 (27,155 - 78,900) | 0.365 (0.201 - 0.585) | 0.47 (0.39 - 0.58) | 0.28 (0.26 - 0.30) | 77.06 (65.08 - 93.99) |
| Chahaar Mahal & Bakhtiari** | 3,624 (1,089 - 7,452) | 0.376 (0.113 - 0.774) | 0.48 (0.37 - 0.59) | 0.24 (0.22 - 0.26) | 76.28 (63.01 - 101.11) |
| South Khorasan | 1,736 (473 - 4,466) | 0.210 (0.059 - 0.558) | 0.50 (0.43 - 0.59) | 0.26 (0.24 - 0.29) | 41.27 (34.65 - 48.50) |
| Razavi Khorasan | 20,847 (9,945 - 36,946) | 0.300 (0.147 - 0.549) | 0.36 (0.29 - 0.43) | 0.23 (0.22 - 0.25) | 90.19 (71.76 - 113.32) |
| North Khorasan | 2,478 (985 - 5,250) | 0.283 (0.112 - 0.600) | 0.36 (0.27 - 0.44) | 0.22 (0.21 - 0.24) | 77.73 (61.94 - 100.78) |
| Khuzestan | 15,366 (8,235 - 25,189) | 0.317 (0.170 - 0.520) | 0.32 (0.26 - 0.40) | 0.19 (0.18 - 0.20) | 98.48 (79.82 - 120.59) |
| Zanjan | 3,938 (1,442 - 7,797) | 0.367 (0.134 - 0.720) | 0.50 (0.42 - 0.61) | 0.28 (0.26 - 0.30) | 73.11 (60.85 - 87.84) |
| Semnan | 2,057 (742 - 4,226) | 0.275 (0.099 - 0.566) | 0.49 (0.43 - 0.58) | 0.27 (0.25 - 0.29) | 57.26 (49.85 - 65.92) |
| Sistan & Baluchistan | 4,401 (1,104 - 12,829) | 0.140 (0.036 - 0.419) | 0.37 (0.25 - 0.49) | 0.13 (0.12 - 0.13) | 46.60 (35.38 - 67.60) |
| Fars | 13,825 (6,361 - 25,441) | 0.282 (0.130 - 0.520) | 0.50 (0.40 - 0.62) | 0.26 (0.24 - 0.28) | 56.63 (42.64 - 71.82) |
| Qazvin | 5,255 (1,793 - 9,699) | 0.404 (0.138 - 0.747) | 0.47 (0.38 - 0.59) | 0.25 (0.23 - 0.27) | 85.97 (69.52 - 106.44) |
| Qom | 5,519 (3,193 - 8,925) | 0.399 (0.231 - 0.646) | 0.49 (0.33 - 0.62) | 0.21 (0.19 - 0.22) | 84.74 (66.12 - 116.36) |
| Kurdistan | 6,421 (3,015 - 11,243) | 0.394 (0.185 - 0.69) | 0.47 (0.37 - 0.58) | 0.25 (0.23 - 0.26) | 80.05 (66.82 - 103.86) |
| Kerman | 7,340 (2,761 - 14,621) | 0.224 (0.084 - 0.446) | 0.46 (0.36 - 0.57) | 0.22 (0.20 - 0.23) | 50.58 (40.64 - 63.22) |
| Kermanshah | 6,844 (3,662 - 11,734) | 0.356 (0.190 - 0.610) | 0.51 (0.39 - 0.63) | 0.27 (0.25 - 0.29) | 66.67 (54.07 - 86.87) |
| Kohgiluyeh & Boyer Ahmad | 1,417 (301 - 3,569) | 0.191 (0.040 - 0.481) | 0.32 (0.26 - 0.39) | 0.20 (0.19 - 0.22) | 59.41 (48.25 - 74.93) |
| Golestan | 6,920 (3,138 - 12,352) | 0.357 (0.162 - 0.630) | 0.35 (0.27 - 0.43) | 0.20 (0.19 - 0.22) | 105.43 (87.31 - 125.82) |
| Gilan | 8,788 (3,888 - 17,078) | 0.362 (0.160 - 0.704) | 0.64 (0.51 - 0.75) | 0.36 (0.34 - 0.39) | 52.62 (45.21 - 67.60) |
| Lorestan | 4,942 (1,895 - 9,850) | 0.280 (0.100 - 0.566) | 0.48 (0.40 - 0.59) | 0.25 (0.23 - 0.27) | 56.51 (47.10 - 68.36) |
| Mazandaran | 10,500 (4,372 - 19,563) | 0.324 (0.130 - 0.603) | 0.55 (0.46 - 0.64) | 0.31 (0.29 - 0.34) | 58.36 (48.01 - 69.78) |
| Markazi | 5,200 (2,273 - 9,601) | 0.367 (0.160 - 0.678) | 0.55 (0.46 - 0.67) | 0.32 (0.29 - 0.34) | 64.71 (53.01 - 75.59) |
| Hormozgan | 3,753 (1,369 - 7,864) | 0.194 (0.070 - 0.407) | 0.42 (0.33 - 0.54) | 0.17 (0.16 - 0.18) | 48.52 (38.76 - 64.10) |
| Hamedan | 5,952 (2,868 - 10,475) | 0.300 (0.168 - 0.615) | 0.53 (0.44 - 0.65) | 0.30 (0.28 - 0.32) | 63.97 (53.45 - 76.72) |
| Yazd | 3,287 (1,572 - 6,475) | 0.270 (0.129 - 0.533) | 0.35 (0.30 - 0.40) | 0.24 (0.22 - 0.26) | 80.50 (71.43 - 93.68) |

\* Based on the estimates from Ghafari et al. [7]. \*\* Year 1394 Solar Hijri is excluded from the calculation of background mortality for this province due to a record-high excess mortality in that year [7].

**Figure 2:**
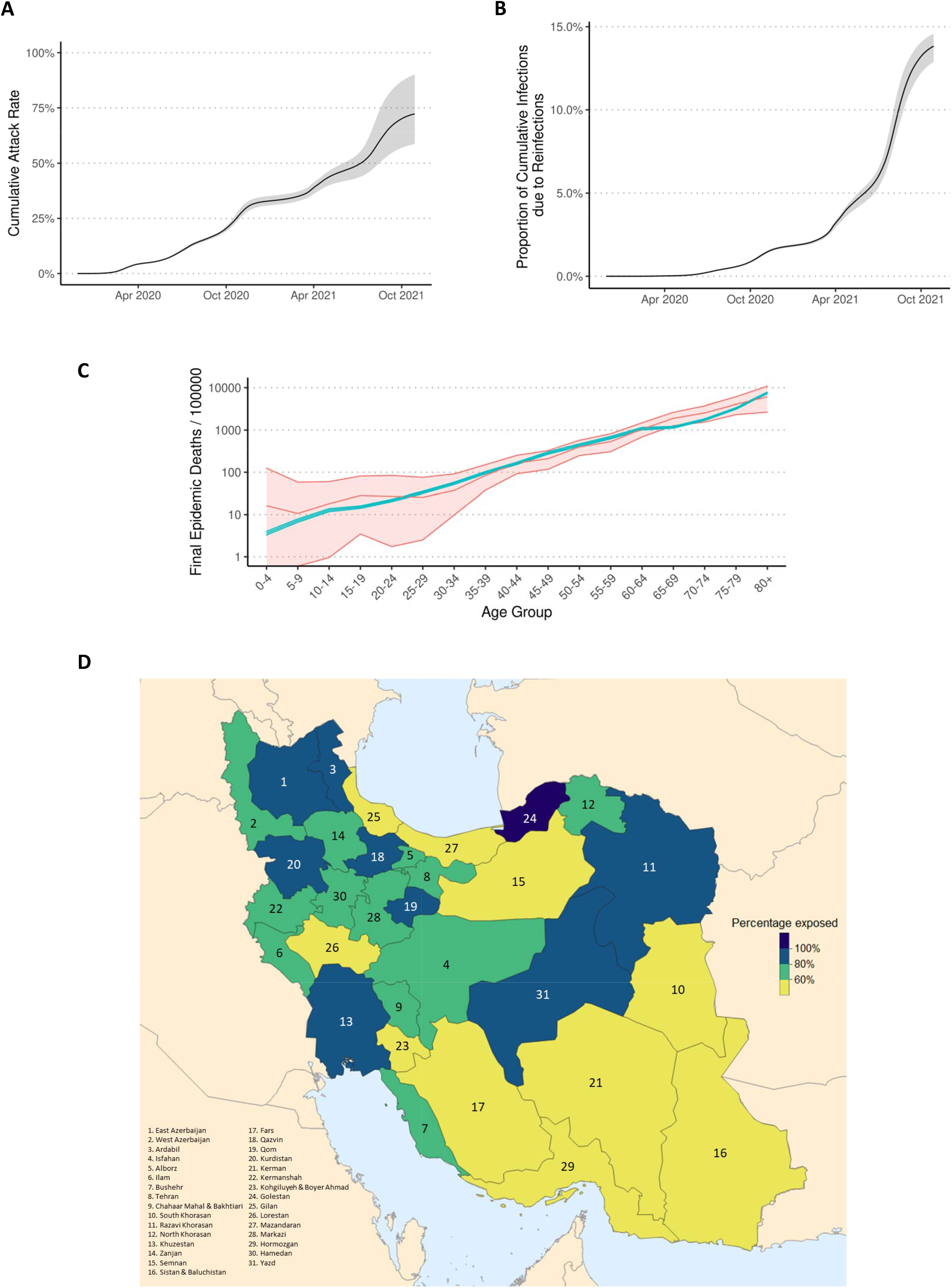
**(A)** Cumulative nationwide attack rate of SARS-CoV-2, **(B)** percentage of individuals reinfected, and **(C)** per-capita excess mortality per age-group according to the transmission model (green) and observed data (red). Shaded area in black shows the estimated quantities from the three sets of model assumptions used for estimating attack rates. Shaded areas in green and red show the 95% confidence interval in modelled and observed excess mortality, respectively. **(D)** Cumulative attack rate of SARS-CoV-2 per province as of 2021-10-22.

Using our transmission model, we also calculate the cumulative attack rate by 2021-10-22 for each province (**Figure 2D**). Our results show a high degree of heterogeneity in the estimated attack rates across the country with 11 provinces reaching close to and higher than 100% attack rates after the fifth wave (**Table S1**). While in most regions, the majority of infections took place during the third and fifth waves, the Delta variant of SARS-CoV-2 was responsible for the highest re-infection rates during the fifth wave (**Figure S5**). Qazvin, Qom, and East Azerbaijan had the highest per capita mortality rate with cumulative PFRs of nearly 0.4%, while Golestan, Khuzestan, and Qom had the highest re-infection rate with nearly 20% of infections being due to re-infection (**Table 1**; **Figure S5**). **Figure S1** and **Figure S3** also show that while some provinces such as Ardabil, Alborz, Semnan, Golestan, Hamedan, and Mazandaran experienced five distinct epidemic peaks, others like Bushehr, Qazvin, Gilan, Kohgiluyeh and Boyer Ahmad, and Hormozgan experienced sustained transmission of SARS-CoV-2 for an extended period of time throughout the pandemic. In late September 2021, official reports from MoHME indicate that COVID-19 deaths in Sistan and Baluchistan, Ardabil, Ilam, Bushehr, Chahaar Mahal and Bakhtiari, South Khorasan, North Khorasan, Zanjan, Kohgiluye and Boyer Ahmad, Lorestan, Hormozgan, and Hamedan have all dropped to below 5 individuals per day in recent weeks, an almost record-low since the first wave in March 2020 [28-30]. This is congruent with our estimates of high attack rates in these provinces which may have resulted in a (temporary) herd-immunity to be reached after the fifth wave.

## Discussion

Since the start of the pandemic, the global heterogeneity in COVID-19 reporting systems globally has resulted in substantial debate about the true extent to which COVID-19 has spread in each country [4, 31, 32]. International comparisons of COVID-19 death tolls have subsequently relied on excess mortality due to under-reporting of COVID-19 deaths in many parts of the world [2]. This study is the first to formally explore the suitability of excess mortality as a proxy for estimating under-reporting of COVID-19 infections and deaths, confirming the information contained within reliable excess mortality statistics through the broad agreement with representative seroprevalence estimates. Additionally, this analysis lends further support to evidence that IFR patterns estimated in high income countries may underestimate the IFR in settings with stretched healthcare capacity [26], as experienced in many provinces in Iran at periods during the pandemic [33, 34]. Even though in some provinces we find that the transmission model with a lower estimated IFR provides a better fit to seroprevalence estimates, such a model offers a poor fit across most provinces and often results in epidemiologically unrealistic outcomes such that the modelled transmission dynamics outruns the pool of susceptible individuals. We also note that while some factors such as misidentification of baseline all-cause mortality and deaths attributable to COVID-19, particularly in the younger age-groups with fewer COVID-19-related deaths, can contribute to varied attack rates in some provinces, other factors such as nonrepresentative serosurveys, varied time to seroreversion using different assays, and uncertainty in inferring the true contact matrix between different age-groups within the population can impact the inferred attack rates [35, 36].

Prior to the emergence of the variants of concern, the basic reproduction number of the SARS-CoV-2 virus was estimated to be around 2.8 which corresponds to a herd immunity threshold of about 64% [37]. This threshold does not represent the point at which the outbreak immediately stops, but rather the point where, on average, fewer secondary infections are produced. During unmitigated outbreaks with such reproduction numbers, the attack rate can reach as high as 90% [38, 39]. Similarly, during unmitigated outbreaks with variants of concerns such as the Delta variant with increased reproduction numbers compared to the ancestral SARS-CoV-2 virus, the attack rate can reach even higher levels [40]. While the duration of antibody responses after coronavirus infection over longer timescales remains largely unknown, recent surveillance data and comparative studies have shown that re-infection with SARS-CoV-2 is possible after 3-6 months, and that older age-groups have a much lower protection against reinfection [41, 42]. Previous studies on human coronaviruses and SARS-CoV have also shown that protective immunity typically starts declining after 5-8 months post infection [43-45]. Our findings of high attack rates in several provinces show that herd immunity through natural infection has not been achieved in the population even after nearly 20 months since the start of the Iranian epidemic. This is likely due to substantial reduction in protection against repeat infection over time either due to waning immunity, increased chance of re-infection with variants of concern, or a combination of both. We note that while systematic biases such as identifying the baseline level of all-cause mortality and overestimation of deaths attributable to COVID-19, particularly in the younger age-groups, can contribute to increased attack rates in some provinces, other factors such as heterogeneities in immune protection, limited access to health care in municipalities with low socioeconomic status, and uneven adoption of public health intervention in different regions can also contribute to varied attack rates across the country.

## Methods

### Excess mortality

We collect the weekly time-series data on all-cause mortality per province per age group from NOCR. We use data from the beginning of year 1394 to the end of summer 1398 in Solar Hijri calendar (SH) (from 2015-03-15 to 2019-09-22 in Common Era) to calculate background mortality. We exclude mortality data during autumn 2019 (from 2019-09-23 to 2019-12-21) from the calculation of background mortality as we have previously reported elevated mortality rates across several provinces, unrelated to the COVID-19 pandemic, which can bias our estimates for the expected mortality [7]. We calculate the expected mortality using a linear regression model that accounts for seasonality and trend, previously developed to track excess mortality across more than 100 countries and territories around the world [1].

### Transmission model

We use a previously published age-structured COVID-19 transmission model [46] and fitting framework [24] to fit the weekly estimated excess deaths in each province in Iran. In overview, the model is a population-based age-structured Susceptible-Exposed-Infected-Recovered model, which explicitly represents disease severity, passage through different healthcare levels and the roll out of vaccination. Model fitting was carried out within a Bayesian framework using a Metropolis-Hastings Markov Chain Monte based sampling scheme, which estimates the epidemic start date, *R*_*0*_ and the time varying reproduction number, *R*_*t*_, using a series of pseudo-random walk parameters, which alter transmission every 2-weeks, given by:

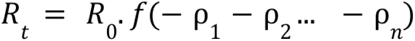

where *f(x) = 2.exp(x)/(1+exp(x))*, i.e., twice the inverse logit function. Each parameter is introduced two weeks after the previous parameter, serving to capture changes in transmission every two weeks. The last change in transmission, *ρ*_*n*_, is maintained for the last 4 weeks prior to the current day to reflect our inability to estimate the effect size of this parameter due to the approximate 21-day delay between infection and death [47].

Each model fit used province-specific demography, with the population size in 5-year age bands sourced from the Statistical Center of Iran [48]. The effective number of general hospital beds and intensive care beds were sourced from the Iran statistical yearbook published in 1396 in Solar Hijri calendar [49]. At the time of writing, MoHME has not published subnational daily vaccination data prior to 2021-10-26, nor the proportion of which vaccines are administered. As a result, we used multiple data sources to approximate the roll of vaccines subnationally. From 2021-10-26, MoHME started to release the total number of first and second vaccine doses subnationally, which we use in combination with national vaccination data [50] to infer the daily vaccination rate per province. A mix of vaccine types have been administered in Iran, with 80% of the population vaccinated with Sinopharm, 10% with AZ and the remaining 10% either Sputkink-V (1.5%) and CoViran Barekat (8.5%). Due to the mix of vaccine types with uncertain efficacy estimates, particularly related to Delta, we use vaccine efficacy estimates for first and second dose efficacy sourced from estimates for the Sinopharm vaccine and other inactivated whole virus vaccine platforms (**Table 2**). Efficacy estimates are weighted to produce an overall population efficacy by the proportion of vaccinated individuals who have received either a first dose or second dose, and by the assumed proportion of the Delta variant in the population, sourced from the sampling dates of Delta sequences in Iran from GISAID (*gisaid.org*), lagged by 14 days to reflect the delay from administering vaccine to protection being conferred.

**Table 2:**
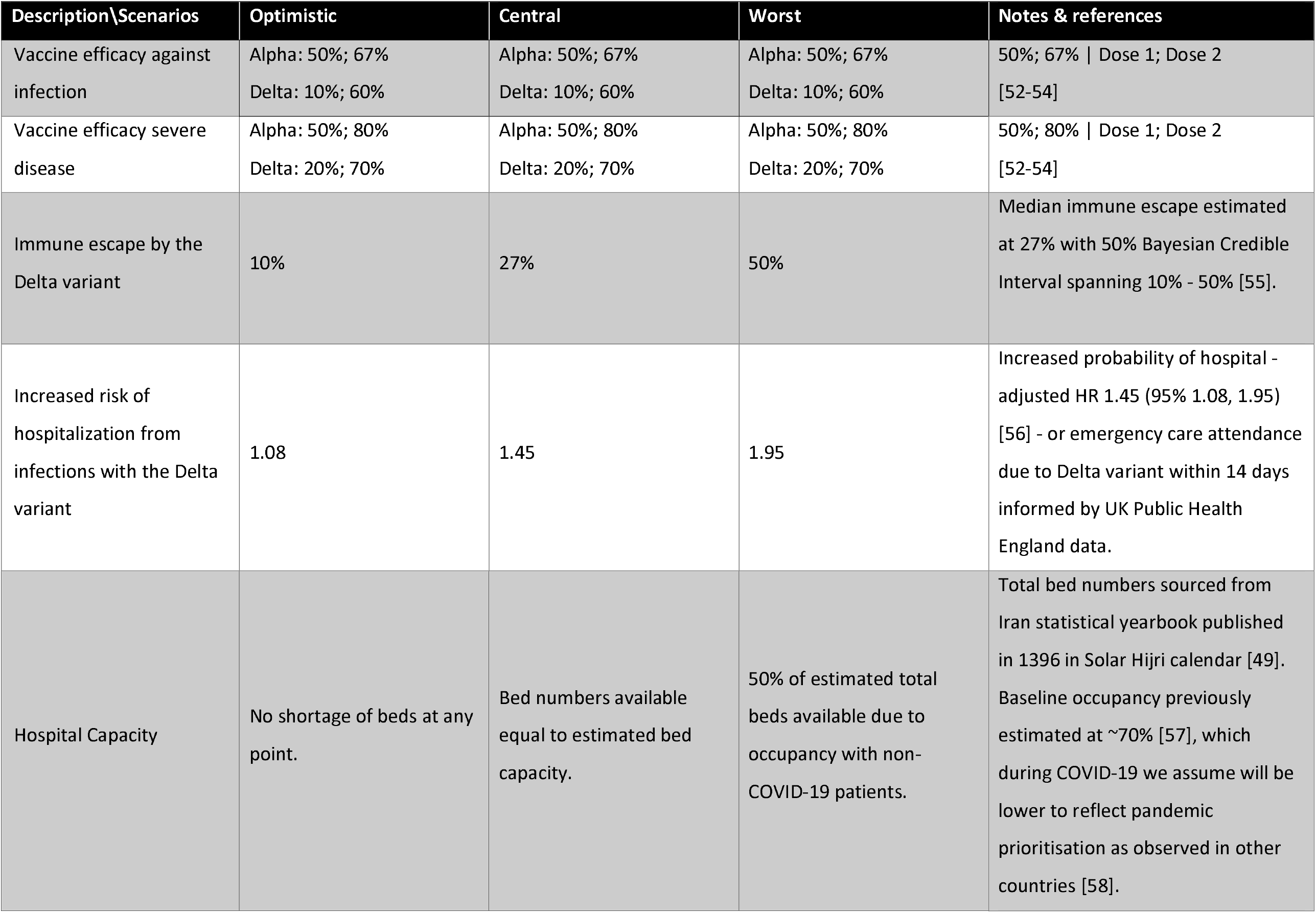
Vaccine efficacy estimates, and parameter uncertainty related to hospitalisation and the Delta variant.

To estimate the attack rate over time, we explore three sets of model assumptions, which scan across the likely range of parameter estimates for key model parameters that impact the attack rate inferred. These parameters relate to the assumed number of general and ICU hospital beds available, the change in hospitalisation related to the Delta variant and the level of immune escape conferred by the Delta variant relative to previous infection by the Alpha variant and the original wild type strains. These ranges are described in **Table 2**. Given uncertainty in the IFR and its impact of the inferred attack rate, we explore two alternative estimates of IFR by age [21, 51] and conduct model comparison between the two references through comparison against province level estimated seroprevalence data [8], with seroprevalence assumed to be described by a Binomial distribution. Lastly, to explore if IFR is changing over time, we explore the ratio of model inferred hospitalisations and observed daily hospitalisations against time using a mixed-effects model, controlling for variation between provinces using a random intercept and slope with respect to time.

## Data Availability

All the data used for the analysis are available online on our GitHub repository (https://github.com/mg878/Iran_WeeklyMortality), with all model fitting code and analysis available at (https://github.com/OJWatson/iran-ascertainment). For regular updates on excess mortality in Iran, you can visit: https://github.com/akarlinsky/world_mortality.

https://github.com/OJWatson/iran-ascertainment

https://github.com/akarlinsky/world_mortality

## Authors contribution

Conceptualization: MG, AK, OJW, LF; Methodology: MG, OJW, AKar; Investigation: MG, OJW; Visualization: MG, OJW; Supervision: AK; Writing – original draft: MG; Writing – review & editing: All authors.

## Extended data

### Supplementary Tables

**Table S1:** The overall attack rate (in percentage) by the end of May 2020 (wave 1), August 2020 (wave 2), January 2021 (wave 3), June 2021 (wave 4), and the week ending on 22 October 2021 (wave 5).

| Province | Wave 1 | Wave 2 | Wave 3 | Wave 4 | Wave 5 |
| --- | --- | --- | --- | --- | --- |
| East Azerbaijan | 5.44 (5.50 - 5.14) | 11.68 (11.86 - 11.45) | 39.85 (37.39 - 42.29) | 60.36 (52.28 - 64.14) | 84.92 (69.06 - 103.05) |
| West Azerbaijan | 4.06 (3.85 - 4.34) | 10.23 (8.87 - 10.50) | 37.85 (34.37 - 42.99) | 52.94 (45.84 - 60.46) | 75.66 (60.25 - 98.53) |
| Ardabil | 11.18 (9.90 - 11.52) | 23.57 (20.61 - 26.34) | 51.23 (45.54 - 54.60) | 69.27 (60.02 - 77.94) | 92.55 (76.19 - 112.14) |
| Isfahan | 5.58 (4.92 - 5.28) | 9.95 (8.66 - 9.62) | 30.09 (26.10 - 30.96) | 42.93 (37.36 - 45.99) | 65.90 (51.80 - 81.46) |
| Alborz | 5.32 (5.44 - 5.16) | 10.50 (10.27 - 10.48) | 27.44 (25.99 - 29.10) | 45.49 (38.88 - 51.17) | 64.09 (51.07 - 81.97) |
| Ilam | 4.49 (4.13 - 4.23) | 10.78 (10.17 - 10.55) | 34.75 (31.70 - 36.22) | 44.19 (40.23 - 47.35) | 61.35 (51.01 - 73.74) |
| Bushehr | 3.20 (2.53 - 3.24) | 11.52 (8.40 - 11.39) | 25.13 (20.63 - 25.25) | 40.97 (32.96 - 45.54) | 61.29 (44.63 - 82.10) |
| Tehran | 7.69 (8.31 - 8.07) | 12.67 (14.37 - 13.17) | 32.42 (33.46 - 34.66) | 51.17 (46.13 - 54.27) | 77.06 (65.08 - 93.99) |
| Chahaar Mahal & Bakhtiari | 7.06 (6.53 - 6.49) | 13.46 (12.52 - 13.15) | 40.07 (36.20 - 45.02) | 56.42 (49.60 - 67.55) | 76.28 (63.01 - 101.11) |
| South Khorasan | 3.11 (2.94 - 3.49) | 5.72 (5.26 - 6.53) | 21.59 (19.20 - 23.92) | 30.87 (27.58 - 33.24) | 41.27 (34.65 - 48.50) |
| Razavi Khorasan | 6.77 (6.27 - 6.66) | 19.33 (17.53 - 21.33) | 43.41 (38.66 - 45.49) | 57.06 (49.79 - 64.07) | 90.19 (71.76 - 113.32) |
| North Khorasan | 6.46 (6.06 - 5.73) | 14.52 (12.58 - 13.86) | 36.11 (32.34 - 40.32) | 48.61 (41.48 - 53.53) | 77.73 (61.94 - 100.78) |
| Khuzestan | 11.17 (10.51 - 13.65) | 26.08 (23.70 - 29.49) | 40.38 (36.19 - 42.13) | 68.54 (58.79 - 76.98) | 98.48 (79.82 - 120.59) |
| Zanjan | 6.85 (7.59 - 7.15) | 14.10 (14.18 - 14.59) | 38.23 (35.99 - 41.37) | 56.79 (51.31 - 63.59) | 73.11 (60.85 - 87.84) |
| Semnan | 4.12 (4.59 - 4.19) | 8.27 (9.25 - 8.31) | 30.87 (29.84 - 31.08) | 42.82 (39.75 - 45.06) | 57.26 (49.85 - 65.92) |
| Sistan & Baluchistan | 1.24 (1.23 - 1.11) | 5.02 (5.51 - 5.35) | 14.02 (14.85 - 14.14) | 26.46 (22.98 - 32.57) | 46.60 (35.38 - 67.60) |
| Fars | 1.64 (1.61 - 1.59) | 6.51 (6.04 - 7.08) | 25.69 (23.57 - 29.32) | 38.77 (33.97 - 43.35) | 56.63 (42.64 - 71.82) |
| Qazvin | 12.92 (12.21 - 13.66) | 19.79 (18.21 - 20.70) | 46.85 (41.92 - 50.43) | 64.95 (55.77 - 72.23) | 85.97 (69.52 - 106.44) |
| Qom | 14.87 (14.23 - 18.22) | 20.10 (18.55 - 24.31) | 40.20 (36.16 - 47.18) | 51.89 (45.12 - 61.98) | 84.74 (66.12 - 116.36) |
| Kurdistan | 7.66 (6.86 - 7.83) | 17.13 (16.40 - 18.52) | 45.93 (43.35 - 55.61) | 59.55 (54.04 - 69.37) | 80.05 (66.82 - 103.86) |
| Kerman | 2.10 (2.17 - 2.15) | 8.11 (7.99 - 9.06) | 23.45 (22.19 - 24.80) | 32.84 (29.26 - 34.56) | 50.58 (40.64 - 63.22) |
| Kermanshah | 4.08 (4.39 - 4.31) | 9.97 (9.86 - 9.13) | 30.16 (28.50 - 32.97) | 41.38 (37.50 - 46.08) | 66.67 (54.07 - 86.87) |
| Kohgiluyeh & Boyer Ahmad | 4.24 (3.88 - 3.79) | 9.37 (8.61 - 8.72) | 27.38 (24.45 - 27.12) | 42.16 (36.62 - 46.16) | 59.41 (48.25 - 74.93) |
| Golestan | 20.27 (18.89 - 21.60) | 33.20 (30.69 - 36.89) | 55.32 (48.44 - 56.16) | 71.67 (61.89 - 75.25) | 105.43 (87.31 - 125.82) |
| Gilan | 15.51 (15.13 - 17.90) | 17.50 (17.26 - 20.12) | 28.36 (27.51 - 31.21) | 36.79 (34.53 - 40.22) | 52.62 (45.21 - 67.60) |
| Lorestan | 5.37 (5.64 - 5.41) | 11.81 (12.20 - 12.09) | 28.87 (27.28 - 30.13) | 40.94 (36.45 - 43.29) | 56.51 (47.10 - 68.36) |
| Mazandaran | 11.19 (10.15 - 11.53) | 18.16 (16.62 - 18.97) | 31.36 (29.63 - 32.70) | 40.39 (37.42 - 42.43) | 58.36 (48.01 - 69.78) |
| Markazi | 5.86 (5.45 - 5.22) | 11.31 (10.48 - 10.60) | 33.26 (31.29 - 33.89) | 47.89 (42.32 - 51.57) | 64.71 (53.01 - 75.59) |
| Hormozgan | 3.51 (3.55 - 4.10) | 12.58 (11.96 - 13.92) | 21.11 (20.27 - 22.03) | 32.48 (27.99 - 35.42) | 48.52 (38.76 - 64.10) |
| Hamedan | 3.74 (3.79 - 3.78) | 10.69 (10.87 - 11.16) | 28.74 (26.68 - 29.71) | 45.23 (40.47 - 48.63) | 63.97 (53.45 - 76.72) |
| Yazd | 11.26 (12.05 - 11.49) | 17.10 (18.44 - 17.86) | 53.88 (51.44 - 59.97) | 64.11 (59.99 - 72.87) | 80.50 (71.43 - 93.68) |

### Supplementary Figures

**Figure S1:**
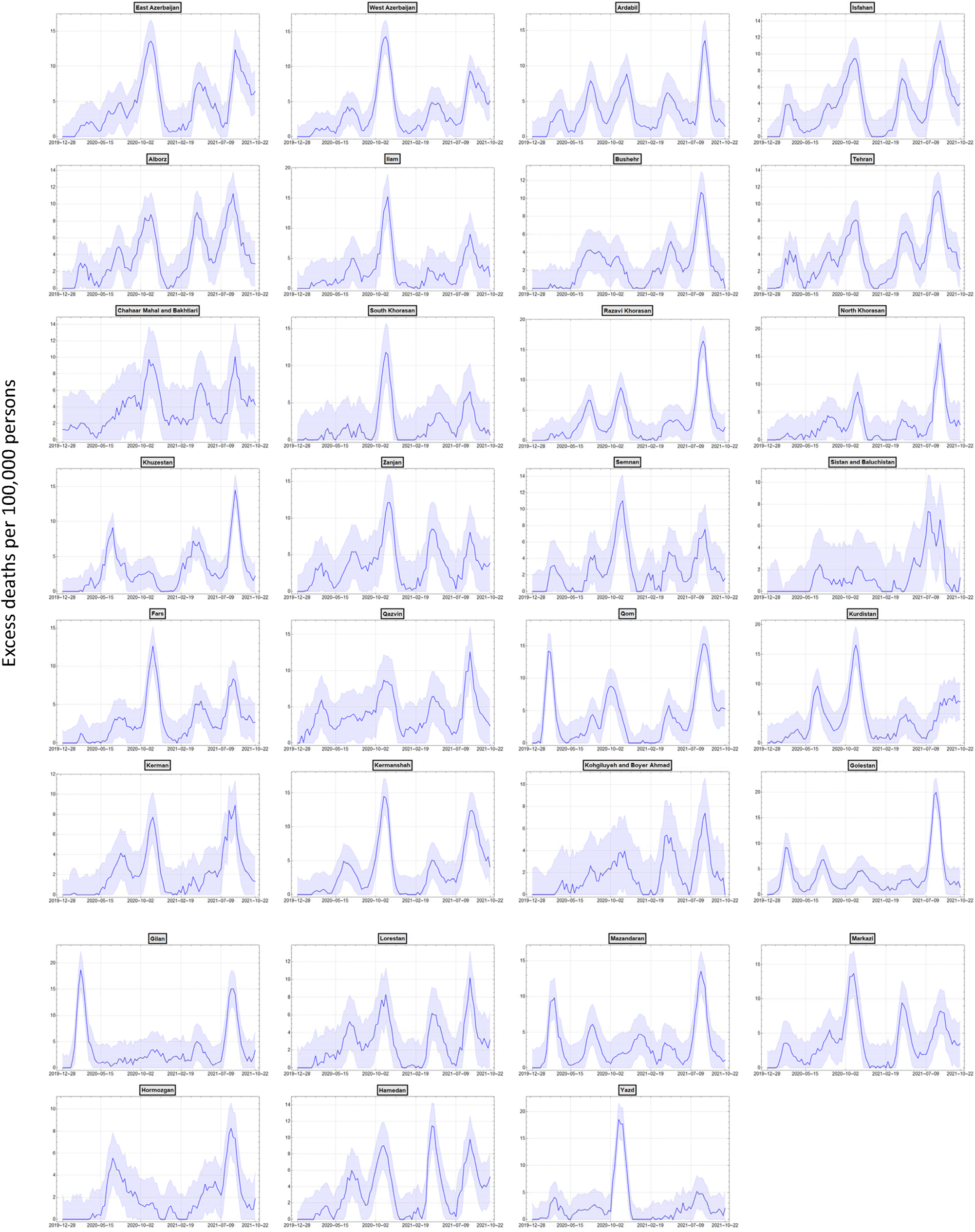
One-month average of weekly excess mortality per 100,000 persons per province over time. Shaded area shows the 95% forecast intervals.

**Figure S2:**
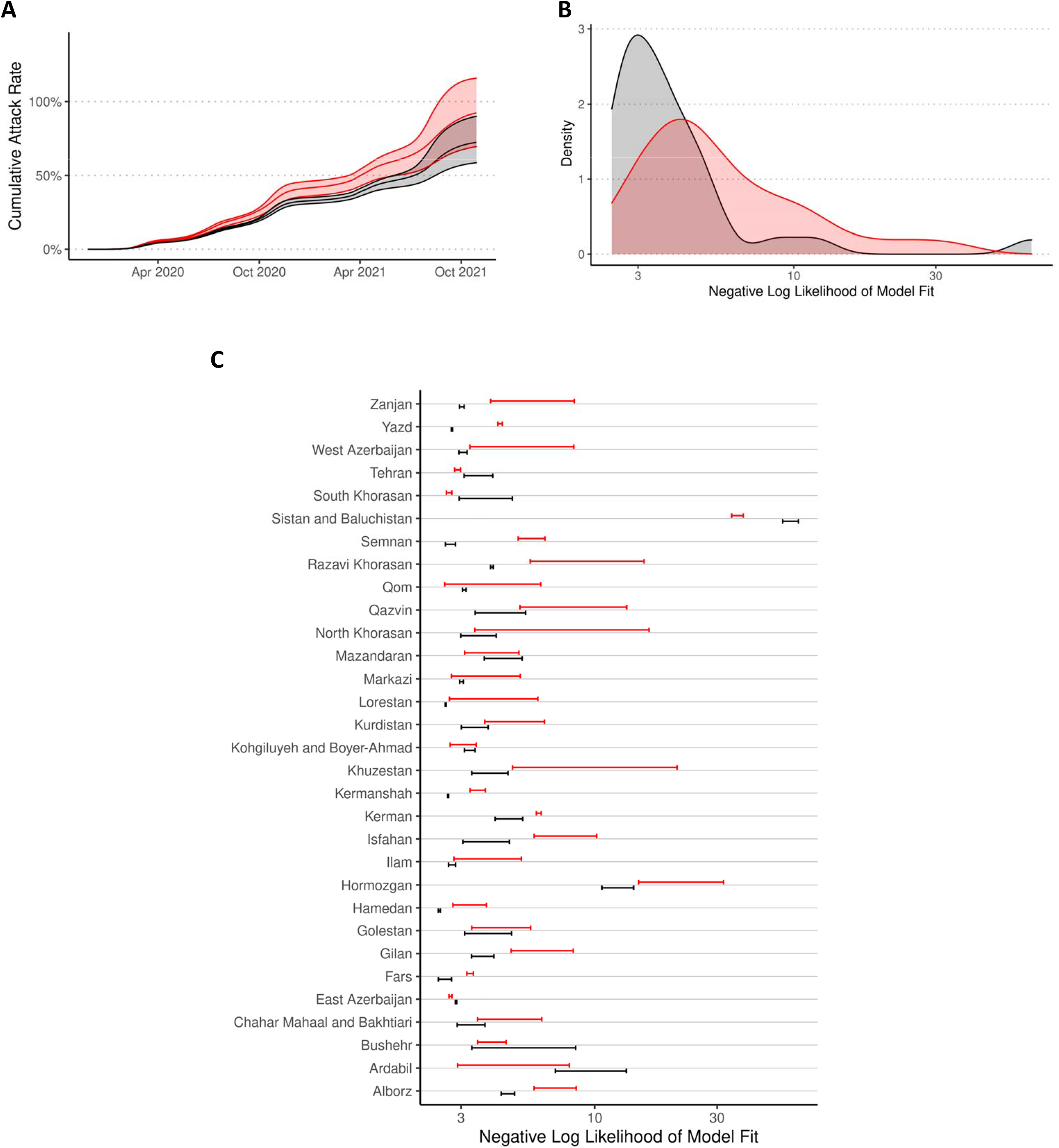
**(A)** Cumulative nationwide attack rates of SARS-CoV-2. Shaded area shows the variation in estimates from the three sets of model assumptions used for estimating attack rates. **(B, C)** Likelihood distribution of the modelled attack rates per province against seroprevalence data [8] using the O’Drsicoll et al. [21] (red) and Brazeau et al. [51] (black) age-stratified infection fatality rate estimates.

**Figure S3:**
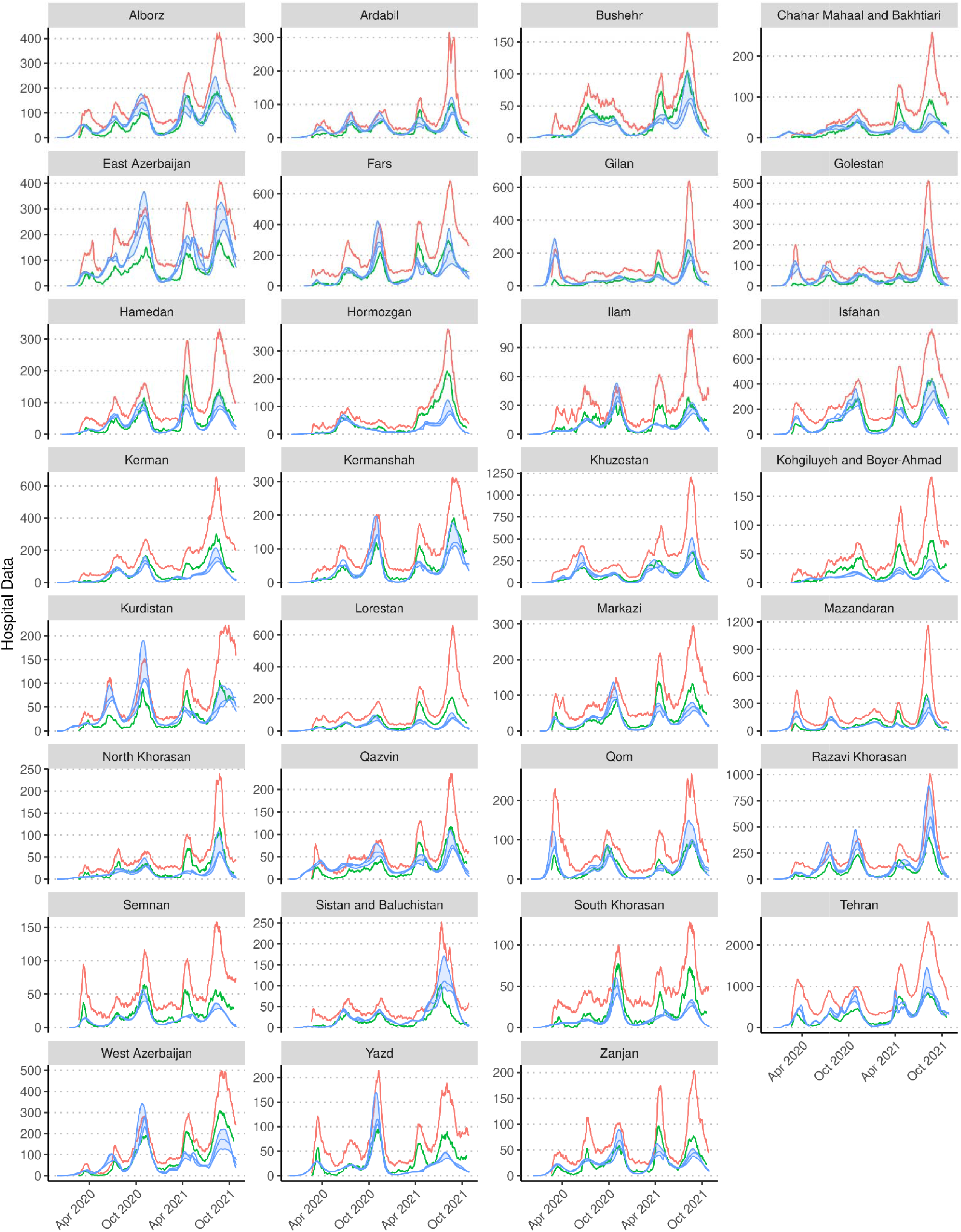
Confirmed (green), suspected (red), and modelled (blue) daily hospital admissions per province over time. Shaded area in blue shows the variation in modelled daily hospital admissions from the three sets of model assumptions used for estimating attack rates.

**Figure S4:**
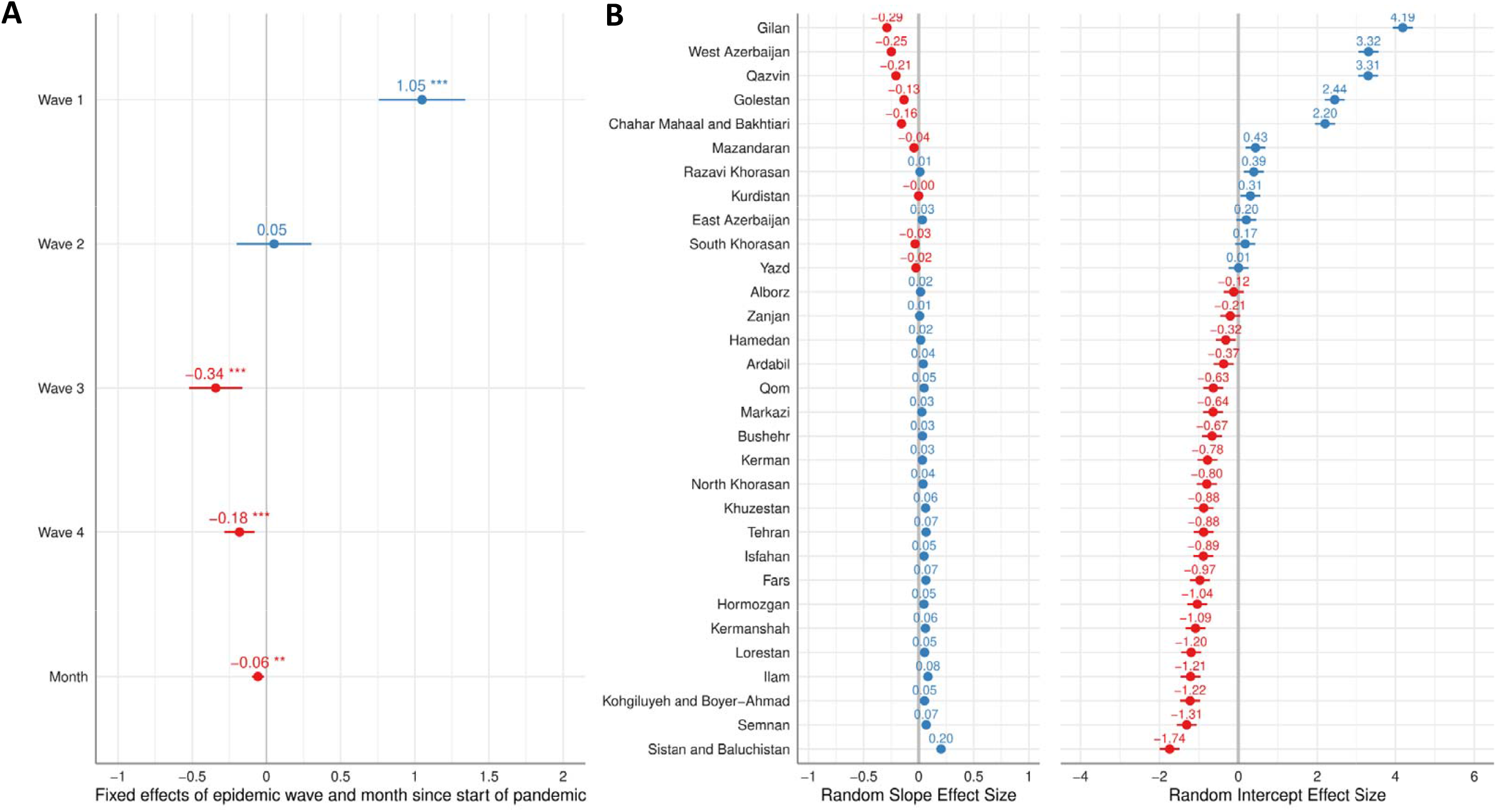
Mixed effect model results investigating the relationship between model inferred hospitalisations and observed daily hospital admissions. In **(A)** the fixed effect sizes are shown, with positive point estimates in blue and negative point estimates in red, with significant effects indicated with asterisks (*p<0.05, **p<0.01 and ***p<0.001%). In **(B)** the random effect sizes are shown for each province ranked by the size of the random intercept for each province.

**Figure S5:**
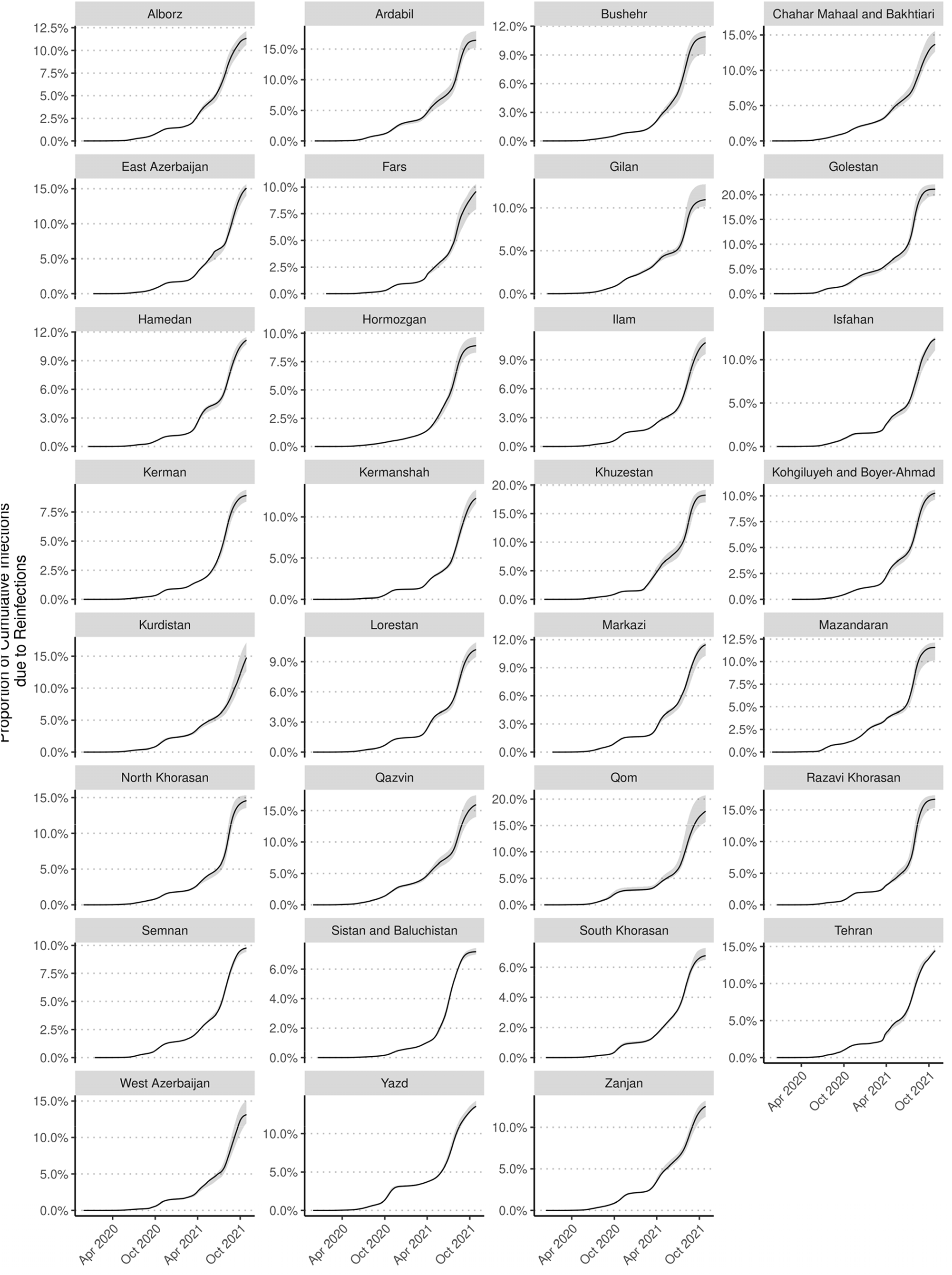
Estimated proportion of infections due to reinfections per province over time. Shaded area shows the variation in estimates from the three sets of model assumptions used for estimating attack rates.

### Supplementary Data

**Data S1-S6:** Transmission model fits per province using weekly excess mortality data (as at week ending on 2021-10-21) under the **(S1)** central, **(S2)** optimistic, and **(S3)** worst scenarios using the Brazeau et al. IFR [51] and the **(S4)** central, **(S5)** optimistic, and **(S6)** worst scenarios using the O’Driscoll et al. IFR [21] estimates. For each province, there are 5 panels showing the model fits for: (i) The time-varying effective reproduction number, Reff, in green. The curve in blue shows the predicted decrease in Reff due to increasing immunity in the population as a result of people being infected by COVID-19. Dark green and blue show the 50% confidence intervals and light green and blue show the 95% confidence intervals. A horizontal dashed line is shown at Reff < 1 indicates a slowing epidemic in which new infections are not increasing. Reff > 1 indicates a growing epidemic in which new infections are increasing over time. (ii) Observed (black circles) and modelled (red line) weekly deaths, with the thick red line depicting the median, the faint depicting individual draws from the posterior and the 95% credible interval shown with dashed black lines. (iii) Modelled cumulative deaths over time. (iv) Modelled seroprevalence (black line) and point estimates by Khalagi et al. [8] (red) and Poustchi et al. [9] (green). Shaded area shows the 95% confidence in modelled seroprevalence. Vertical and horizontal lines around each point estimate represents uncertainty in the reported measurement. (v) Modelled proportion of people exposed to SARS-CoV-2 over time. In both (iv) and (v), the central line depicts the median and the lighter shaded band shows the 95% credible interval.

