## Supplementary data S1 to S6 for "A framework for reconstructing SARS-CoV-2 transmission dynamics using excess mortality data": fitting_vacc_orderly_bundles_derived_vaccine_complete_worst.pdf

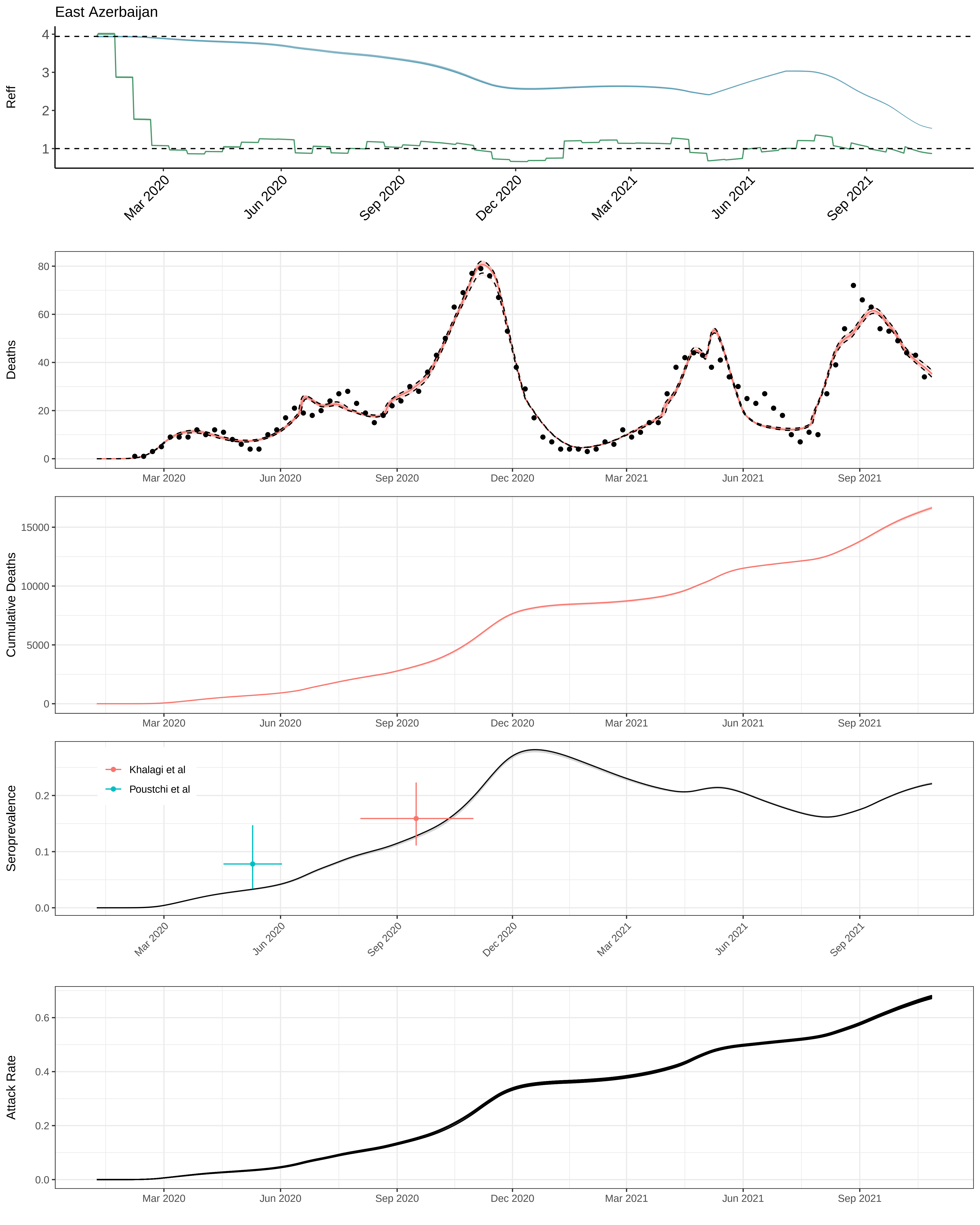

East Azerbaijan
Reff
Deaths
Cumulative Deaths
Seroprevalence
Khalagi et al
Poustchi et al
Attack Rate
Mar 2020
Jun 2020
Sep 2020
Dec 2020
Mar 2021
Jun 2021
Sep 2021

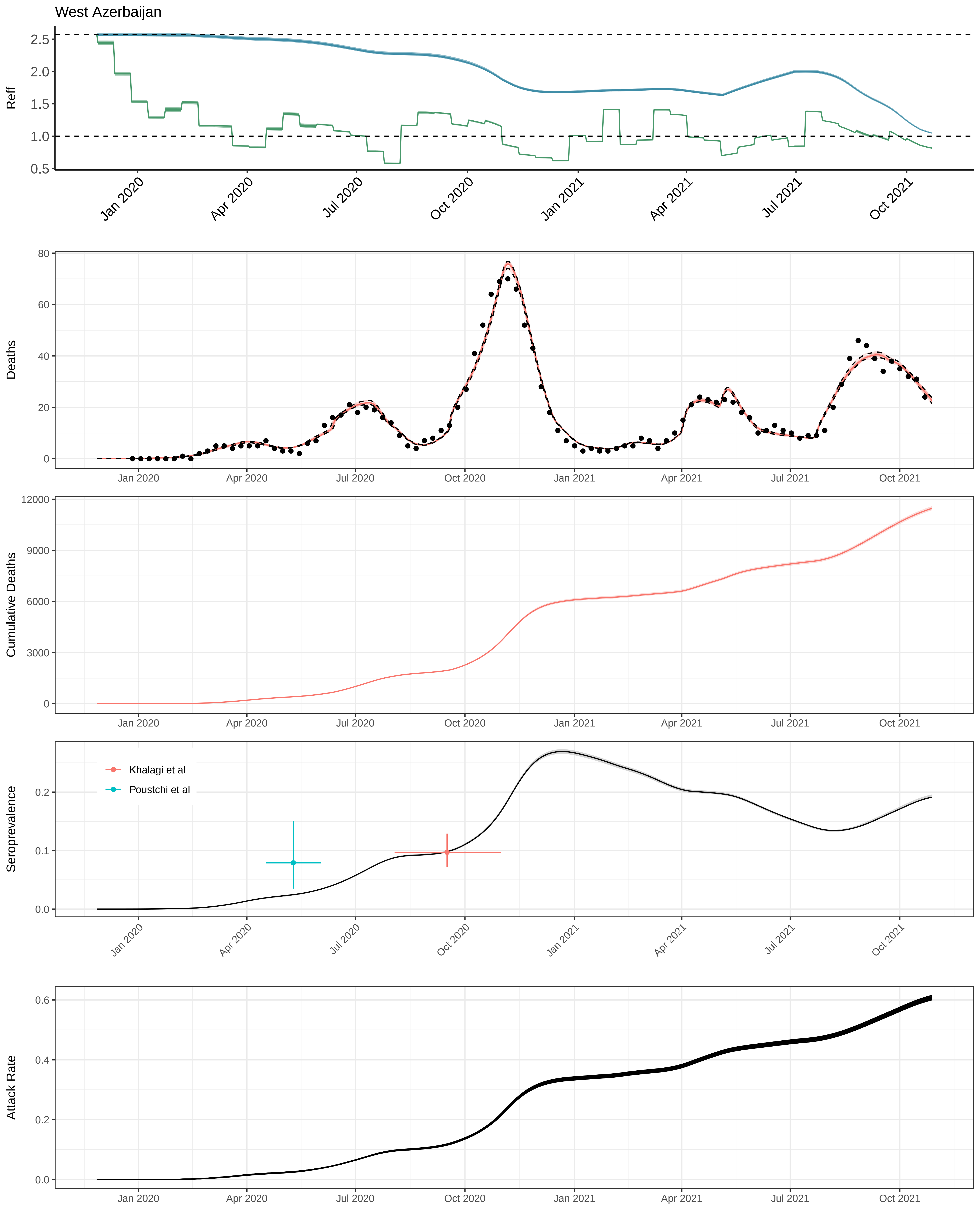

West Azerbaijan
Reff
Deaths
Cumulative Deaths
Seroprevalence
Khalagi et al
Poustchi et al
Attack Rate
Jan 2020
Apr 2020
Jul 2020
Oct 2020
Jan 2021
Apr 2021
Jul 2021
Oct 2021

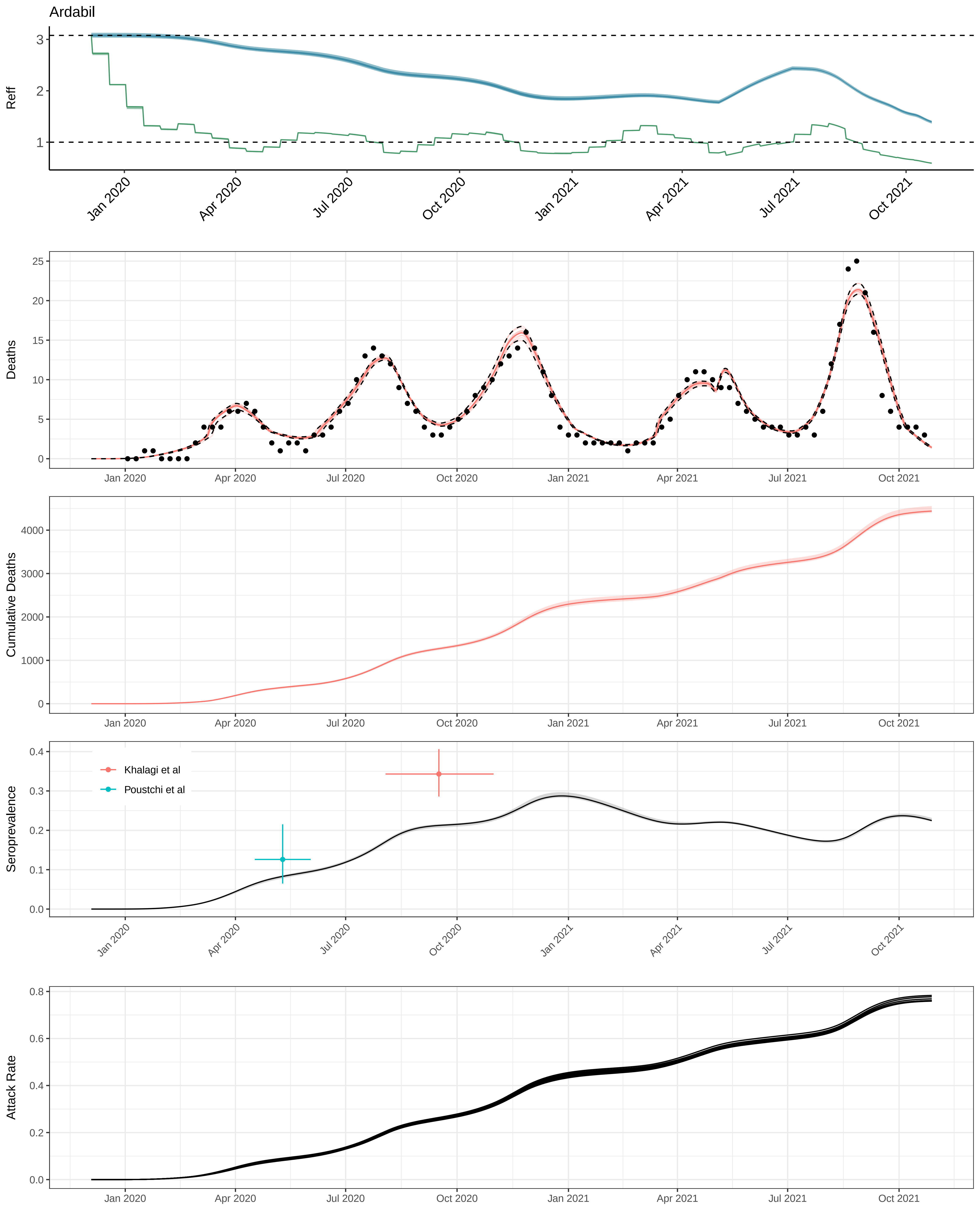

Ardabil
Reff
Deaths
Cumulative Deaths
Seroprevalence
Khalagi et al
Poustchi et al
Attack Rate
Jan 2020
Apr 2020
Jul 2020
Oct 2020
Jan 2021
Apr 2021
Jul 2021
Oct 2021

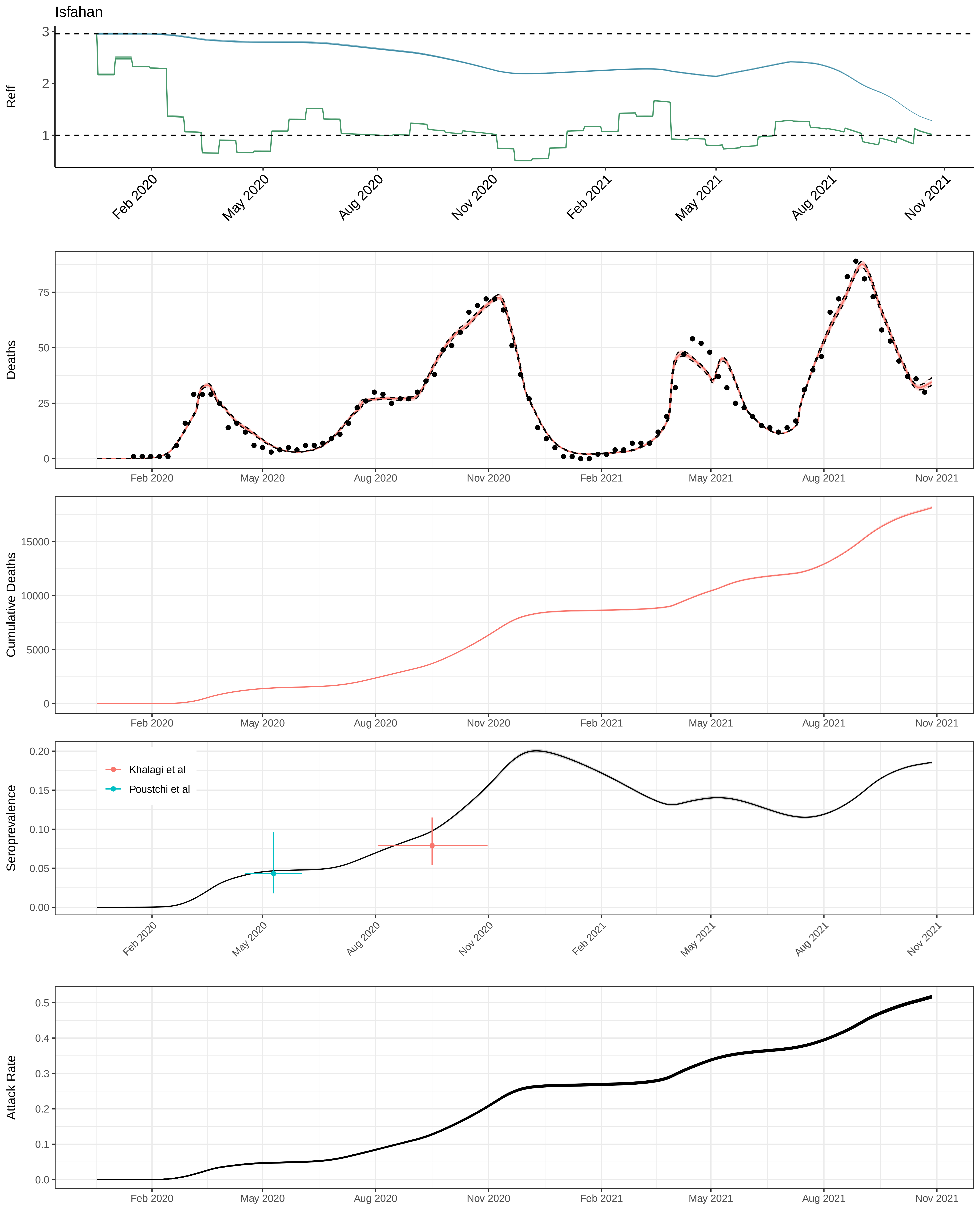
Isfahan
Reff
Deaths
Cumulative Deaths
Seroprevalence
Khalagi et al
Poustchi et al
Attack Rate
Feb 2020
May 2020
Aug 2020
Nov 2020
Feb 2021
May 2021
Aug 2021
Nov 2021

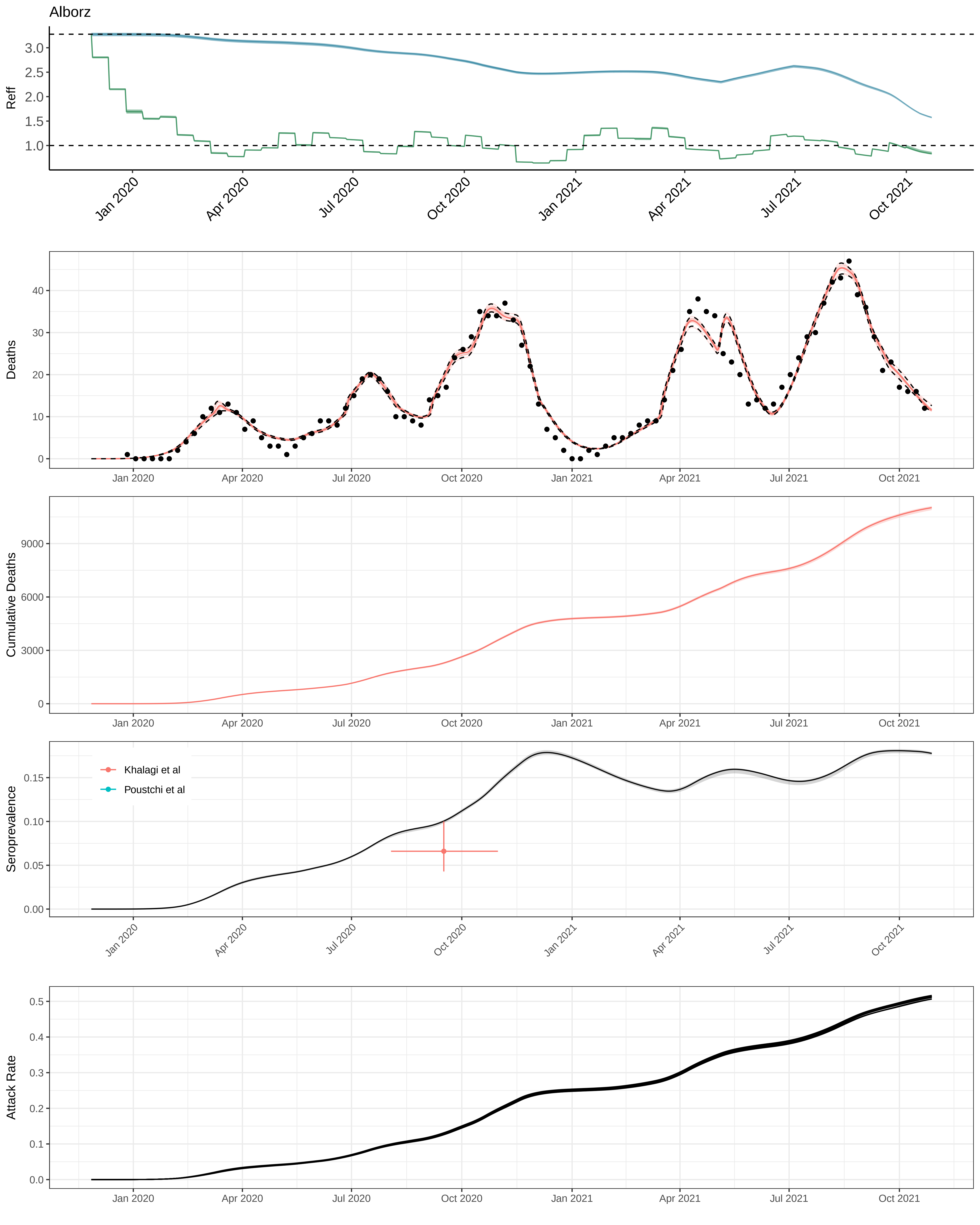
Alborz
Reff
Deaths
Cumulative Deaths
Seroprevalence
Khalagi et al
Poustchi et al
Attack Rate
Jan 2020
Apr 2020
Jul 2020
Oct 2020
Jan 2021
Apr 2021
Jul 2021
Oct 2021

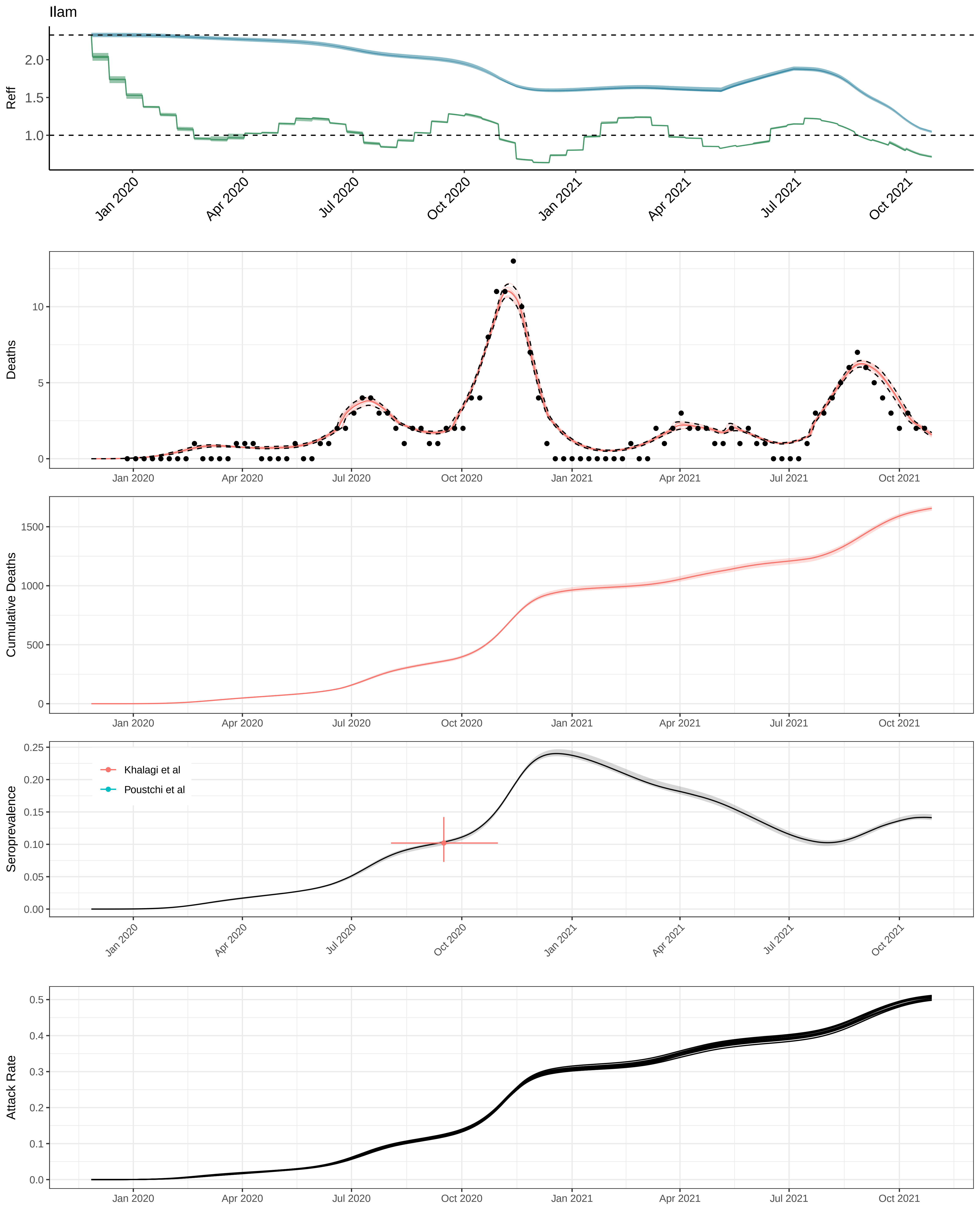
Ilam
Reff
2.0
1.5
1.0
Jan 2020
Apr 2020
Jul 2020
Oct 2020
Jan 2021
Apr 2021
Jul 2021
Oct 2021
Deaths
10
5
0
Cumulative Deaths
1500
1000
500
0
Seroprevalence
0.25
0.20
0.15
0.10
0.05
0.00
Khalagi et al
Poustchi et al
Attack Rate
0.5
0.4
0.3
0.2
0.1
0.0

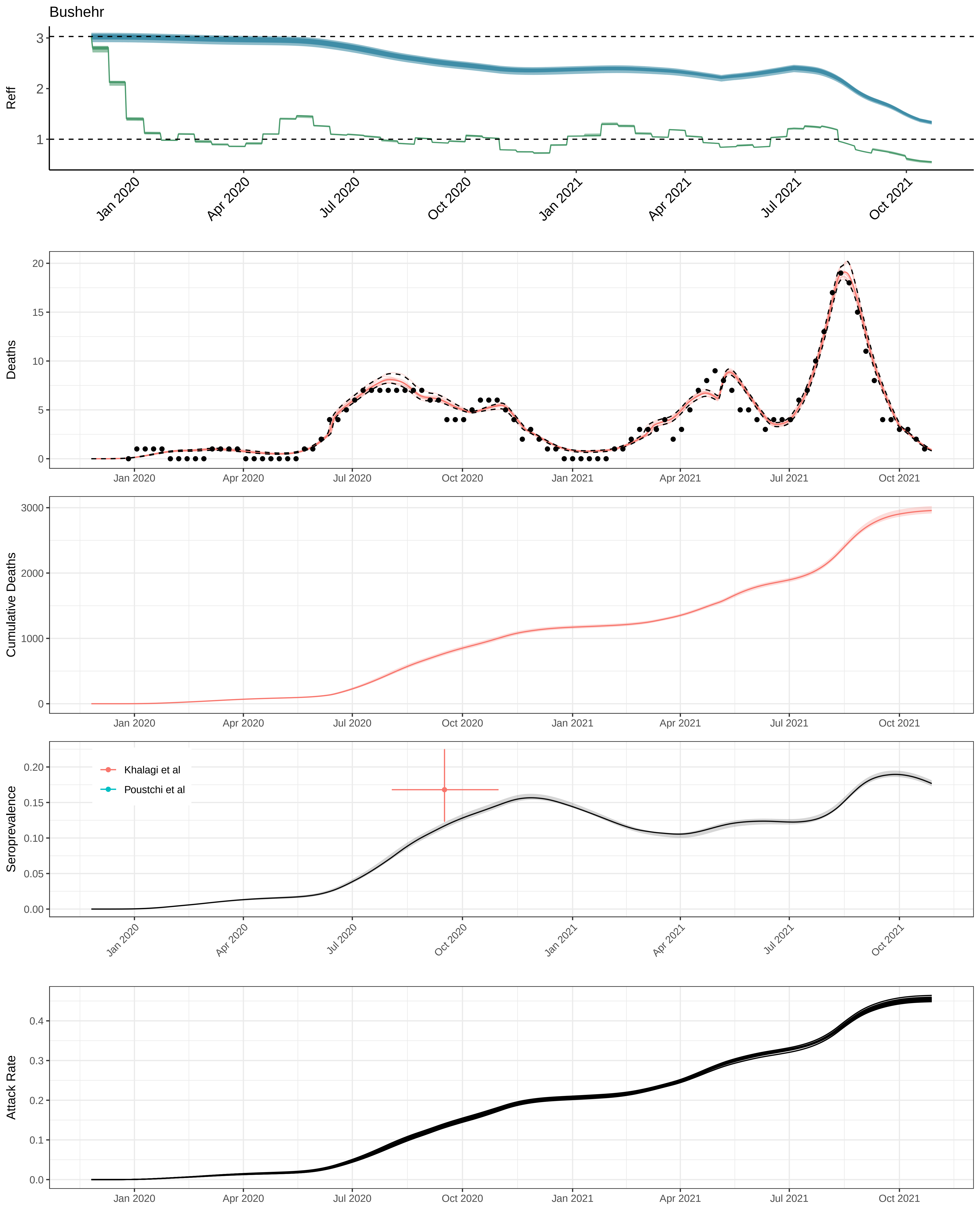

Bushehr
Reff
Deaths
Cumulative Deaths
Seroprevalence
Khalagi et al
Poustchi et al
Attack Rate
Jan 2020
Apr 2020
Jul 2020
Oct 2020
Jan 2021
Apr 2021
Jul 2021
Oct 2021

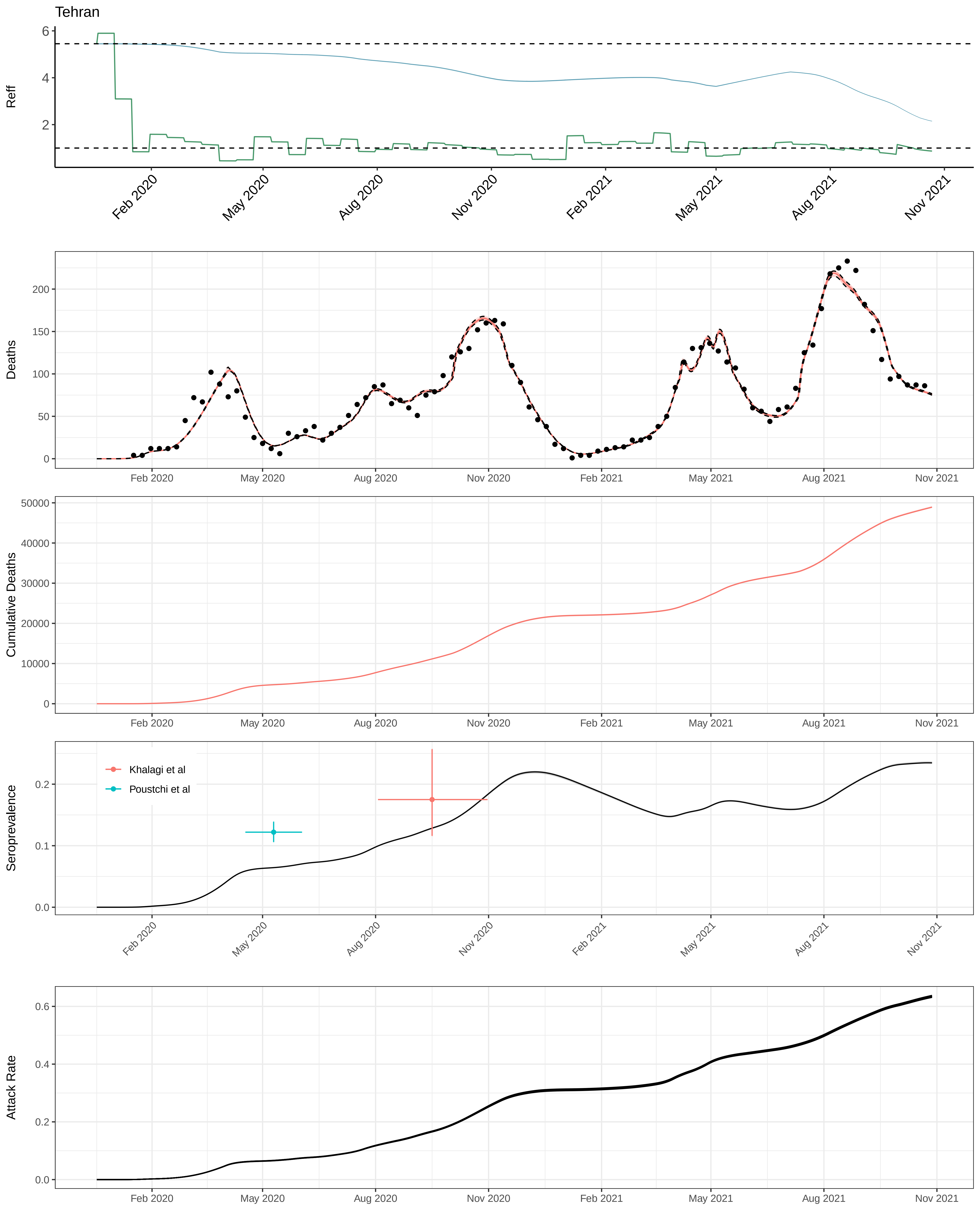
Tehran
Reff
Deaths
Cumulative Deaths
Seroprevalence
Khalagi et al
Poustchi et al
Attack Rate
Feb 2020
May 2020
Aug 2020
Nov 2020
Feb 2021
May 2021
Aug 2021
Nov 2021

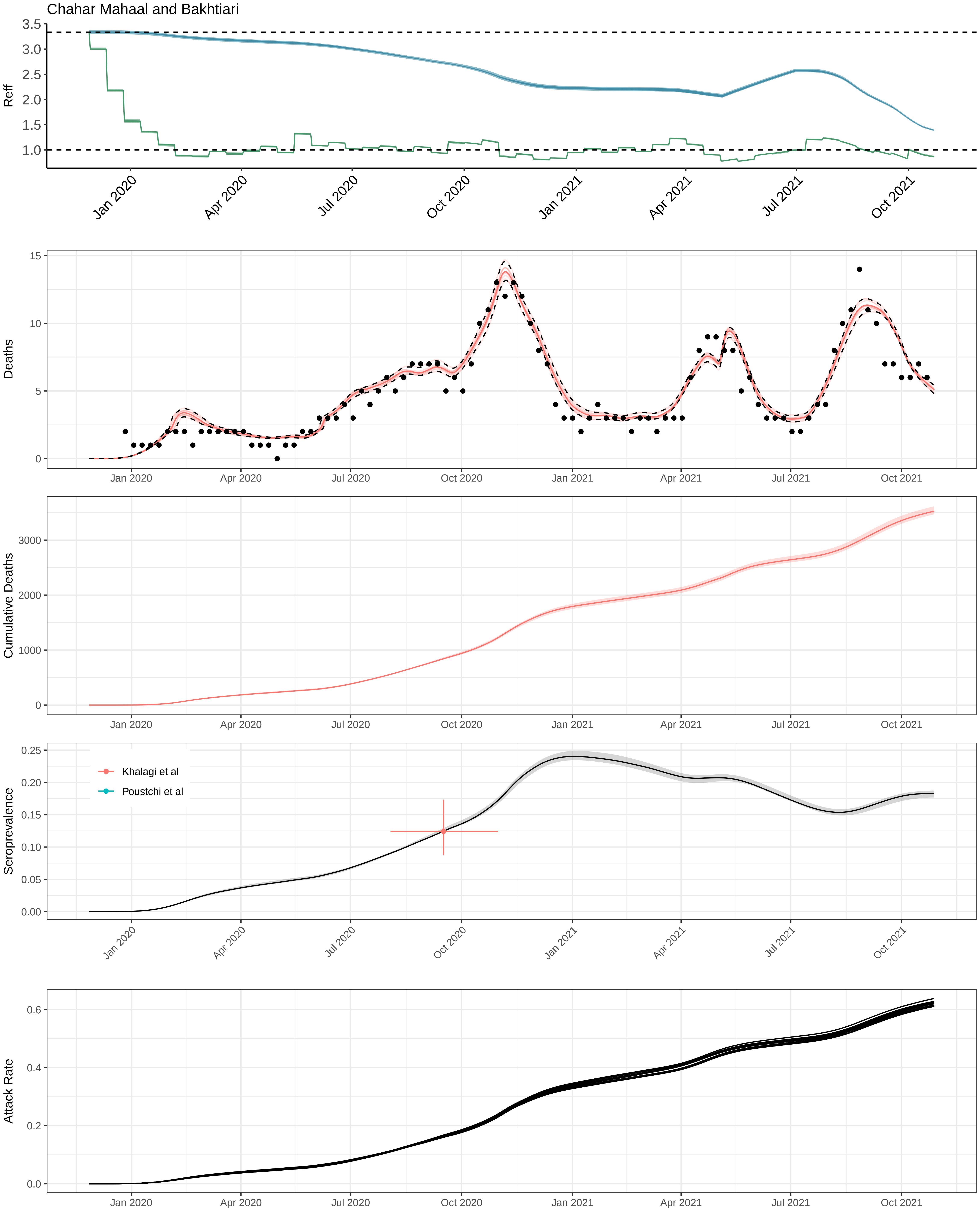
Chahar Mahaal and Bakhtiari
Reff
Deaths
Cumulative Deaths
Seroprevalence
Khalagi et al
Poustchi et al
Attack Rate
Jan 2020
Apr 2020
Jul 2020
Oct 2020
Jan 2021
Apr 2021
Jul 2021
Oct 2021

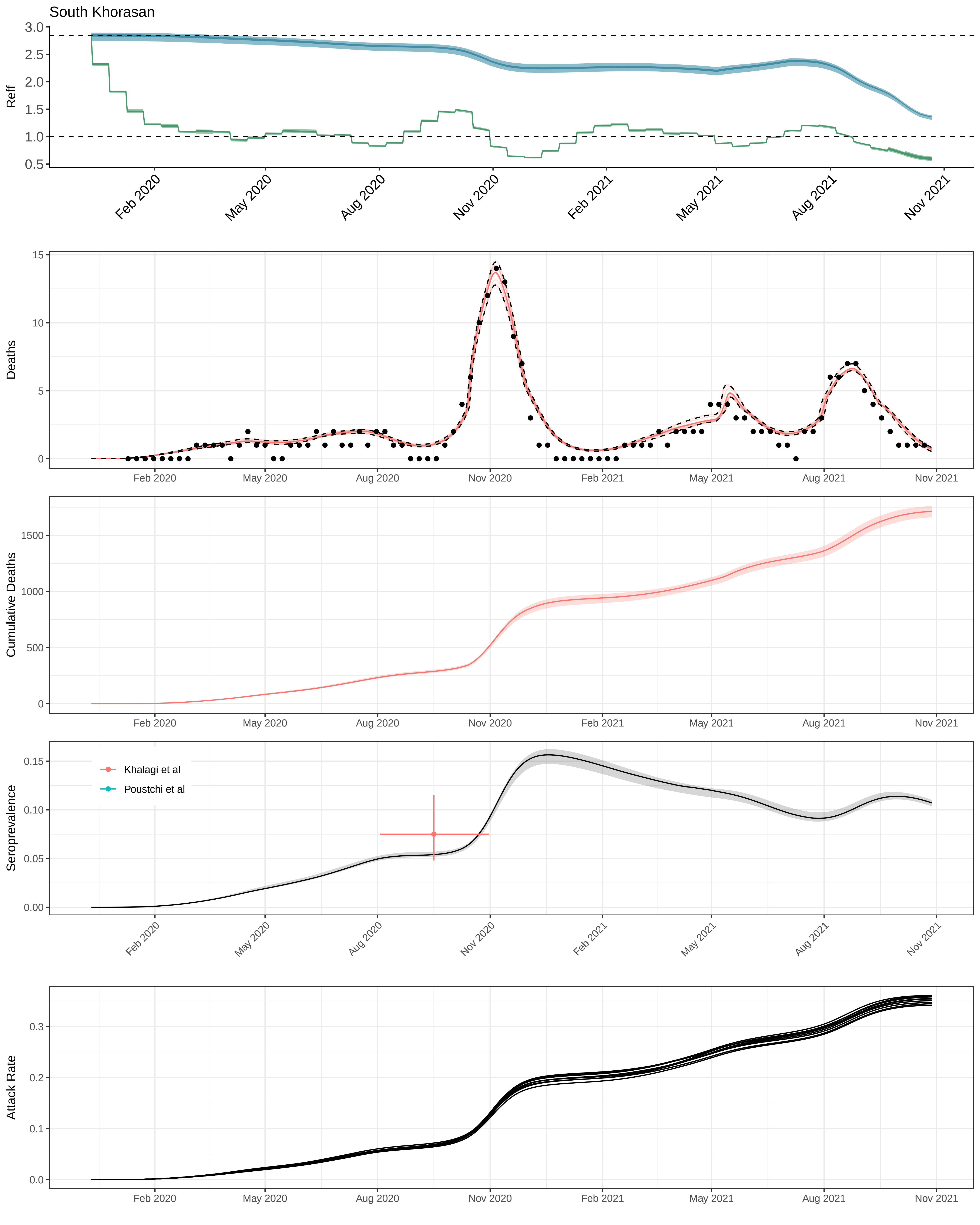

South Khorasan
Reff
3.0
2.5
2.0
1.5
1.0
0.5
Feb 2020
May 2020
Aug 2020
Nov 2020
Feb 2021
May 2021
Aug 2021
Nov 2021
Deaths
15
10
5
0
Cumulative Deaths
1500
1000
500
0
Seroprevalence
0.15
0.10
0.05
0.00
Khalagi et al
Poustchi et al
Attack Rate
0.3
0.2
0.1
0.0

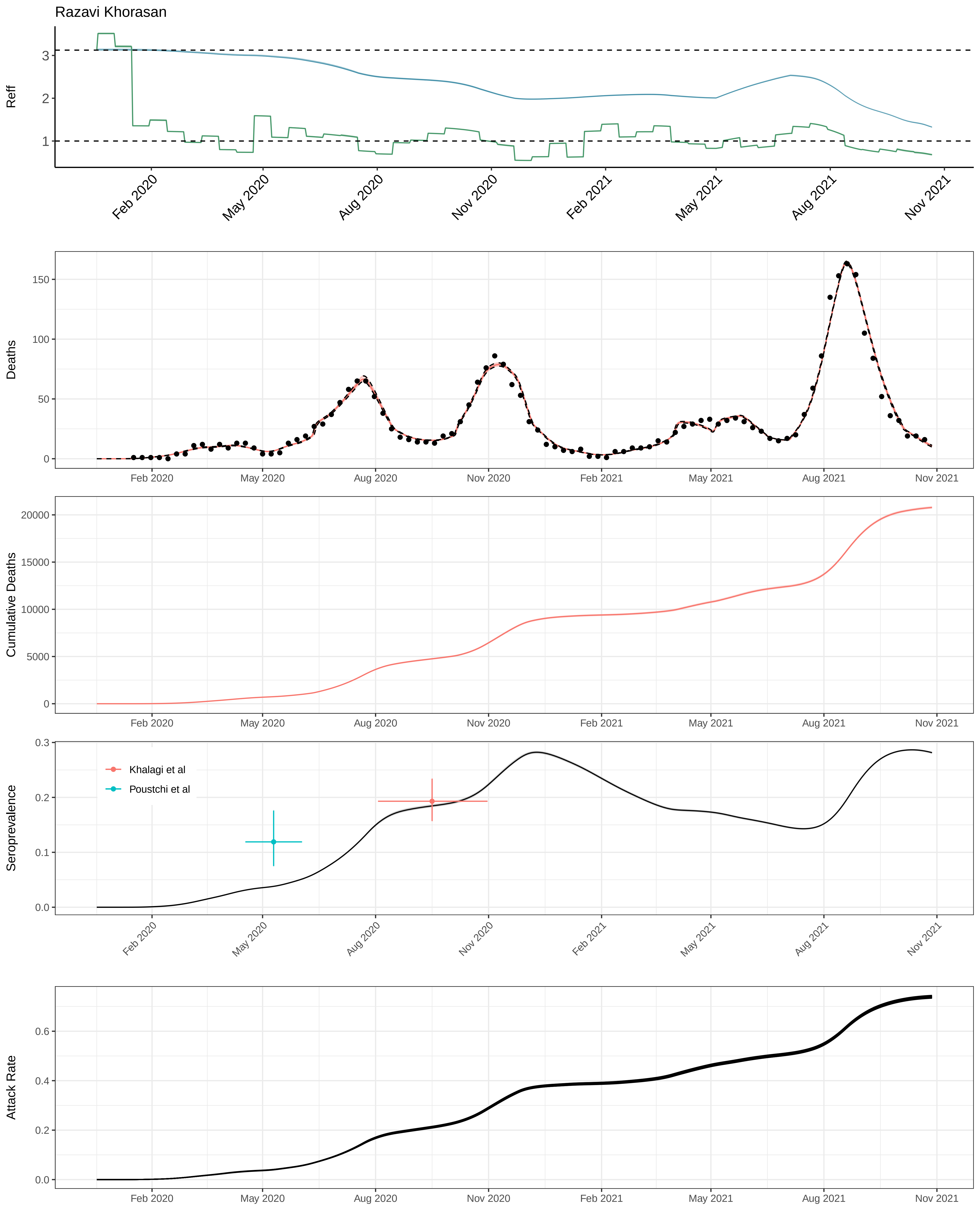

Razavi Khorasan
Reff
3
2
1
Feb 2020
May 2020
Aug 2020
Nov 2020
Feb 2021
May 2021
Aug 2021
Nov 2021
Deaths
150
100
50
0
Cumulative Deaths
20000
15000
10000
5000
0
Seroprevalence
0.3
0.2
0.1
0.0
Khalagi et al
Poustchi et al
Attack Rate
0.6
0.4
0.2
0.0

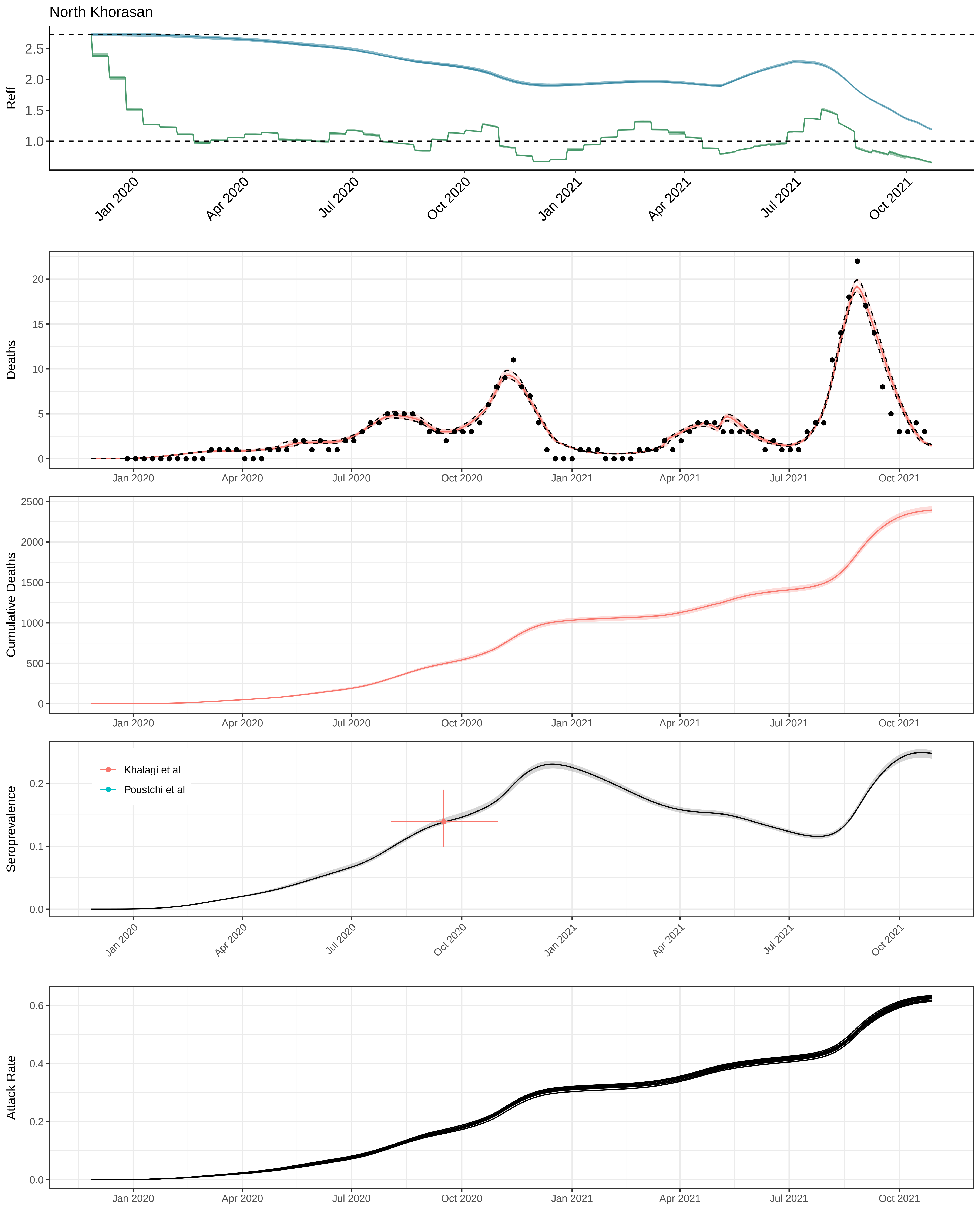

North Khorasan
Reff
Deaths
Cumulative Deaths
Seroprevalence
Khalagi et al
Poustchi et al
Attack Rate
Jan 2020
Apr 2020
Jul 2020
Oct 2020
Jan 2021
Apr 2021
Jul 2021
Oct 2021

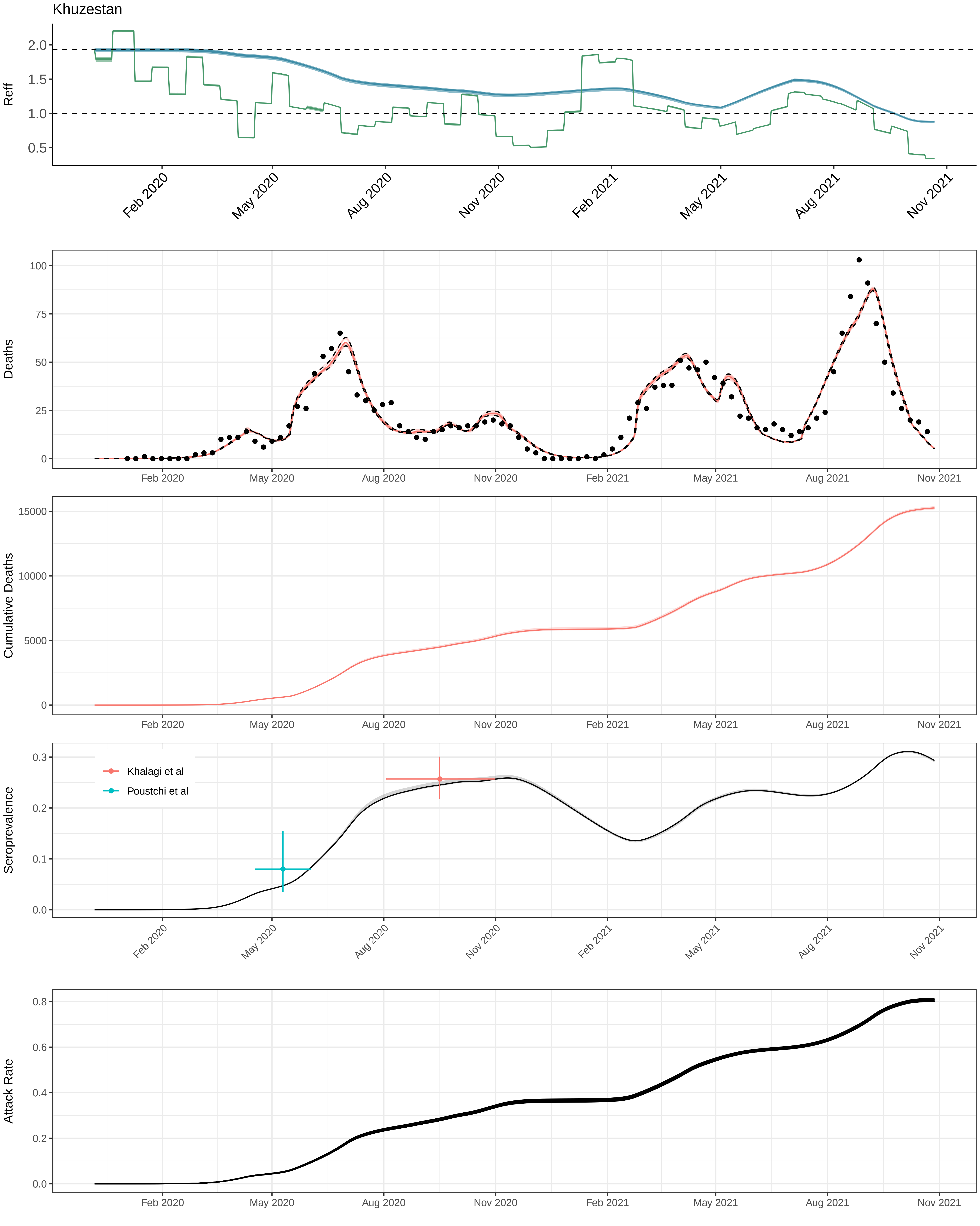
Khuzestan
Reff
2.0
1.5
1.0
0.5
Feb 2020
May 2020
Aug 2020
Nov 2020
Feb 2021
May 2021
Aug 2021
Nov 2021
Deaths
100
75
50
25
0
Cumulative Deaths
15000
10000
5000
0
Seroprevalence
0.3
0.2
0.1
0.0
Khalagi et al
Poustchi et al
Attack Rate
0.8
0.6
0.4
0.2
0.0

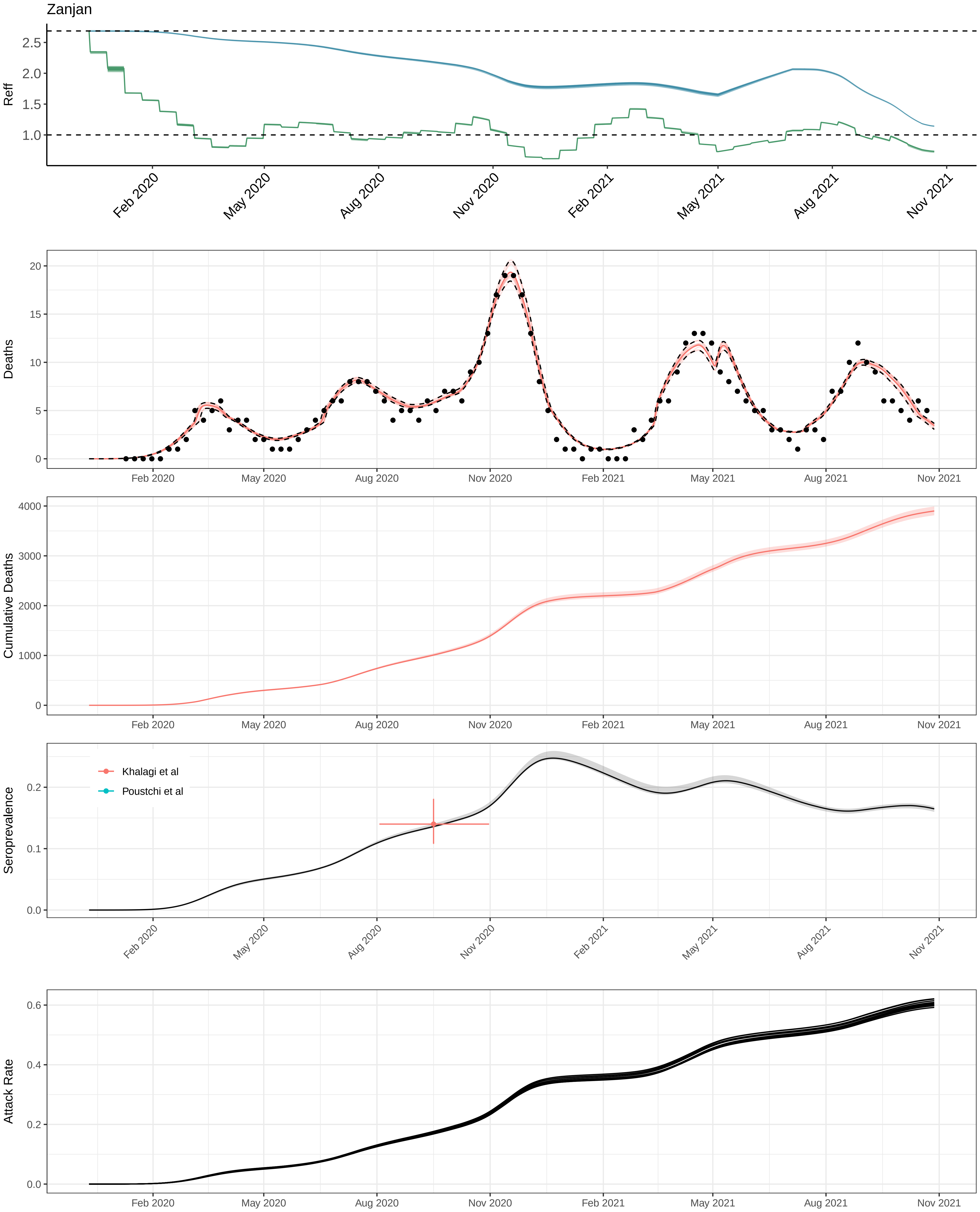

Zanjan
Reff
2.5
2.0
1.5
1.0
Feb 2020
May 2020
Aug 2020
Nov 2020
Feb 2021
May 2021
Aug 2021
Nov 2021
Deaths
20
15
10
5
0
Cumulative Deaths
4000
3000
2000
1000
0
Seroprevalence
Khalagi et al
Poustchi et al
0.2
0.1
0.0
Attack Rate
0.6
0.4
0.2
0.0

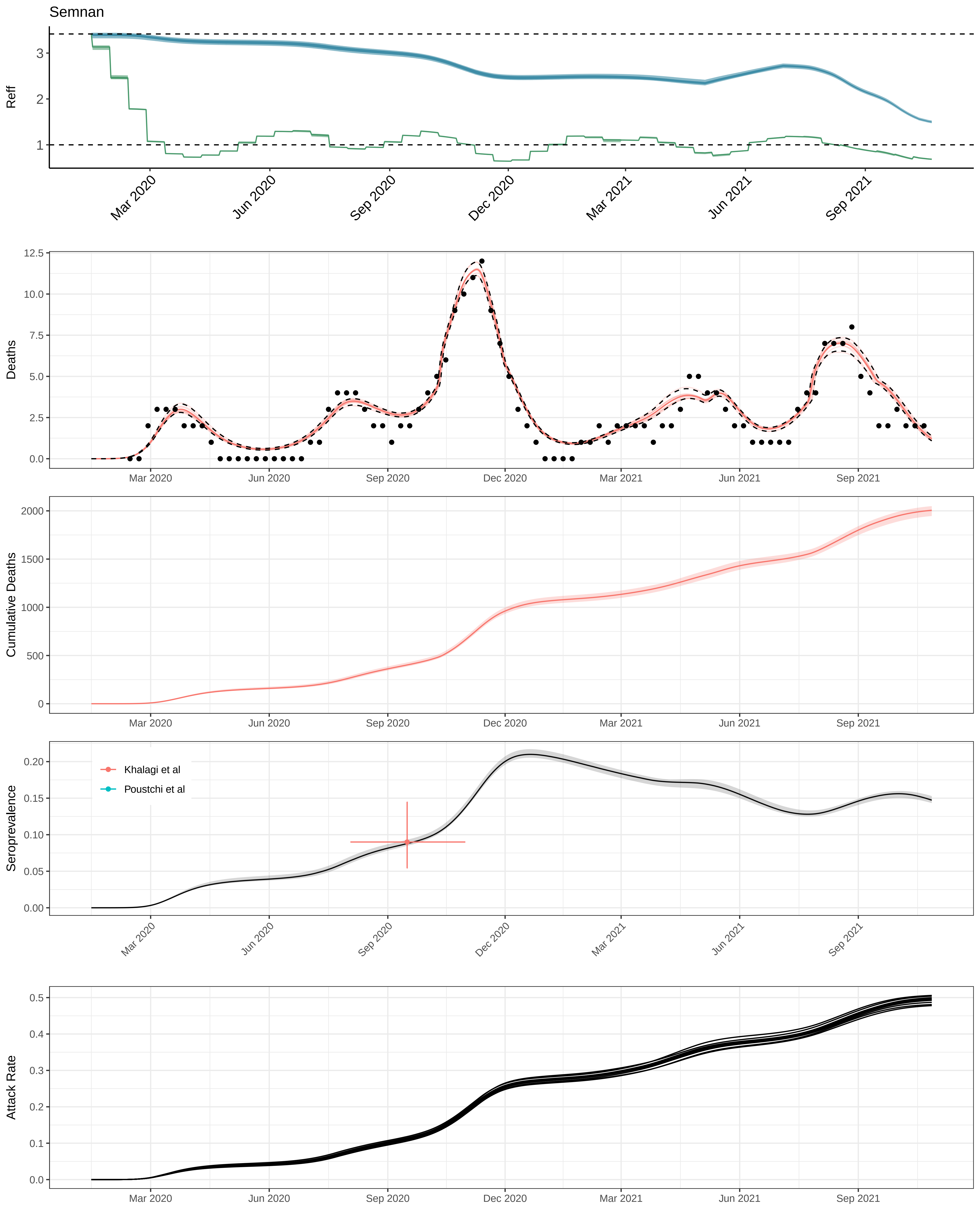

Semnan
Reff
Deaths
Cumulative Deaths
Seroprevalence
Attack Rate
Khalagi et al
Poustchi et al
Mar 2020
Jun 2020
Sep 2020
Dec 2020
Mar 2021
Jun 2021
Sep 2021

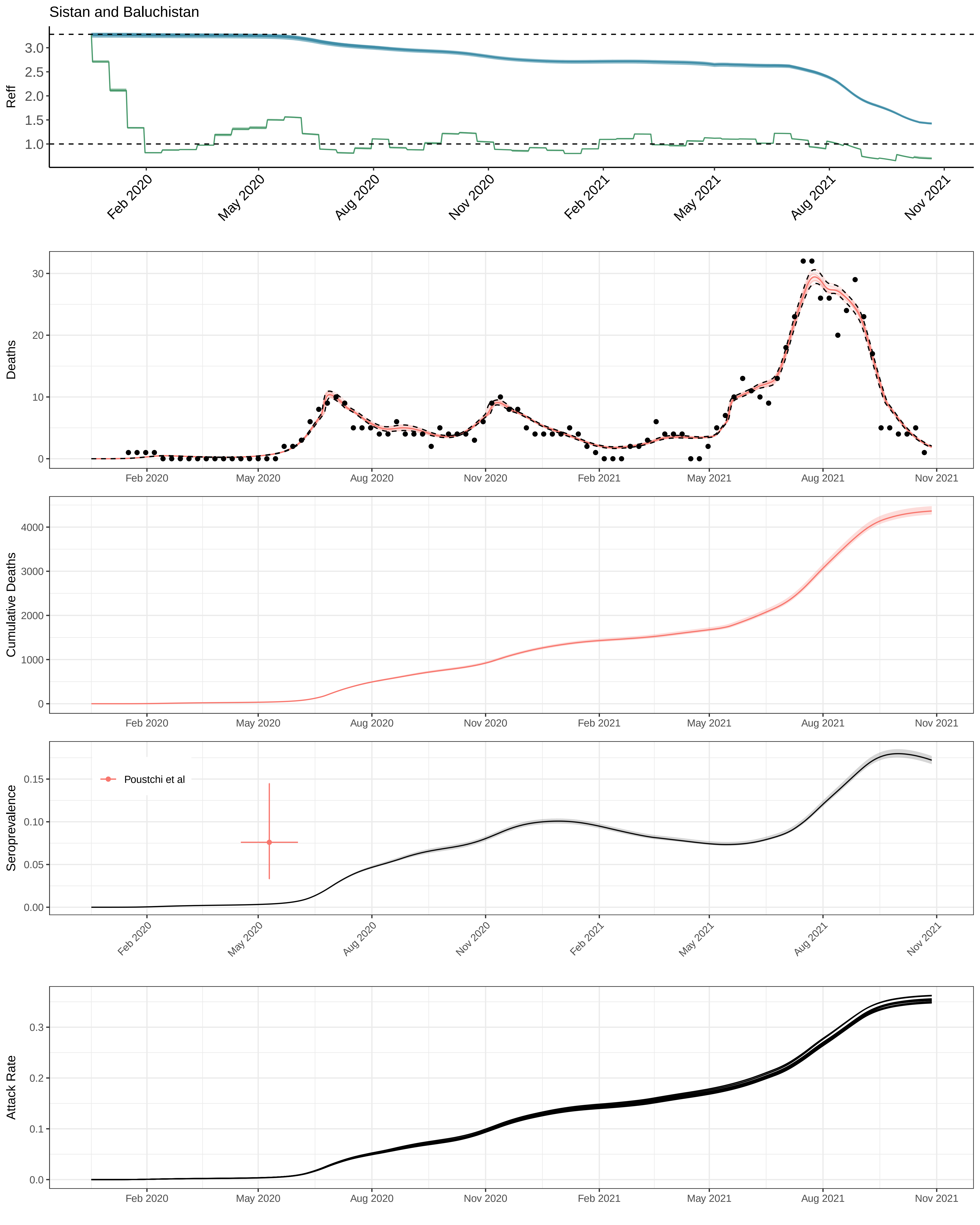
Sistan and Baluchistan
Reff
Deaths
Cumulative Deaths
Seroprevalence
Poustchi et al
Attack Rate
Feb 2020
May 2020
Aug 2020
Nov 2020
Feb 2021
May 2021
Aug 2021
Nov 2021

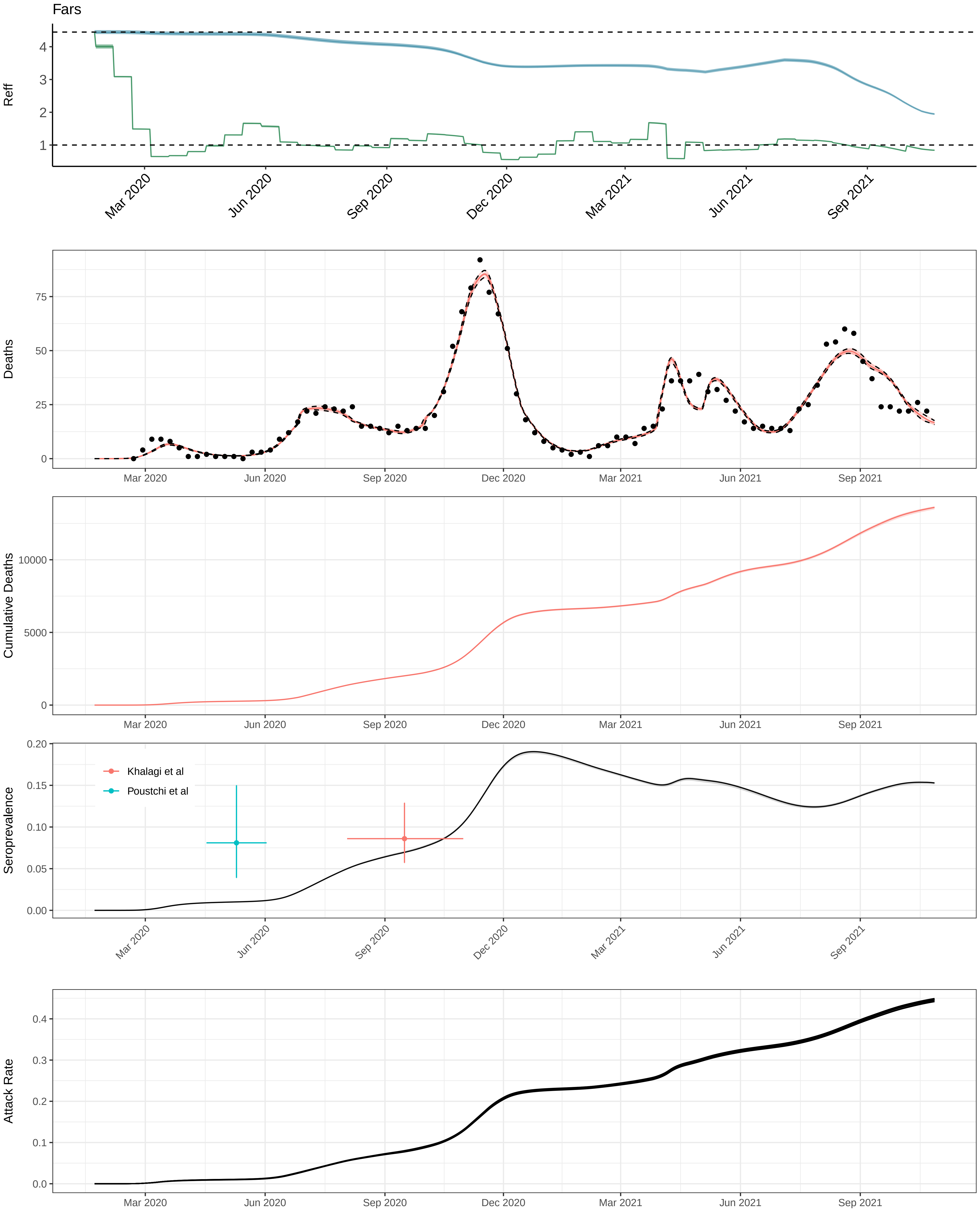
Fars
Reff
Deaths
Cumulative Deaths
Seroprevalence
Khalagi et al
Poustchi et al
Attack Rate
Mar 2020
Jun 2020
Sep 2020
Dec 2020
Mar 2021
Jun 2021
Sep 2021

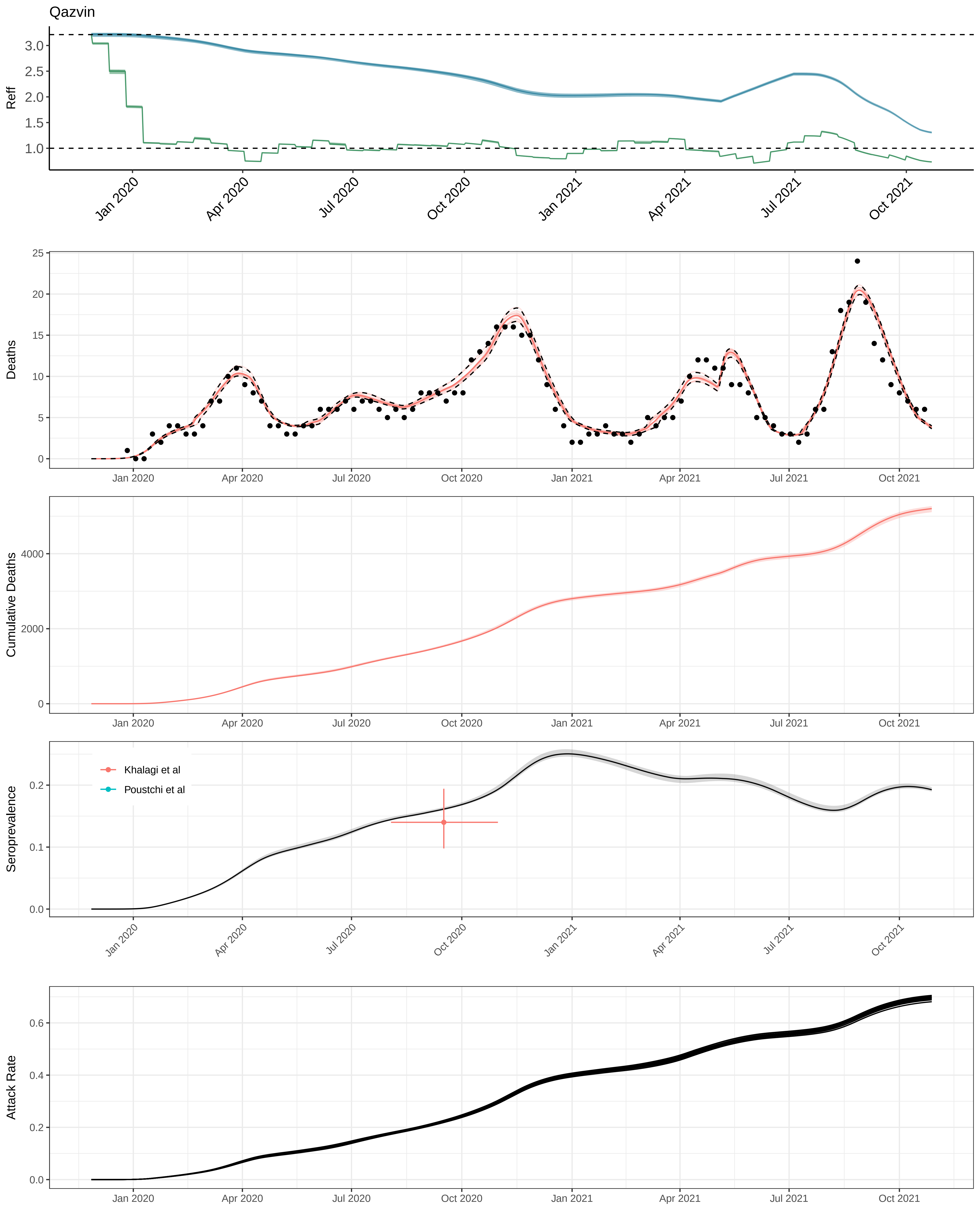
Qazvin
Reff
3.0
2.5
2.0
1.5
1.0
Jan 2020
Apr 2020
Jul 2020
Oct 2020
Jan 2021
Apr 2021
Jul 2021
Oct 2021
Deaths
25
20
15
10
5
0
Cumulative Deaths
4000
2000
0
Seroprevalence
0.2
0.1
0.0
Khalagi et al
Poustchi et al
Attack Rate
0.6
0.4
0.2
0.0

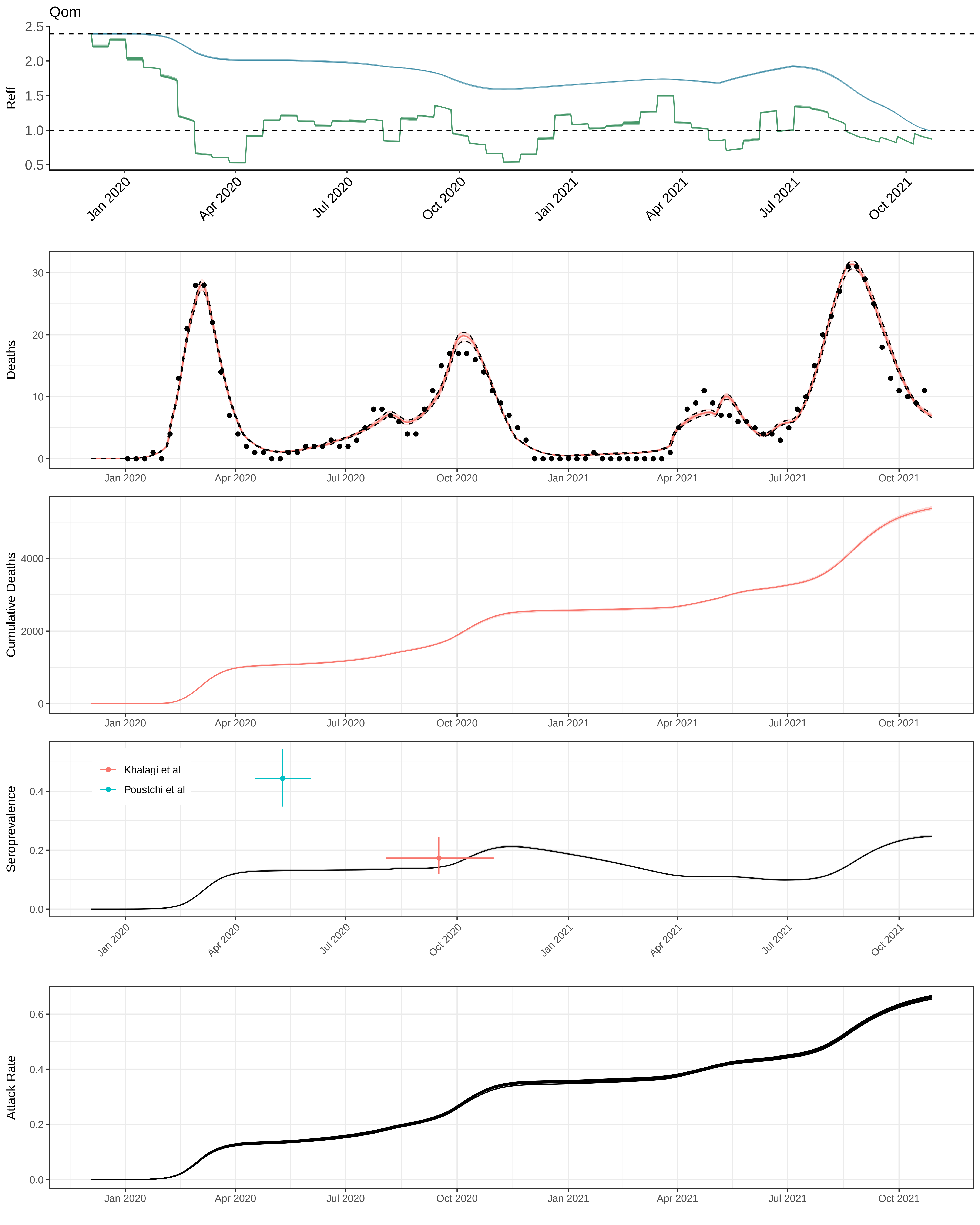
Qom
Reff
Deaths
Cumulative Deaths
Seroprevalence
Khalagi et al
Poustchi et al
Attack Rate
Jan 2020
Apr 2020
Jul 2020
Oct 2020
Jan 2021
Apr 2021
Jul 2021
Oct 2021

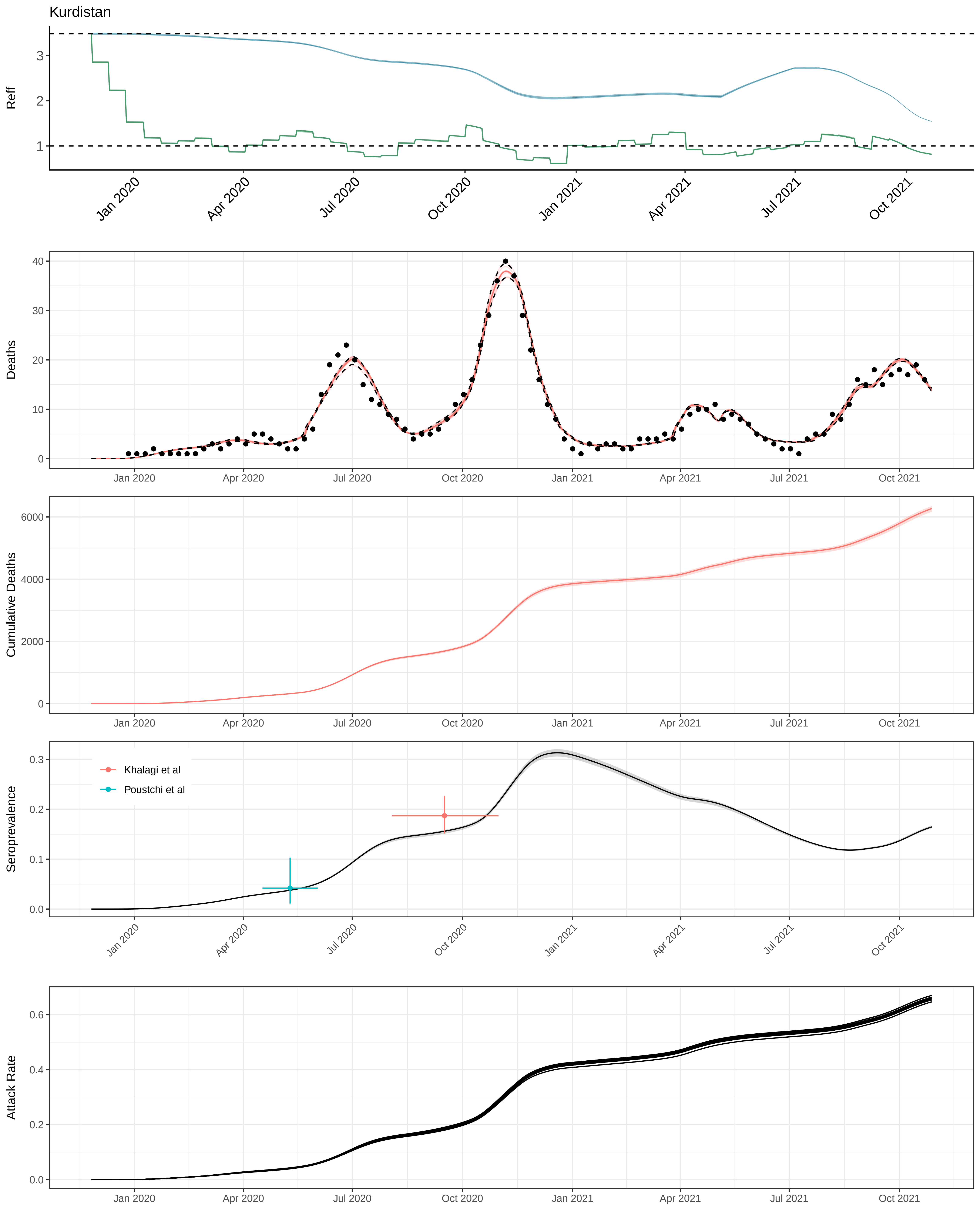

Kurdistan
Reff
Deaths
Cumulative Deaths
Seroprevalence
Khalagi et al
Poustchi et al
Attack Rate
Jan 2020
Apr 2020
Jul 2020
Oct 2020
Jan 2021
Apr 2021
Jul 2021
Oct 2021

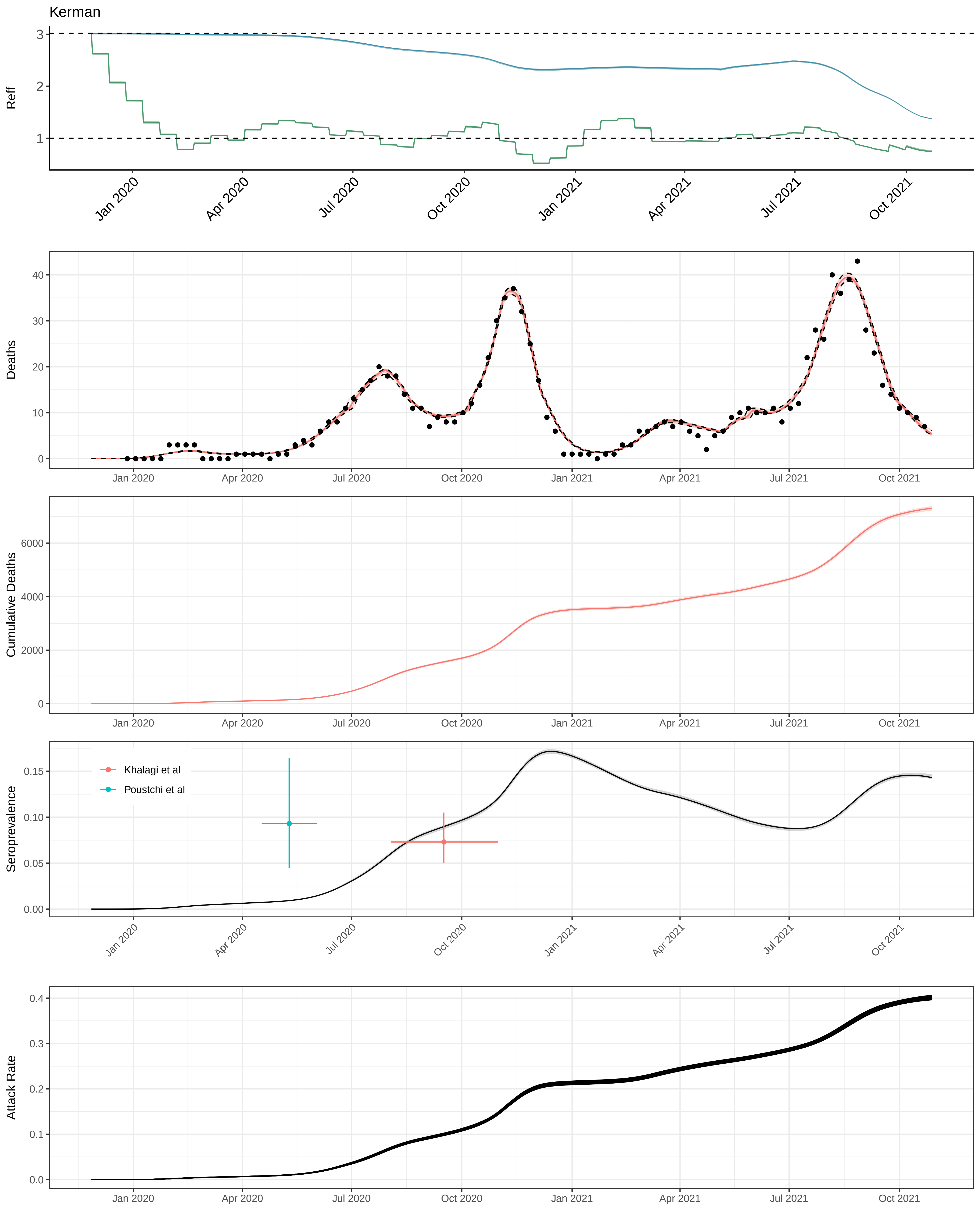

Kerman
Reff
Deaths
Cumulative Deaths
Seroprevalence
Khalagi et al
Poustchi et al
Attack Rate
Jan 2020
Apr 2020
Jul 2020
Oct 2020
Jan 2021
Apr 2021
Jul 2021
Oct 2021

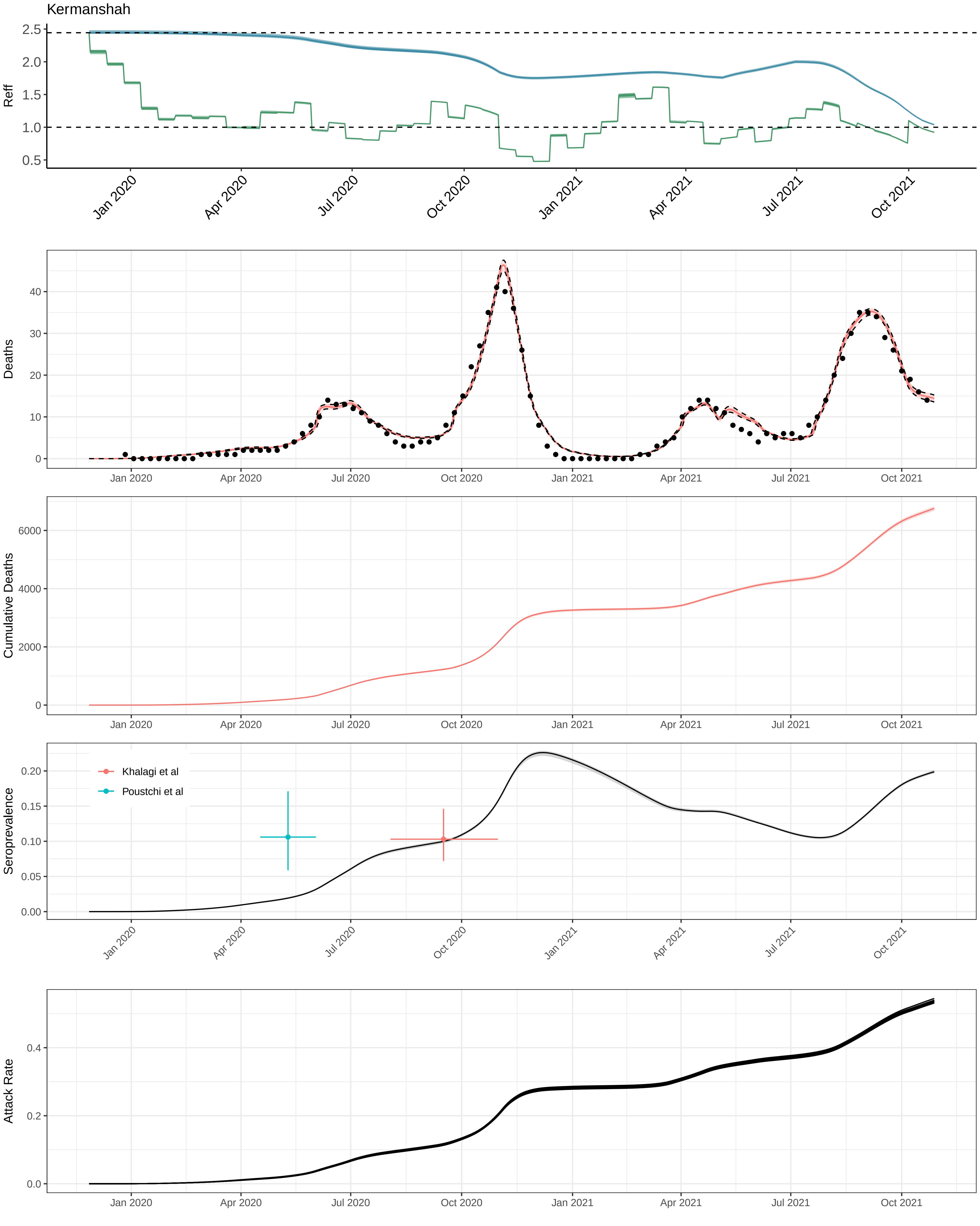

Kermanshah
Reff
Deaths
Cumulative Deaths
Seroprevalence
Khalagi et al
Poustchi et al
Attack Rate
Jan 2020
Apr 2020
Jul 2020
Oct 2020
Jan 2021
Apr 2021
Jul 2021
Oct 2021

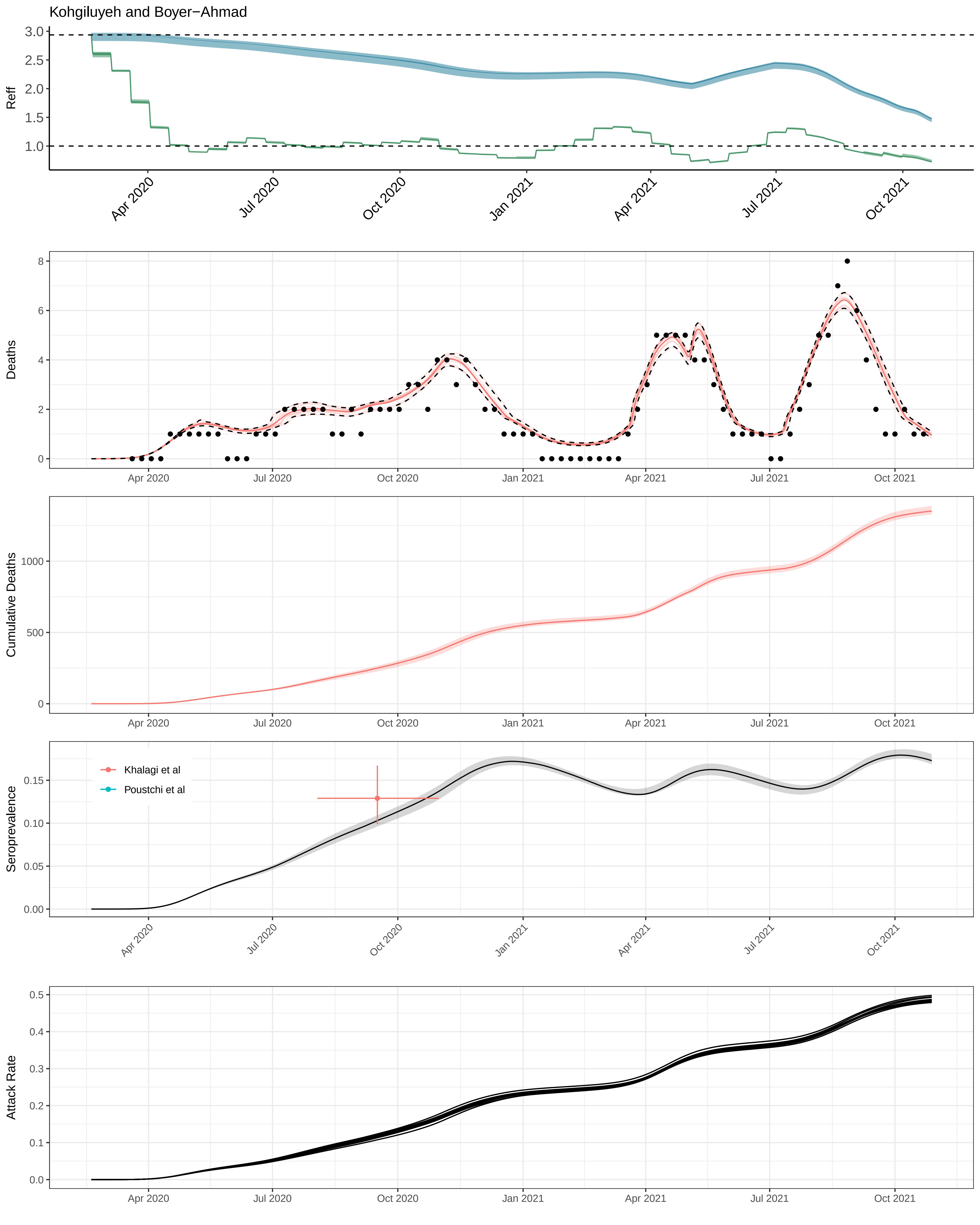

Kohgiluyeh and Boyer−Ahmad
Reff
Deaths
Cumulative Deaths
Seroprevalence
Khalagi et al
Poustchi et al
Attack Rate
Apr 2020
Jul 2020
Oct 2020
Jan 2021
Apr 2021
Jul 2021
Oct 2021

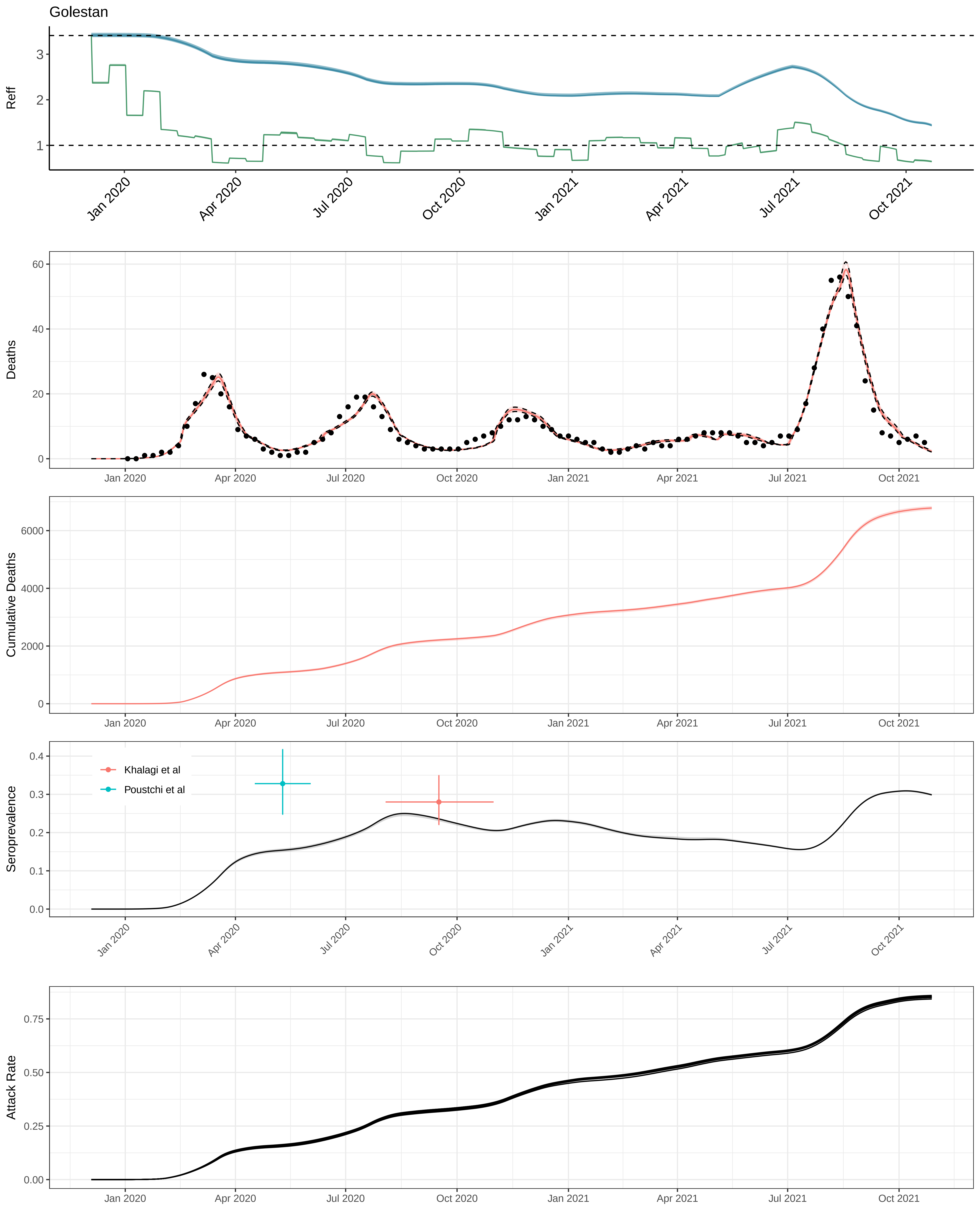

Golestan
Reff
Deaths
Cumulative Deaths
Seroprevalence
Khalagi et al
Poustchi et al
Attack Rate
Jan 2020
Apr 2020
Jul 2020
Oct 2020
Jan 2021
Apr 2021
Jul 2021
Oct 2021

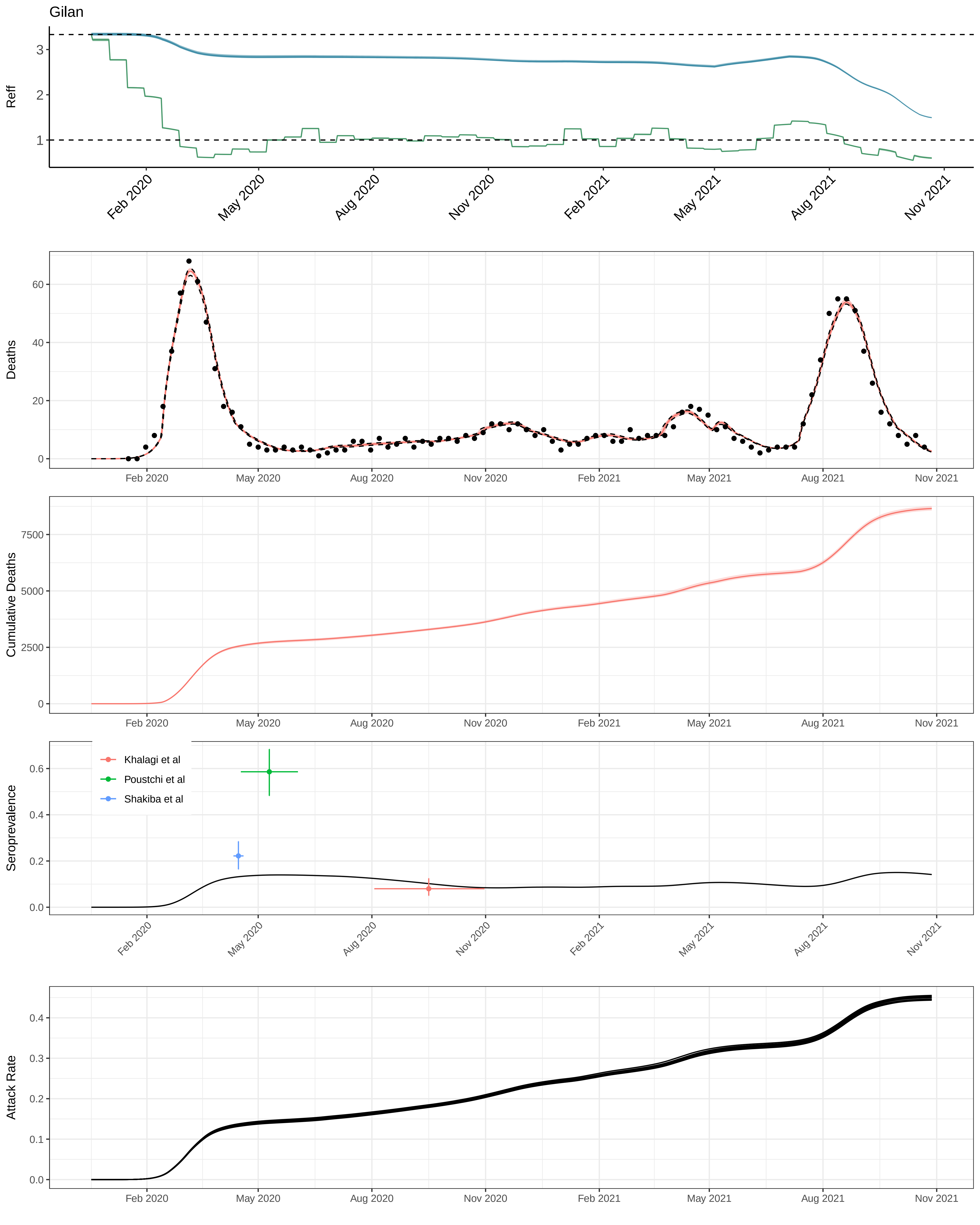

Gilan
Reff
Deaths
Cumulative Deaths
Seroprevalence
Attack Rate
Khalagi et al
Poustchi et al
Shakiba et al
Feb 2020
May 2020
Aug 2020
Nov 2020
Feb 2021
May 2021
Aug 2021
Nov 2021

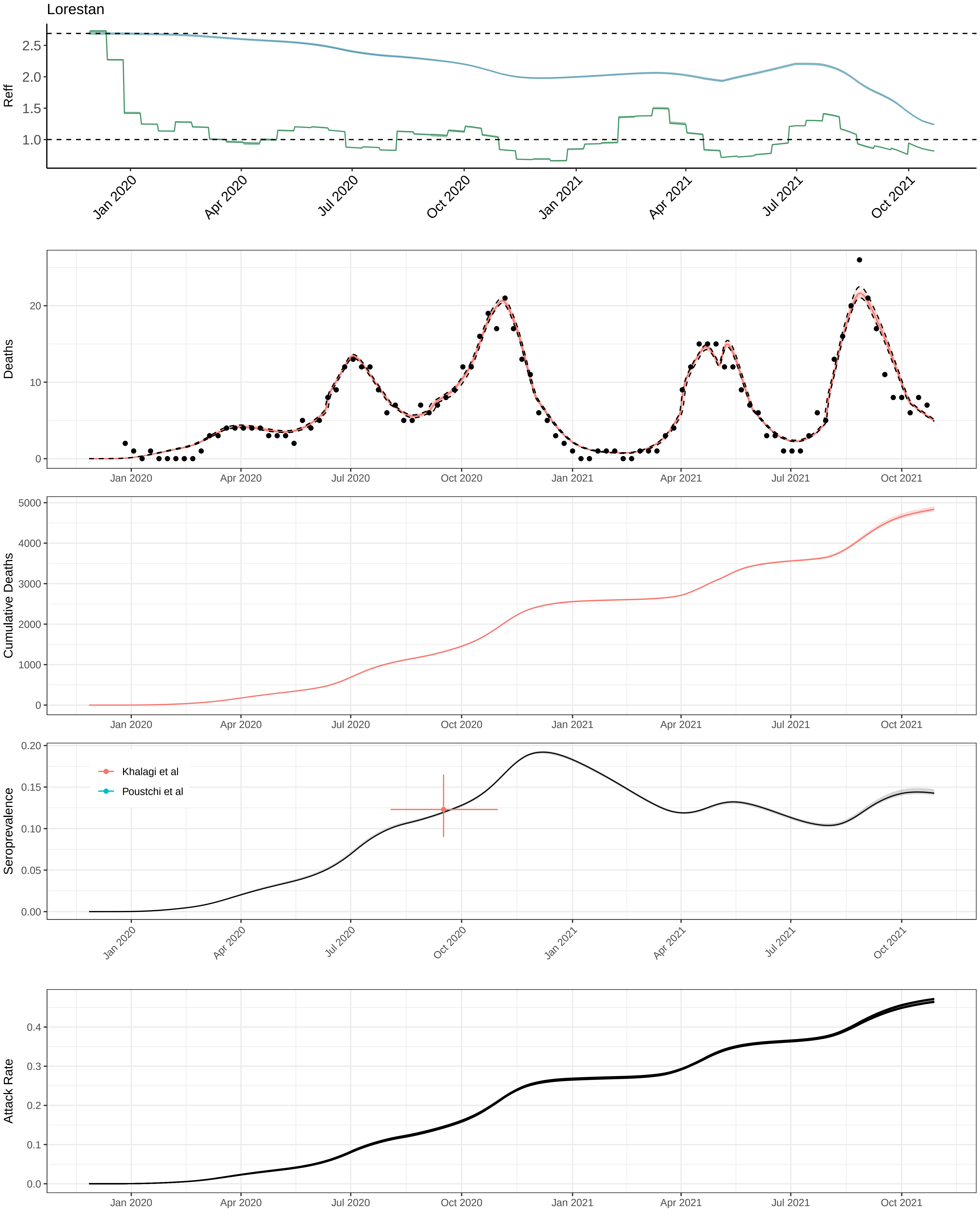
Lorestan
Reff
2.5
2.0
1.5
1.0
Jan 2020
Apr 2020
Jul 2020
Oct 2020
Jan 2021
Apr 2021
Jul 2021
Oct 2021
Deaths
20
10
0
Cumulative Deaths
5000
4000
3000
2000
1000
0
Seroprevalence
0.20
0.15
0.10
0.05
0.00
Khalagi et al
Poustchi et al
Attack Rate
0.4
0.3
0.2
0.1
0.0

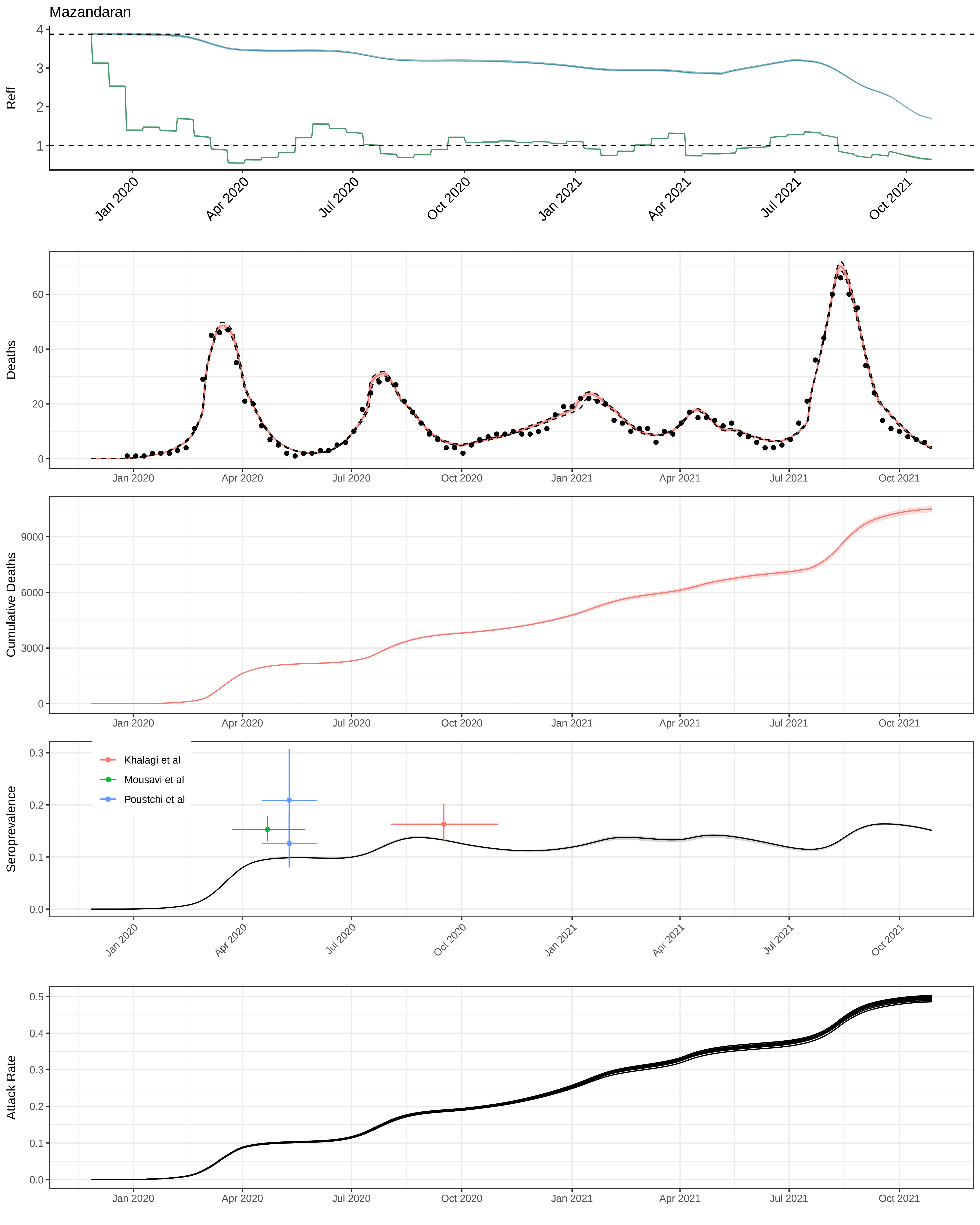

Mazandaran
Reff
Deaths
Cumulative Deaths
Seroprevalence
Khalagi et al
Mousavi et al
Poustchi et al
Attack Rate
Jan 2020
Apr 2020
Jul 2020
Oct 2020
Jan 2021
Apr 2021
Jul 2021
Oct 2021

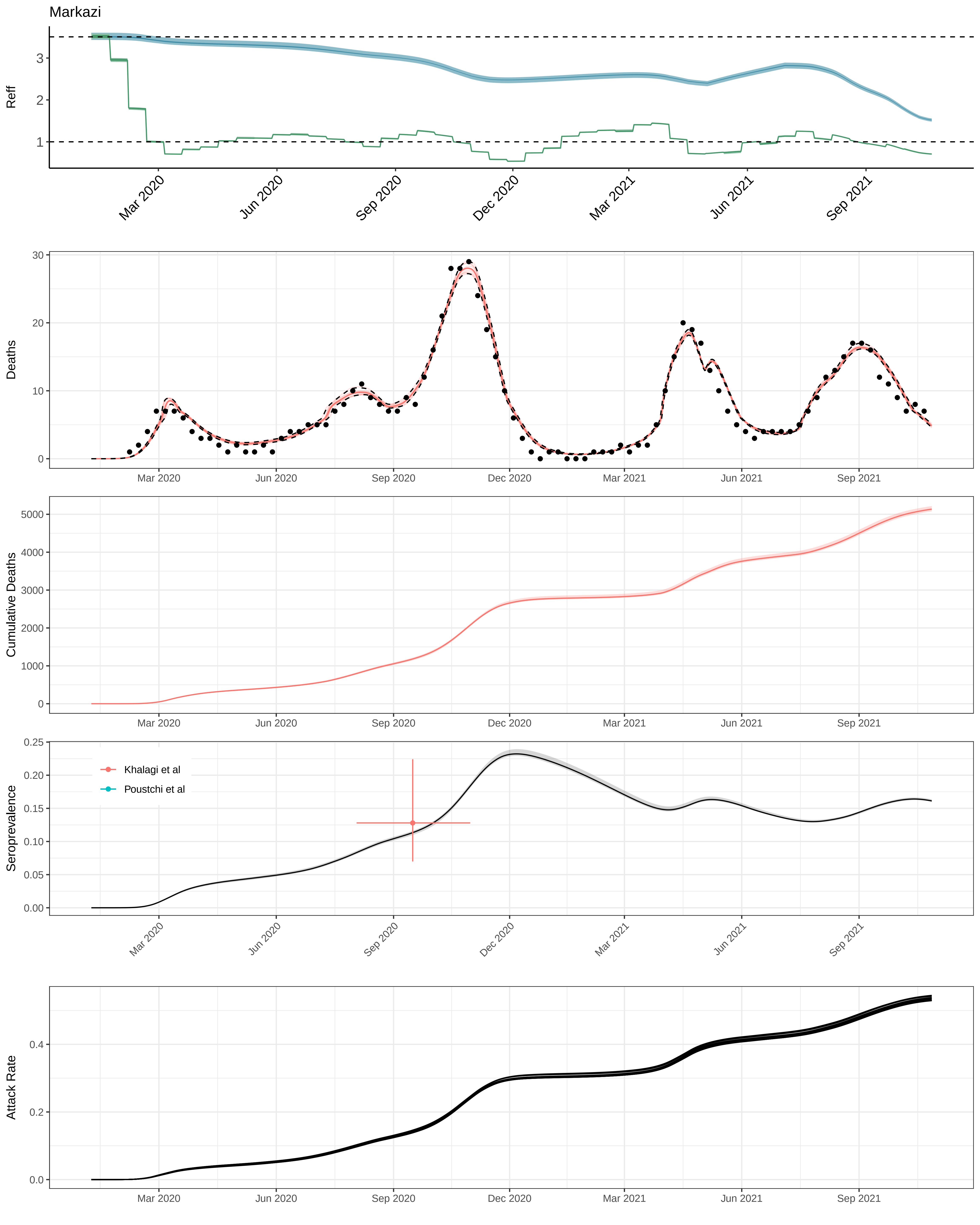

Markazi
Reff
Deaths
Cumulative Deaths
Seroprevalence
Khalagi et al
Poustchi et al
Attack Rate
Mar 2020
Jun 2020
Sep 2020
Dec 2020
Mar 2021
Jun 2021
Sep 2021

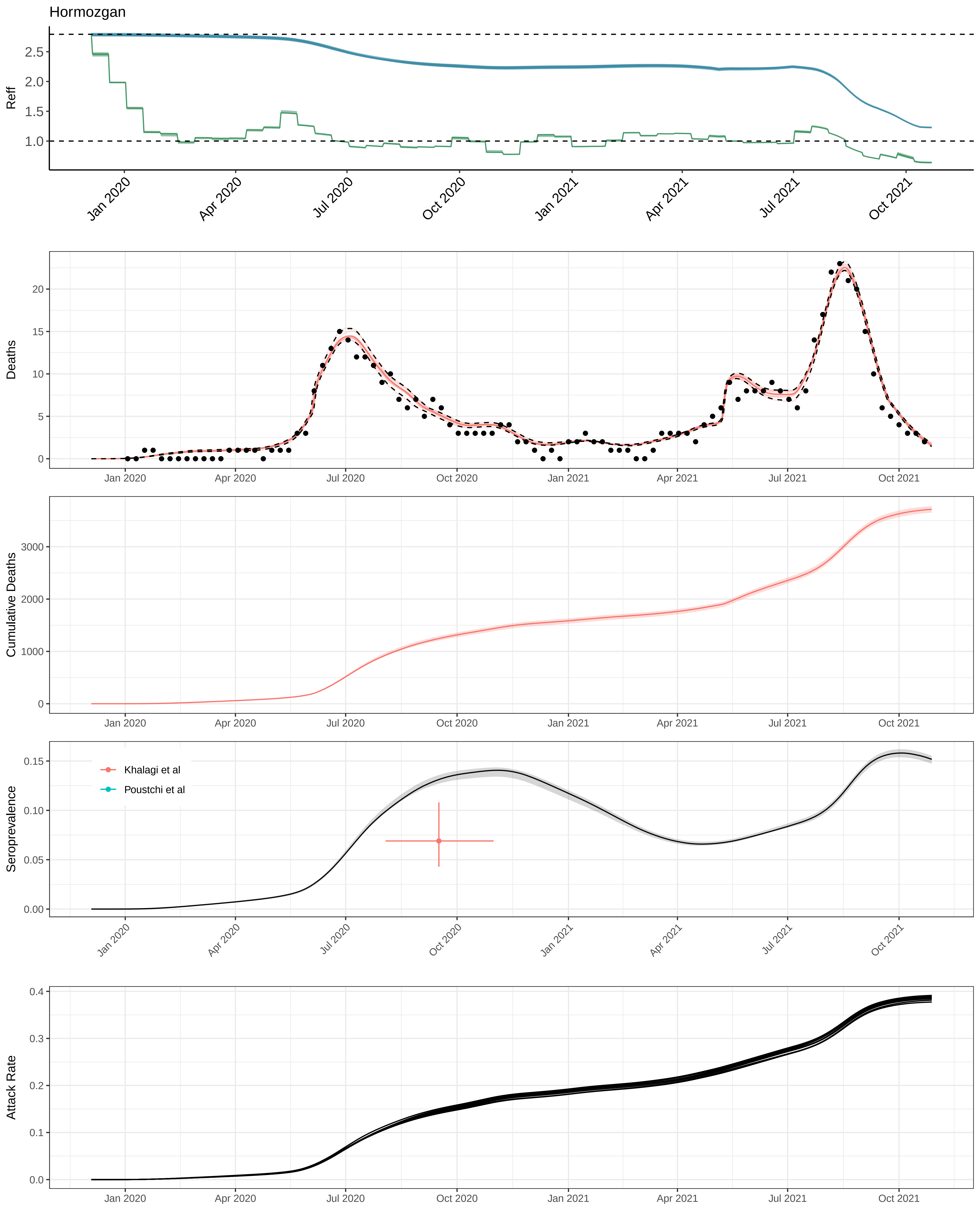
Hormozgan
Reff
2.5
2.0
1.5
1.0
Jan 2020
Apr 2020
Jul 2020
Oct 2020
Jan 2021
Apr 2021
Jul 2021
Oct 2021
Deaths
20
15
10
5
0
Cumulative Deaths
3000
2000
1000
0
Seroprevalence
0.15
0.10
0.05
0.00
Khalagi et al
Poustchi et al
Attack Rate
0.4
0.3
0.2
0.1
0.0

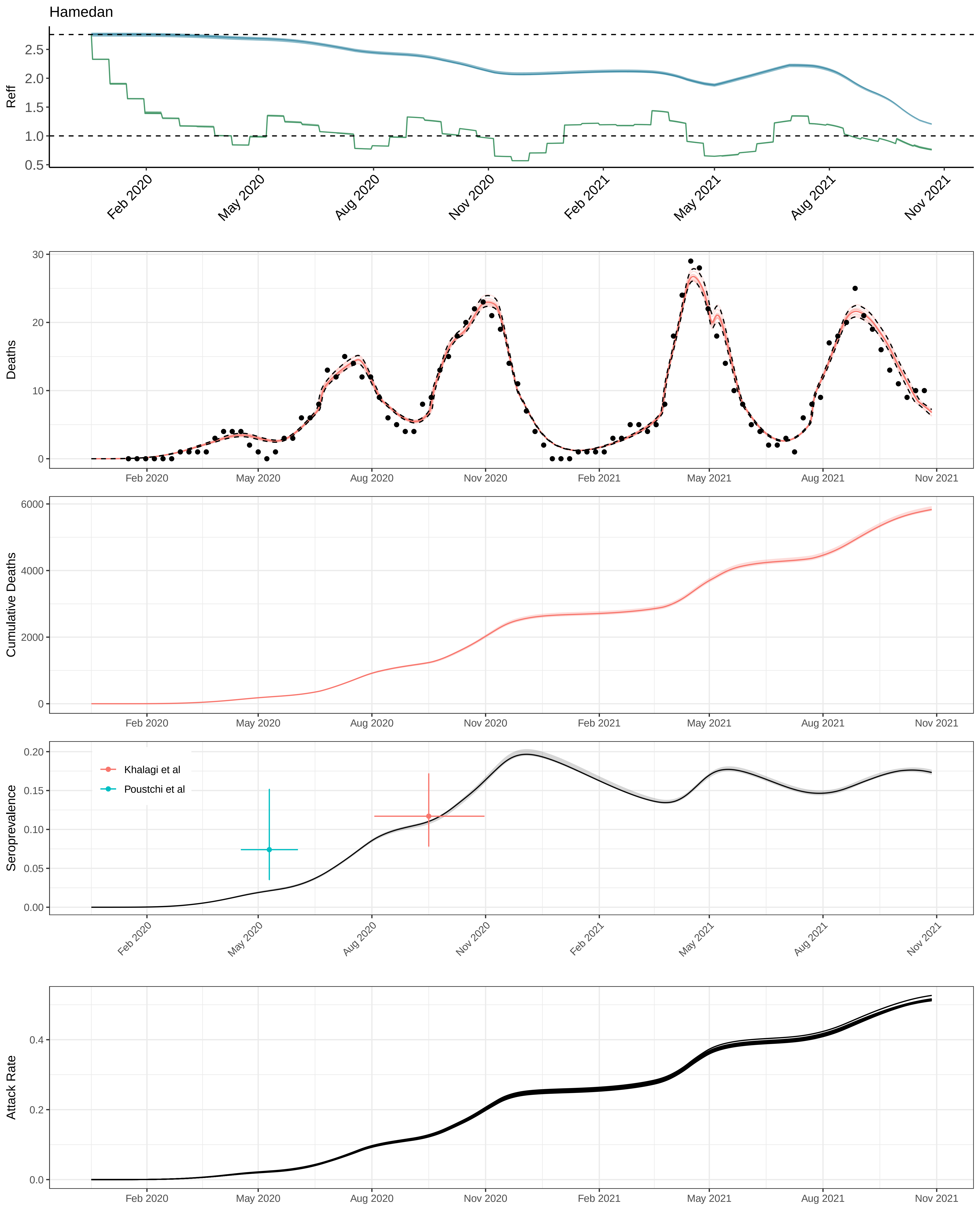

Hamedan
Reff
Deaths
Cumulative Deaths
Seroprevalence
Khalagi et al
Poustchi et al
Attack Rate
Feb 2020
May 2020
Aug 2020
Nov 2020
Feb 2021
May 2021
Aug 2021
Nov 2021

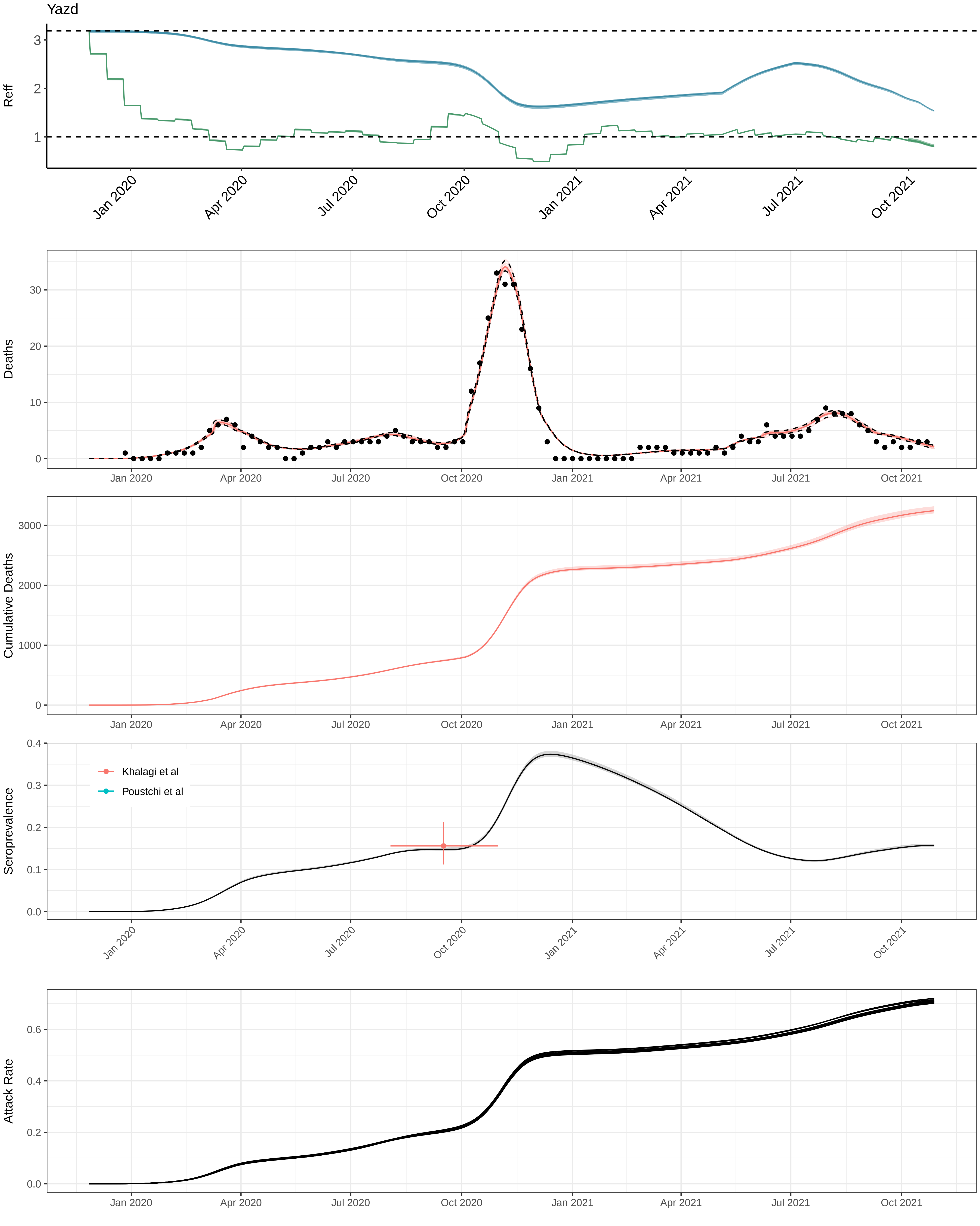
Yazd
Reff
Deaths
Cumulative Deaths
Seroprevalence
Khalagi et al
Poustchi et al
Attack Rate
Jan 2020
Apr 2020
Jul 2020
Oct 2020
Jan 2021
Apr 2021
Jul 2021
Oct 2021
