## Supplementary data S1 to S6 for "A framework for reconstructing SARS-CoV-2 transmission dynamics using excess mortality data": fitting_vacc_orderly_bundles_derived_vaccine_new_odriscoll_best.pdf

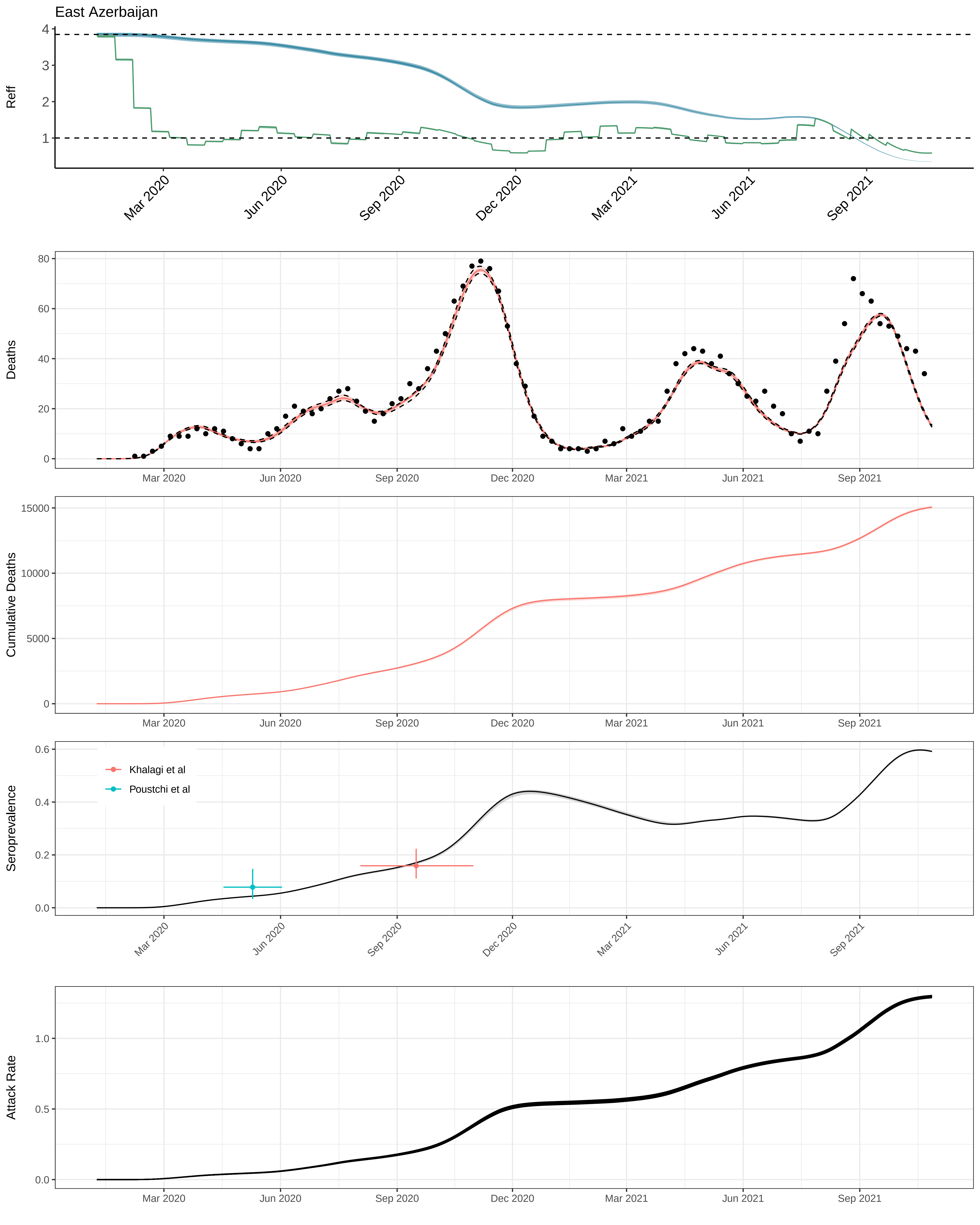
East Azerbaijan
Reff
4
3
2
1
Mar 2020
Jun 2020
Sep 2020
Dec 2020
Mar 2021
Jun 2021
Sep 2021
Deaths
80
60
40
20
0
Cumulative Deaths
15000
10000
5000
0
Seroprevalence
0.6
0.4
0.2
0.0
Khalagi et al
Poustchi et al
Attack Rate
1.0
0.5
0.0

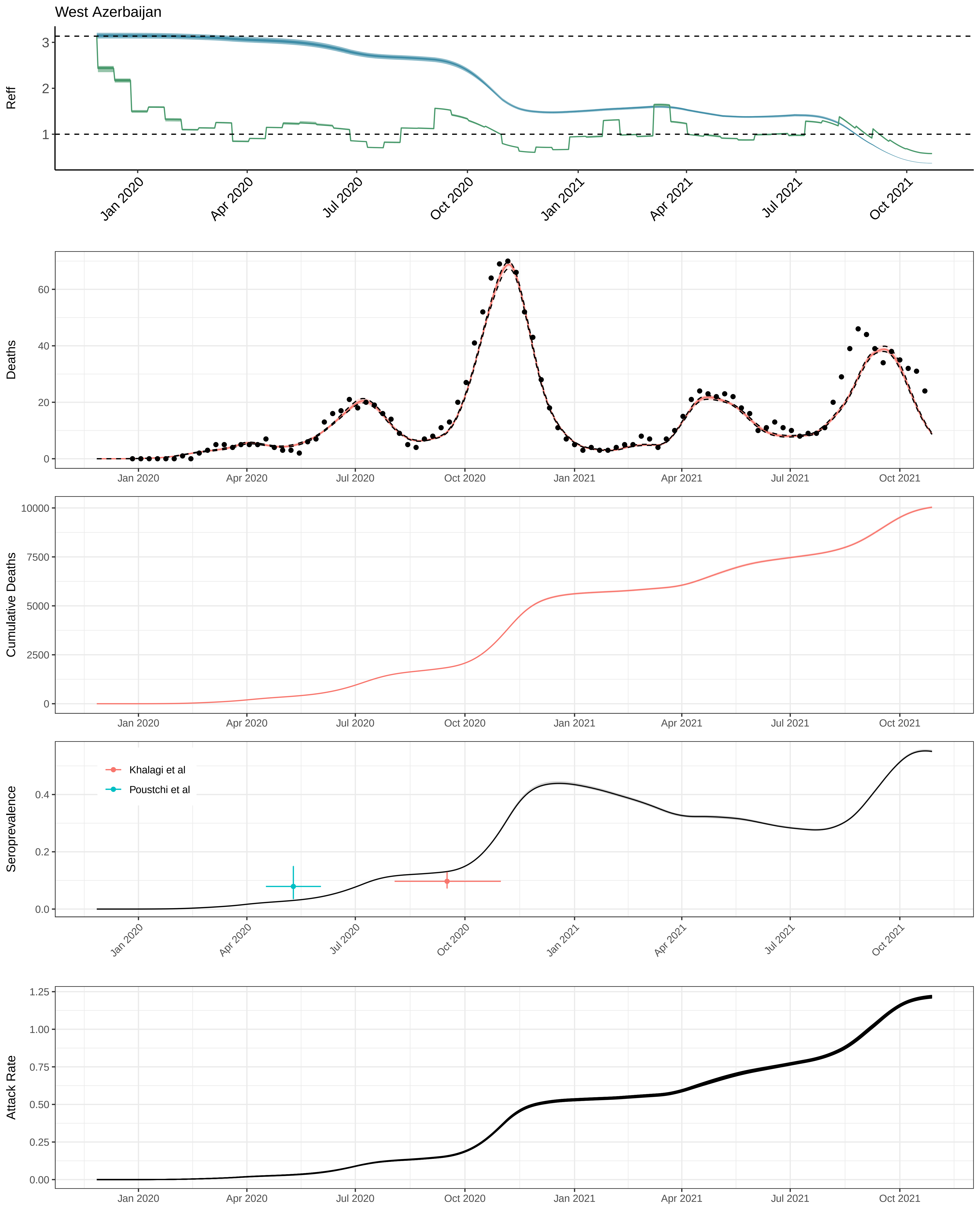
West Azerbaijan
Reff
Deaths
Cumulative Deaths
Seroprevalence
Khalagi et al
Poustchi et al
Attack Rate
Jan 2020
Apr 2020
Jul 2020
Oct 2020
Jan 2021
Apr 2021
Jul 2021
Oct 2021

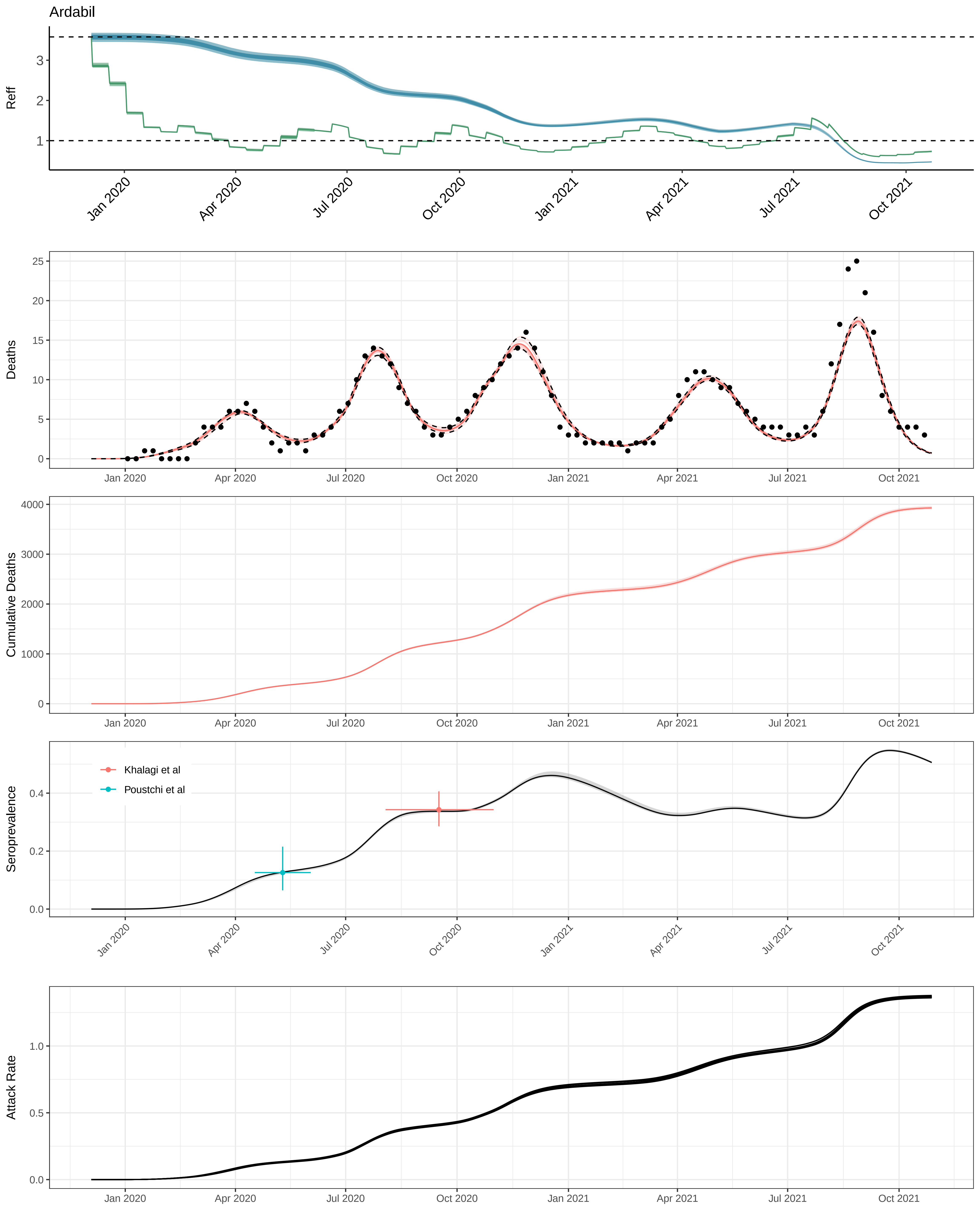

Ardabil
Reff
Deaths
Cumulative Deaths
Seroprevalence
Attack Rate
Khalagi et al
Poustchi et al
Jan 2020
Apr 2020
Jul 2020
Oct 2020
Jan 2021
Apr 2021
Jul 2021
Oct 2021

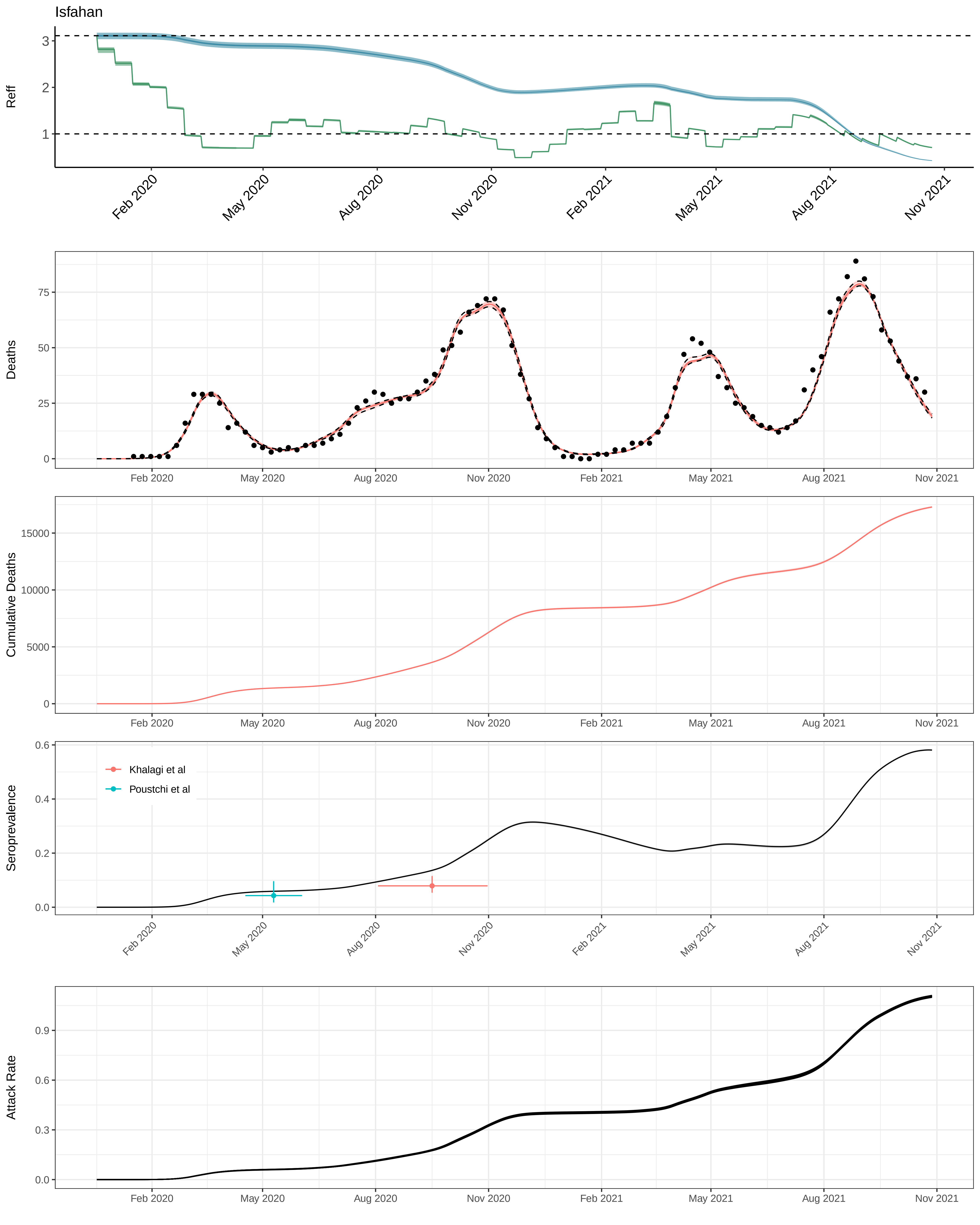

Isfahan
Reff
3
2
1
Feb 2020
May 2020
Aug 2020
Nov 2020
Feb 2021
May 2021
Aug 2021
Nov 2021
Deaths
75
50
25
0
Cumulative Deaths
15000
10000
5000
0
Seroprevalence
0.6
0.4
0.2
0.0
Khalagi et al
Poustchi et al
Attack Rate
0.9
0.6
0.3
0.0

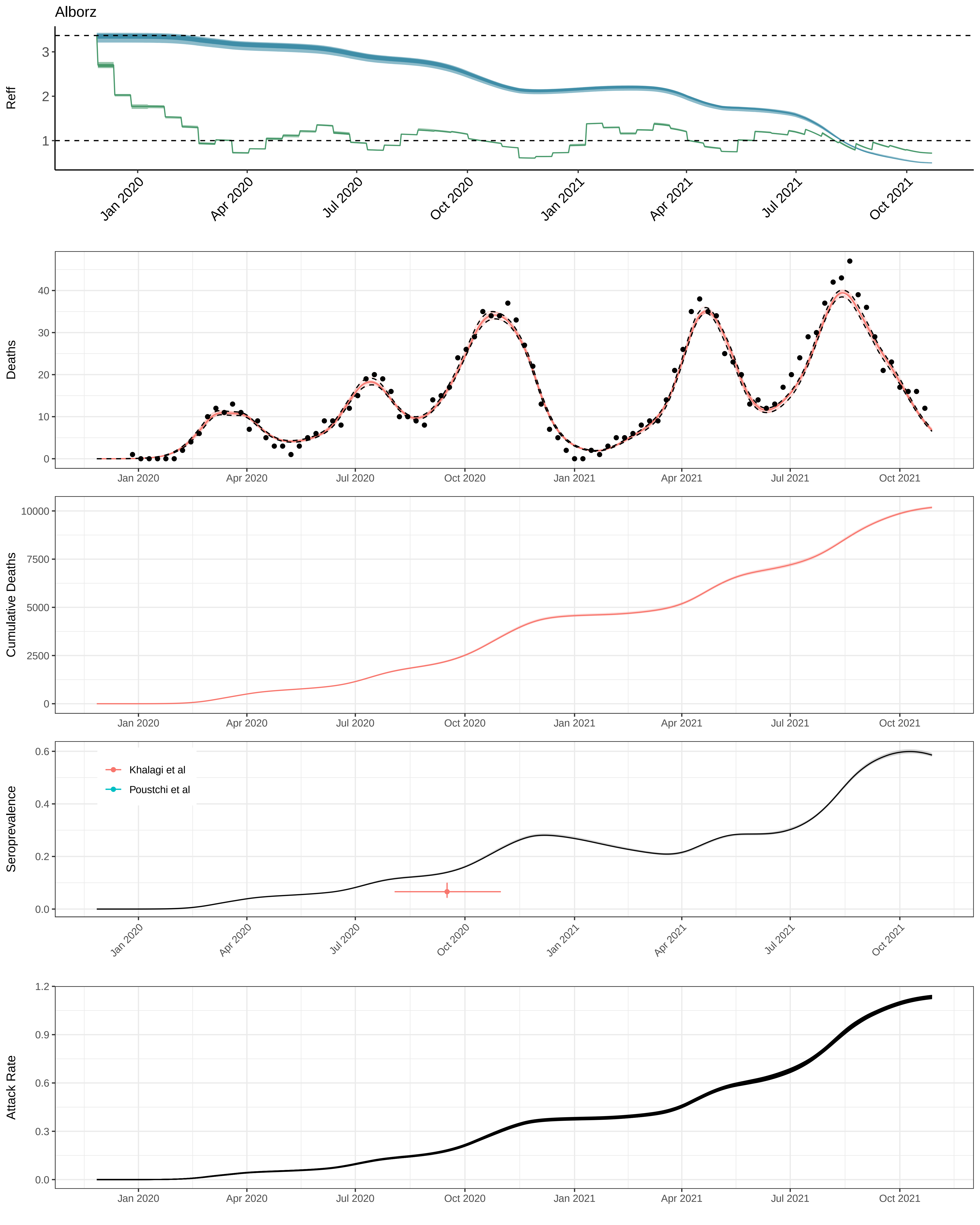

Alborz
Reff
Deaths
Cumulative Deaths
Seroprevalence
Khalagi et al
Poustchi et al
Attack Rate
Jan 2020
Apr 2020
Jul 2020
Oct 2020
Jan 2021
Apr 2021
Jul 2021
Oct 2021

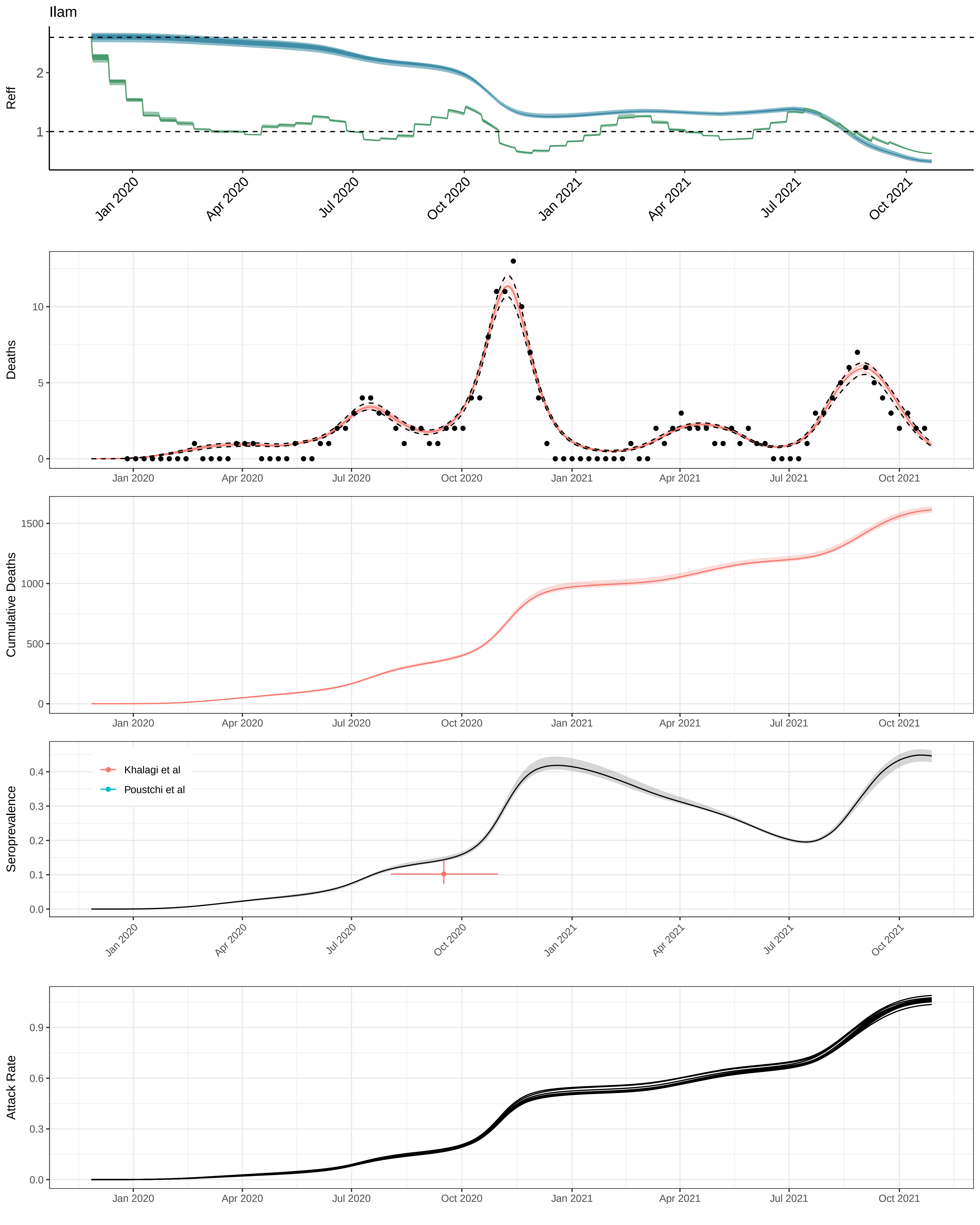
Ilam
Reff
Deaths
Cumulative Deaths
Seroprevalence
Khalagi et al
Poustchi et al
Attack Rate
Jan 2020
Apr 2020
Jul 2020
Oct 2020
Jan 2021
Apr 2021
Jul 2021
Oct 2021

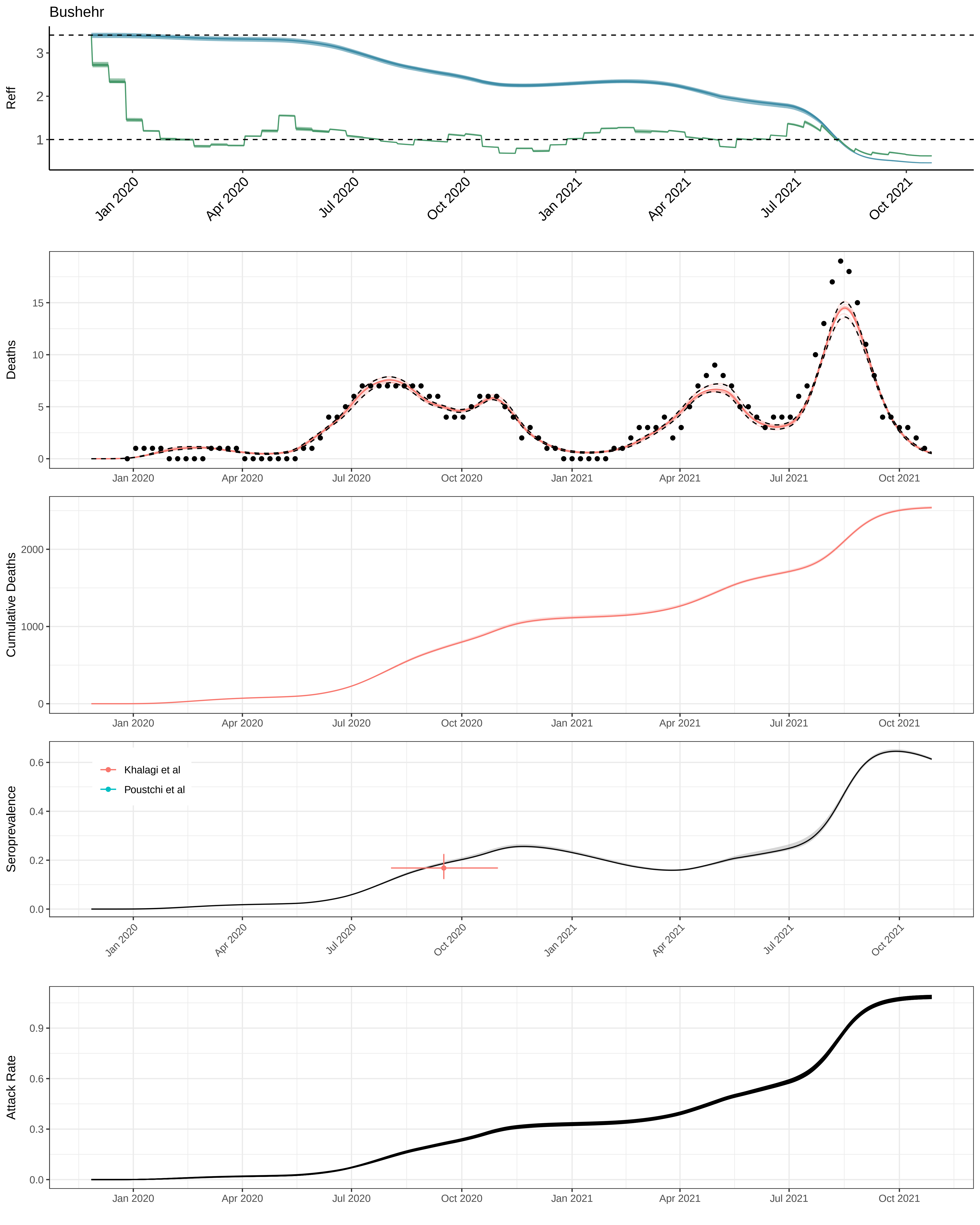

Bushehr
Reff
Deaths
Cumulative Deaths
Seroprevalence
Khalagi et al
Poustchi et al
Attack Rate
Jan 2020
Apr 2020
Jul 2020
Oct 2020
Jan 2021
Apr 2021
Jul 2021
Oct 2021

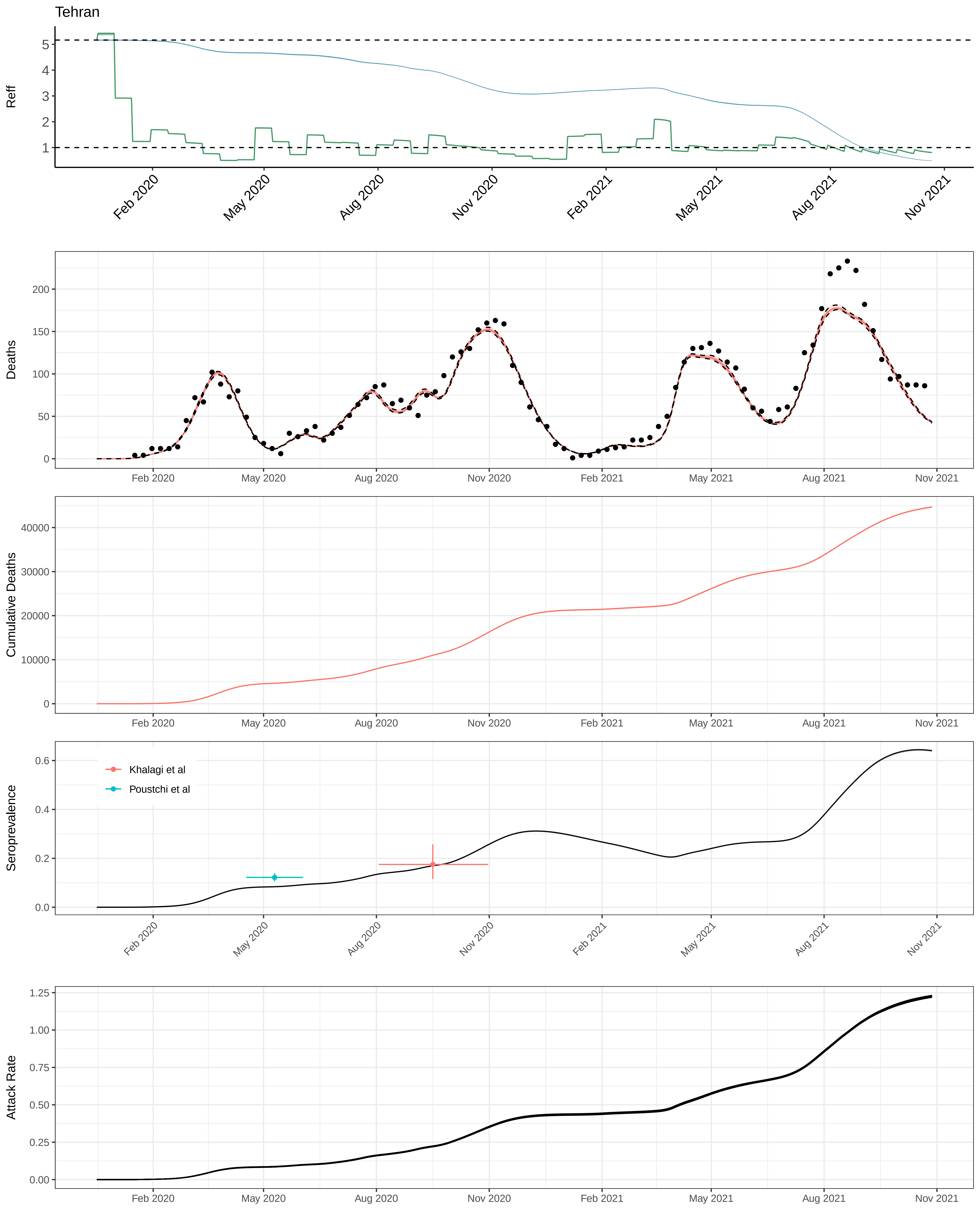

Tehran
Reff
Deaths
Cumulative Deaths
Seroprevalence
Khalagi et al
Poustchi et al
Attack Rate
Feb 2020
May 2020
Aug 2020
Nov 2020
Feb 2021
May 2021
Aug 2021
Nov 2021

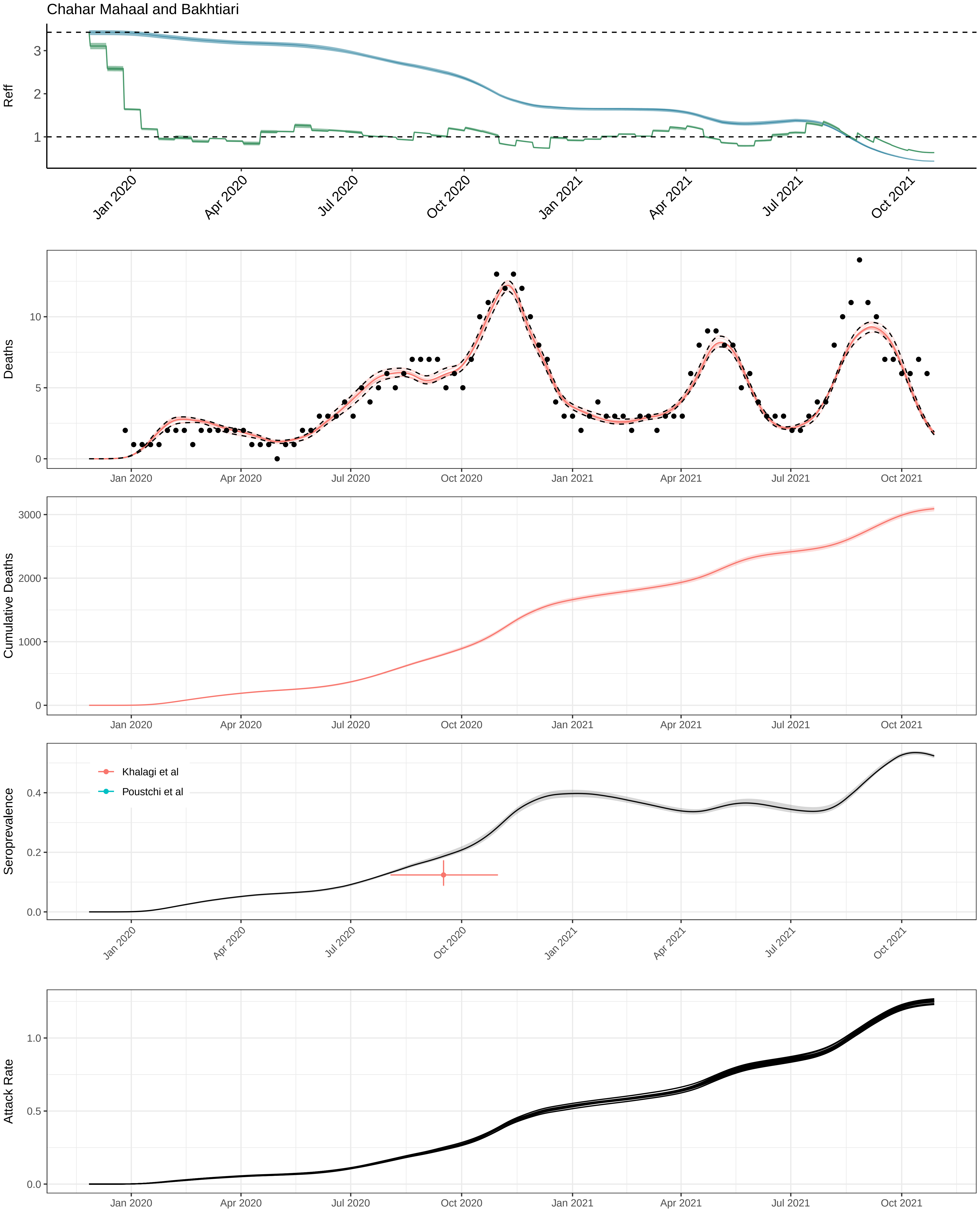
Chahar Mahaal and Bakhtiari
Reff
3
2
1
Jan 2020
Apr 2020
Jul 2020
Oct 2020
Jan 2021
Apr 2021
Jul 2021
Oct 2021
Deaths
10
5
0
Cumulative Deaths
3000
2000
1000
0
Seroprevalence
Khalagi et al
Poustchi et al
0.4
0.2
0.0
Attack Rate
1.0
0.5
0.0

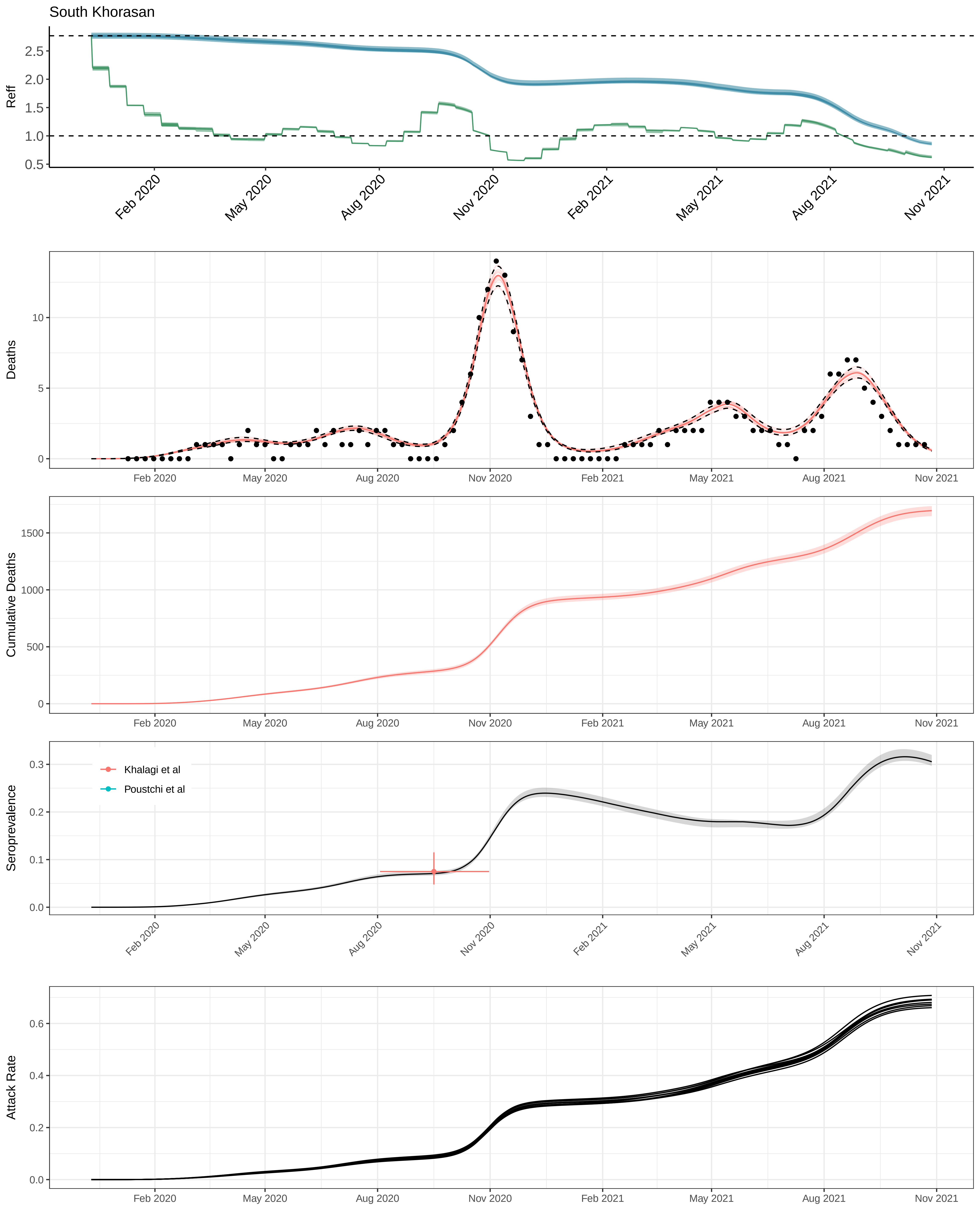

South Khorasan
Reff
2.5
2.0
1.5
1.0
0.5
Feb 2020
May 2020
Aug 2020
Nov 2020
Feb 2021
May 2021
Aug 2021
Nov 2021
Deaths
10
5
0
Cumulative Deaths
1500
1000
500
0
Seroprevalence
0.3
0.2
0.1
0.0
Khalagi et al
Poustchi et al
Attack Rate
0.6
0.4
0.2
0.0

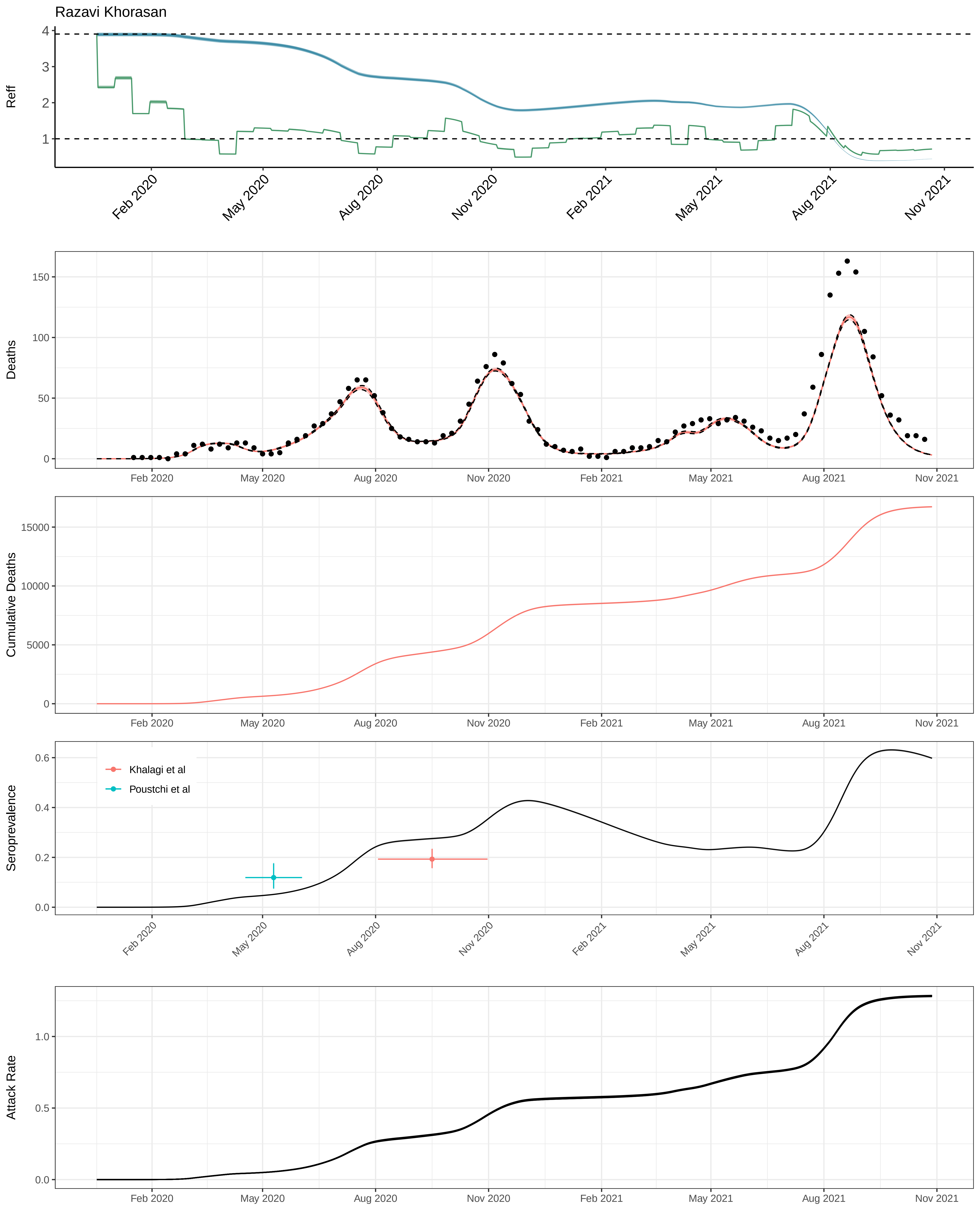

Razavi Khorasan
Reff
Deaths
Cumulative Deaths
Seroprevalence
Khalagi et al
Poustchi et al
Attack Rate
Feb 2020
May 2020
Aug 2020
Nov 2020
Feb 2021
May 2021
Aug 2021
Nov 2021

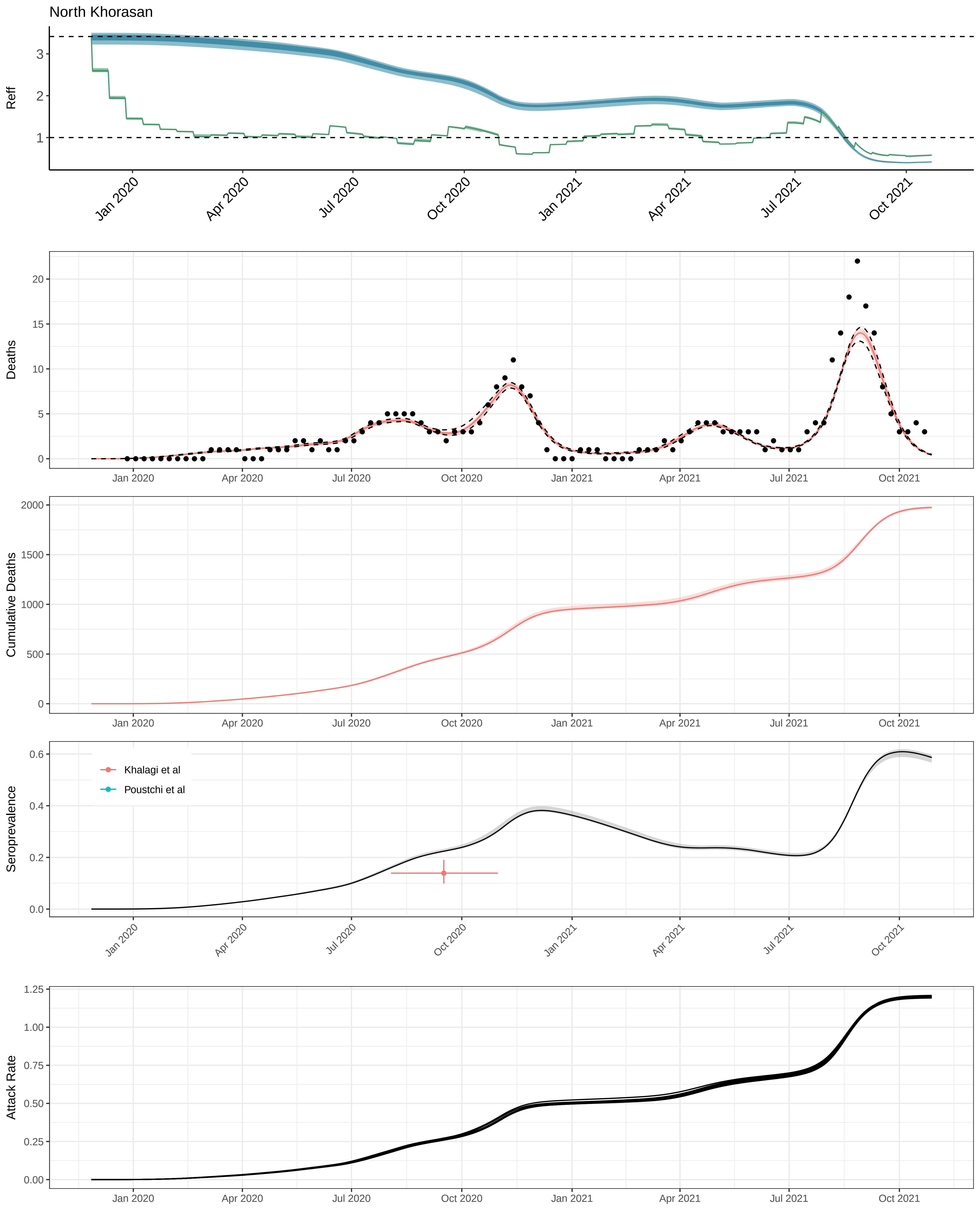
North Khorasan
Reff
Jan 2020
Apr 2020
Jul 2020
Oct 2020
Jan 2021
Apr 2021
Jul 2021
Oct 2021
Deaths
Cumulative Deaths
Seroprevalence
Khalagi et al
Poustchi et al
Attack Rate

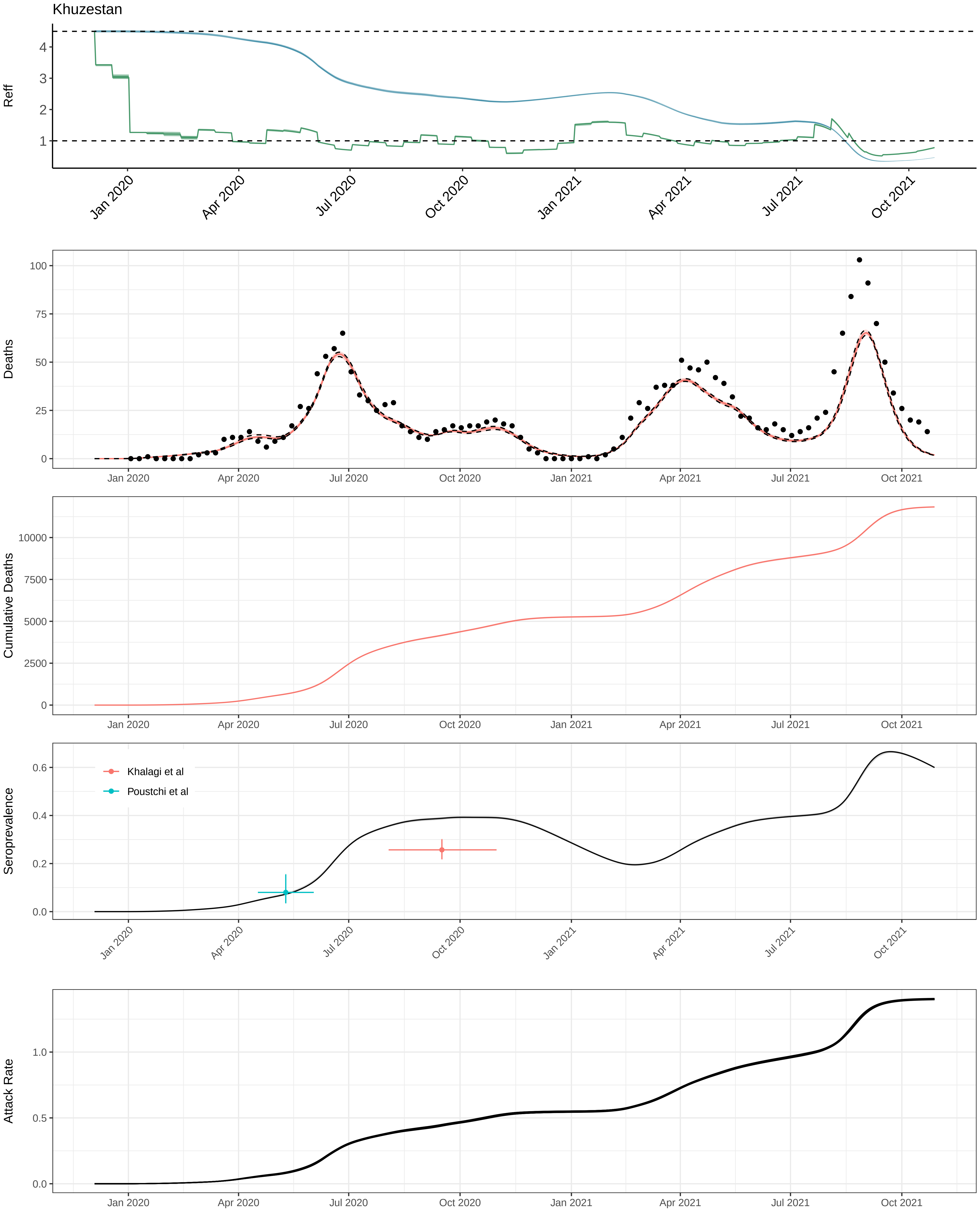
Khuzestan
Reff
Deaths
Cumulative Deaths
Seroprevalence
Khalagi et al
Poustchi et al
Attack Rate
Jan 2020
Apr 2020
Jul 2020
Oct 2020
Jan 2021
Apr 2021
Jul 2021
Oct 2021

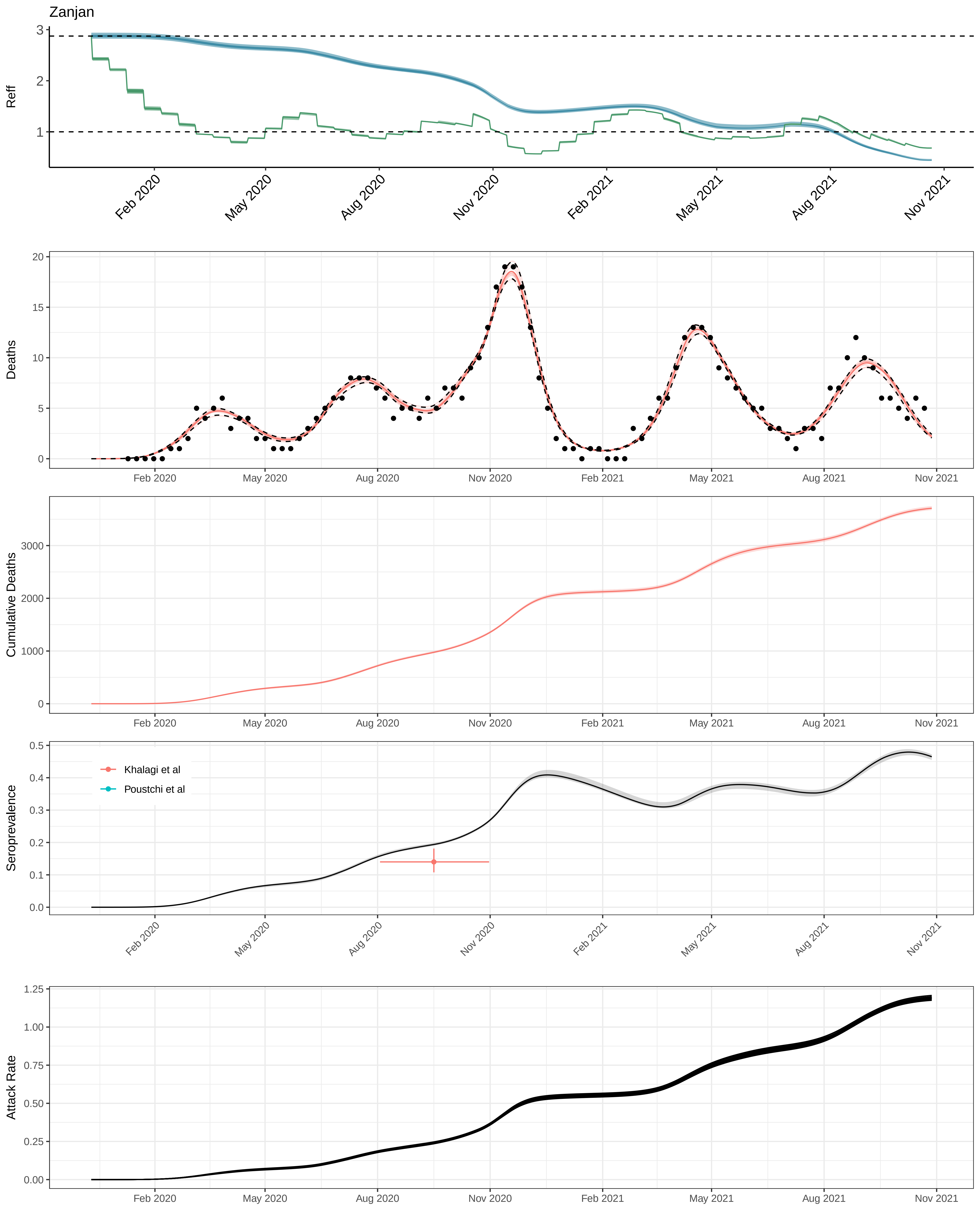
Zanjan
Reff
Deaths
Cumulative Deaths
Seroprevalence
Khalagi et al
Poustchi et al
Attack Rate
Feb 2020
May 2020
Aug 2020
Nov 2020
Feb 2021
May 2021
Aug 2021
Nov 2021

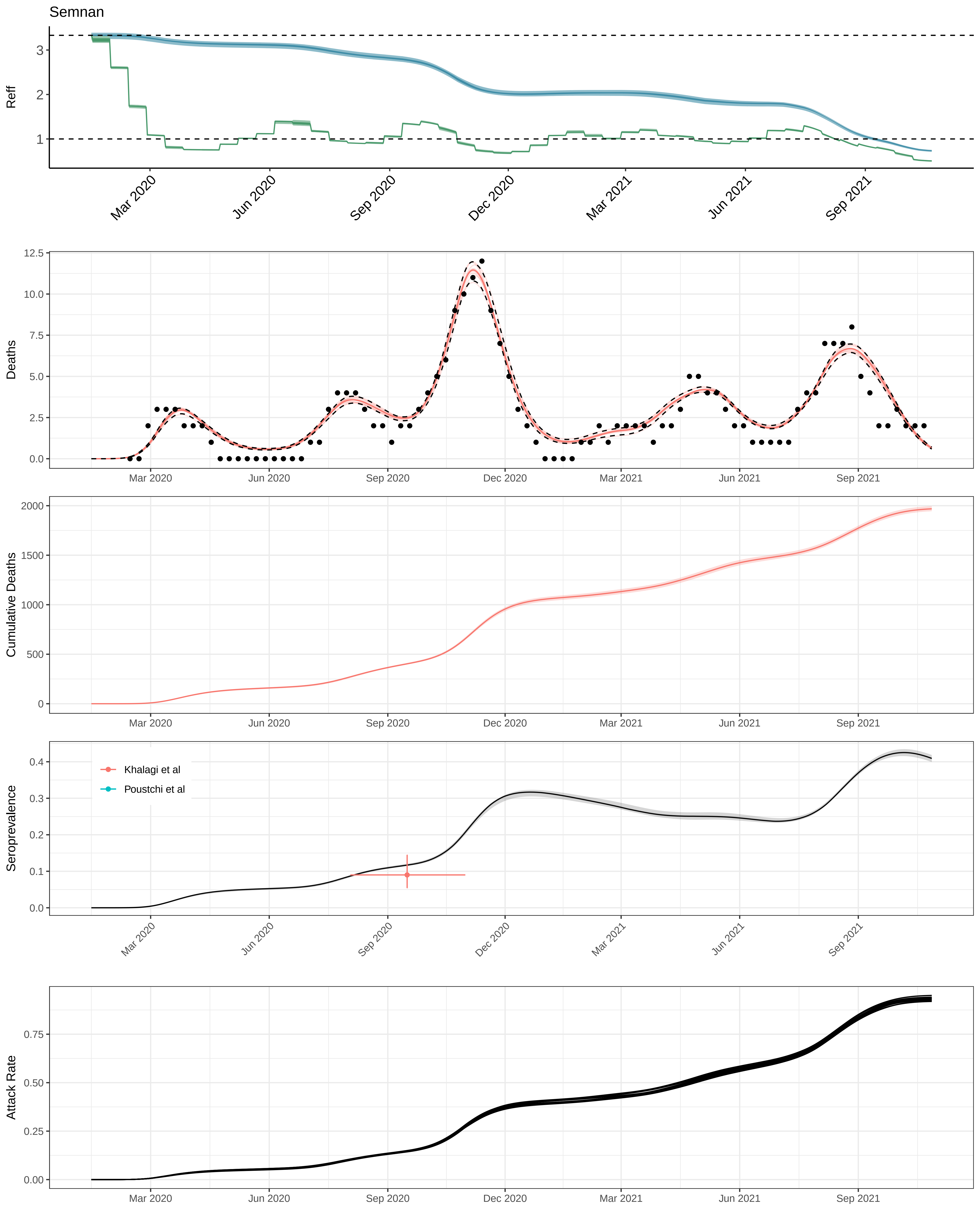
Semnan
Reff
3
2
1
Mar 2020
Jun 2020
Sep 2020
Dec 2020
Mar 2021
Jun 2021
Sep 2021
Deaths
12.5
10.0
7.5
5.0
2.5
0.0
Cumulative Deaths
2000
1500
1000
500
0
Seroprevalence
0.4
0.3
0.2
0.1
0.0
Khalagi et al
Poustchi et al
Attack Rate
0.75
0.50
0.25
0.00

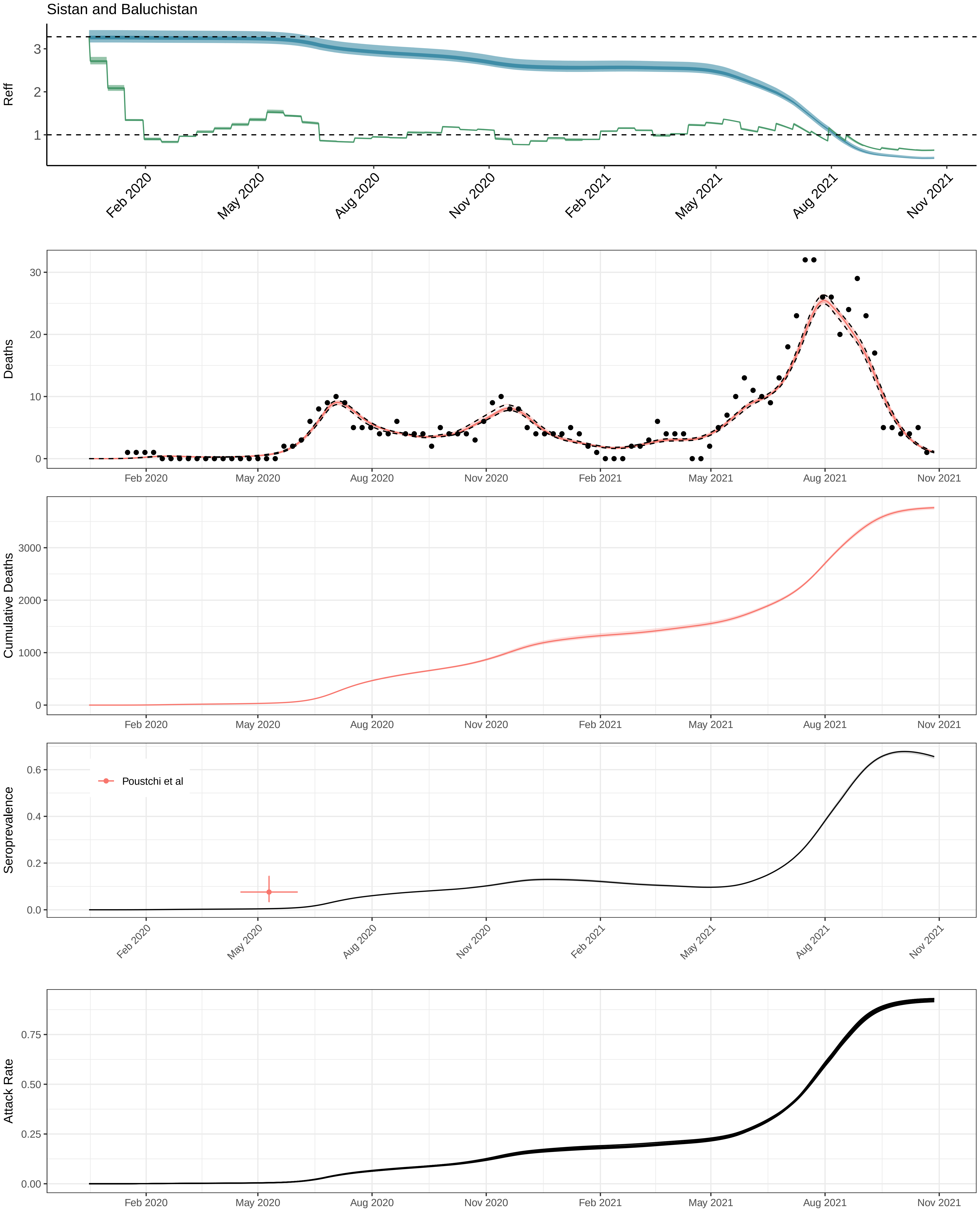
Sistan and Baluchistan
Reff
Deaths
Cumulative Deaths
Seroprevalence
Poustchi et al
Attack Rate
Feb 2020
May 2020
Aug 2020
Nov 2020
Feb 2021
May 2021
Aug 2021
Nov 2021

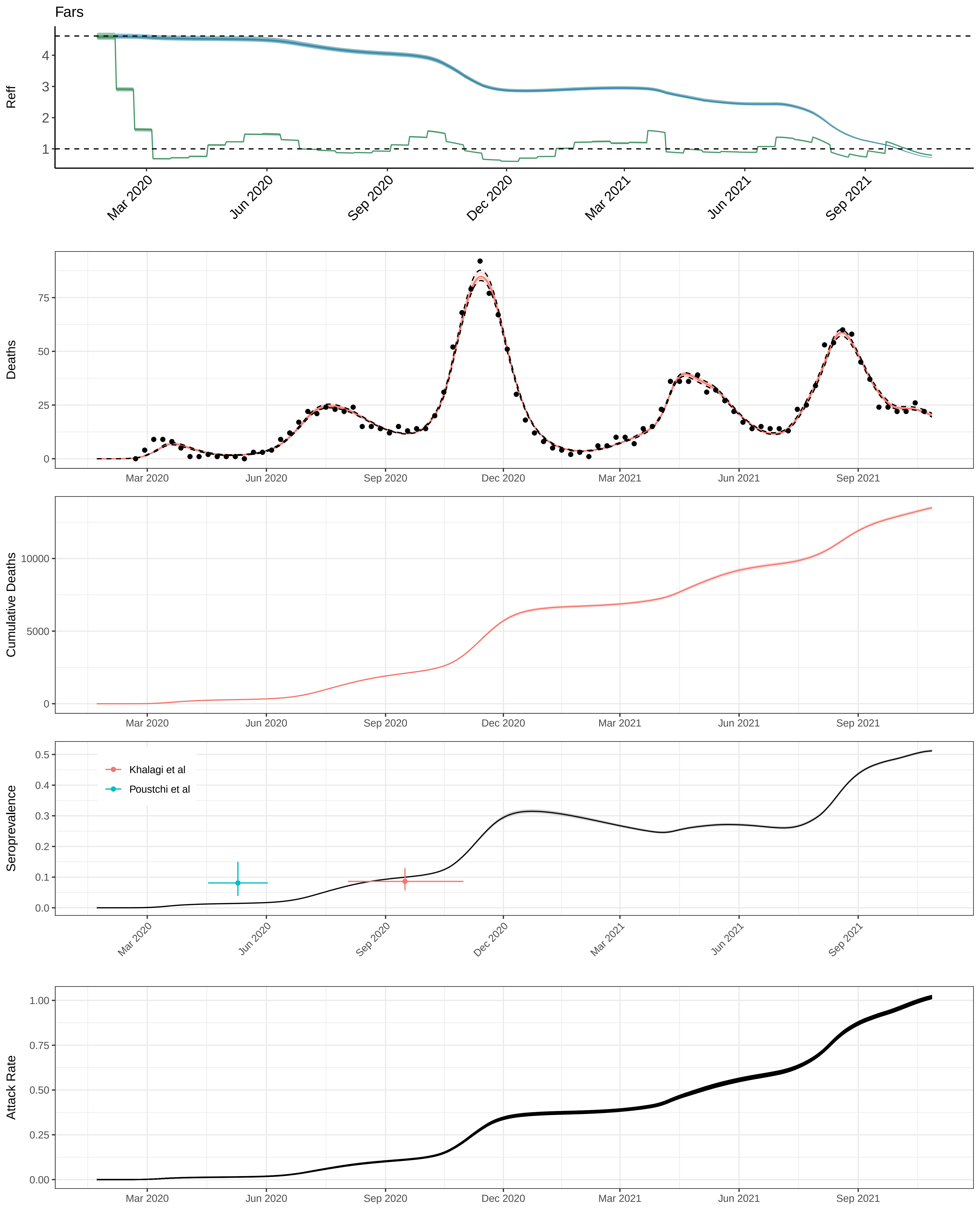

Fars
Reff
Deaths
Cumulative Deaths
Seroprevalence
Attack Rate
Khalagi et al
Poustchi et al
Mar 2020
Jun 2020
Sep 2020
Dec 2020
Mar 2021
Jun 2021
Sep 2021

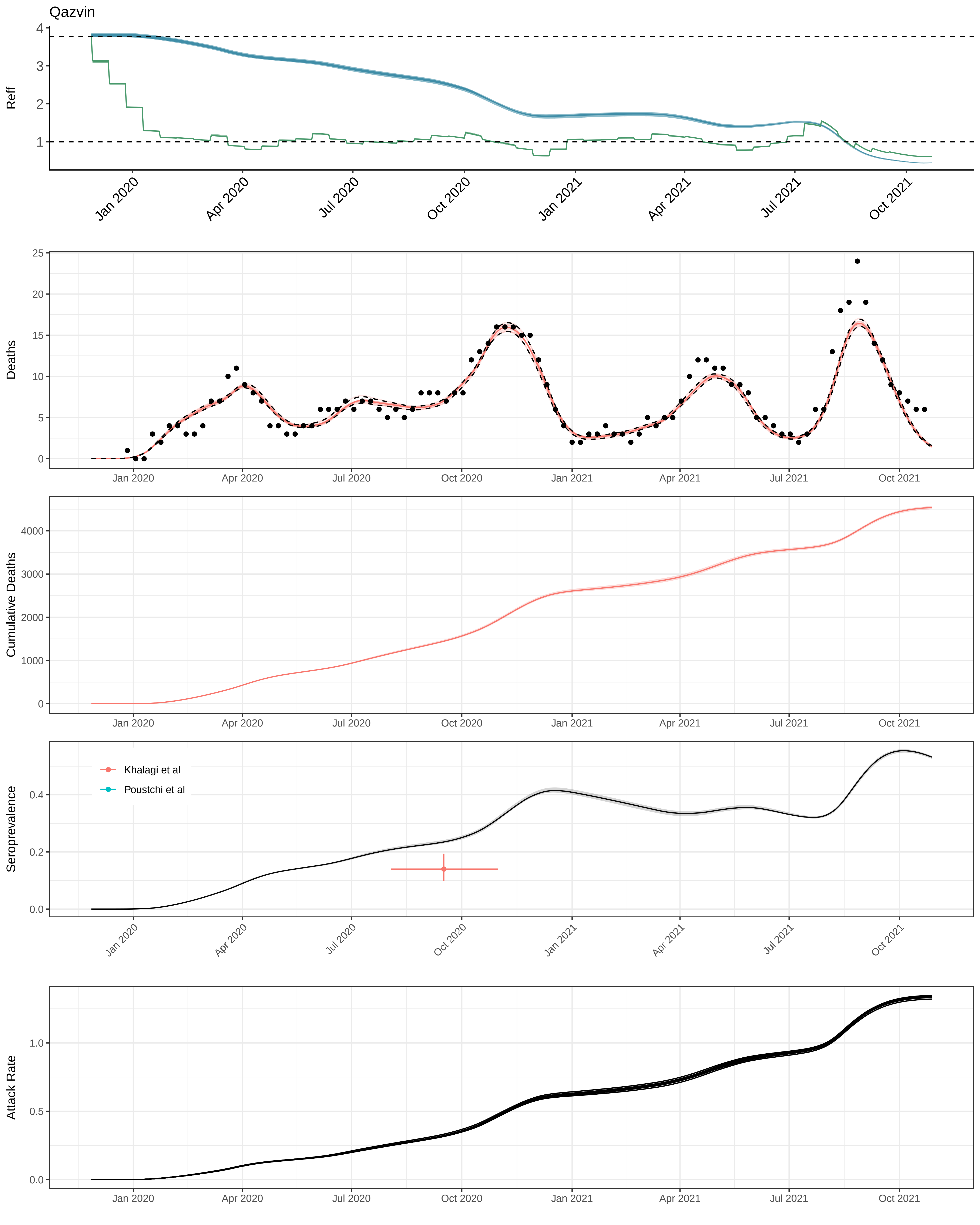

Qazvin
Reff
Deaths
Cumulative Deaths
Seroprevalence
Attack Rate
Khalagi et al
Poustchi et al
Jan 2020
Apr 2020
Jul 2020
Oct 2020
Jan 2021
Apr 2021
Jul 2021
Oct 2021

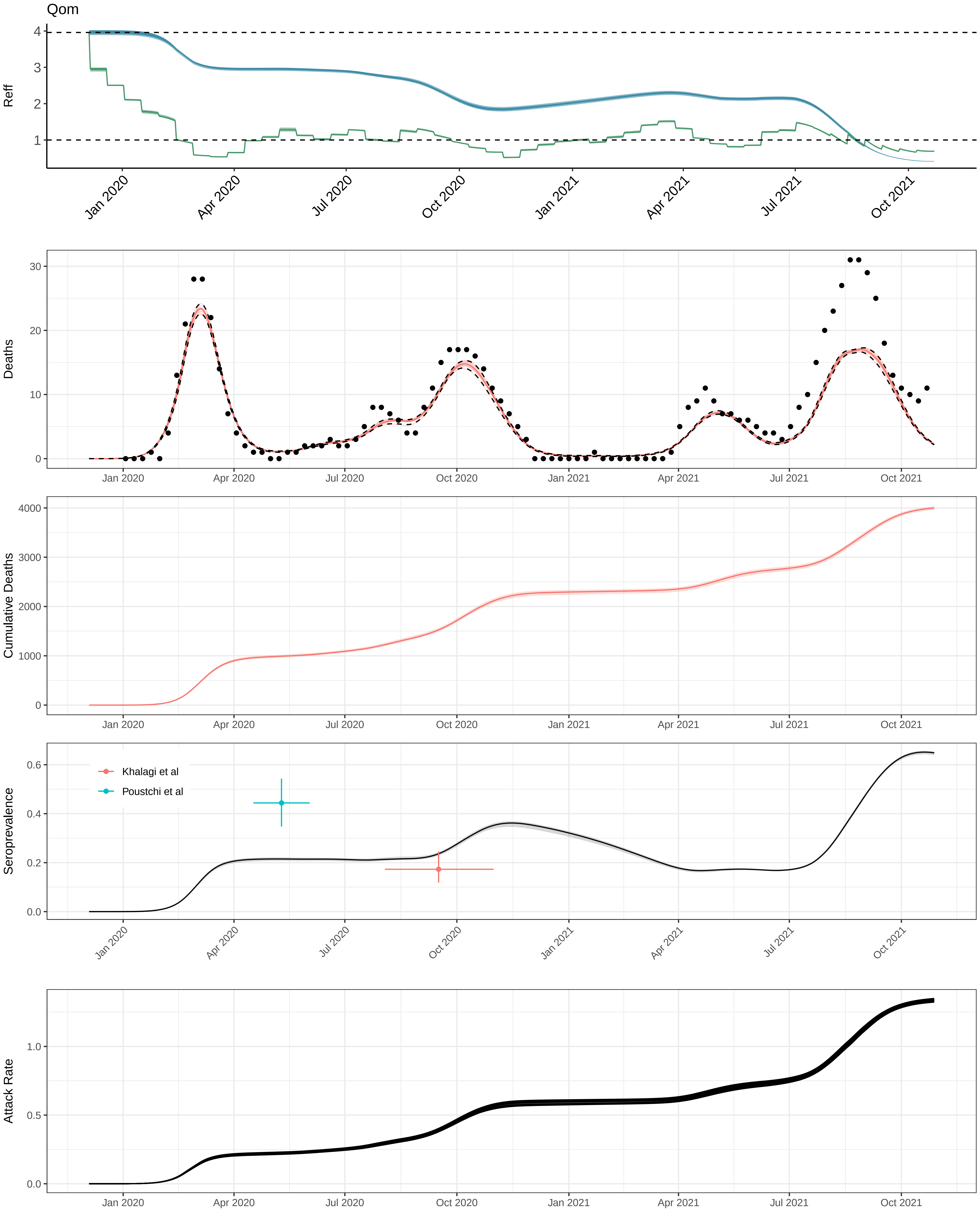

Qom
Reff
4
3
2
1
Jan 2020
Apr 2020
Jul 2020
Oct 2020
Jan 2021
Apr 2021
Jul 2021
Oct 2021
Deaths
30
20
10
0
Cumulative Deaths
4000
3000
2000
1000
0
Seroprevalence
0.6
0.4
0.2
0.0
Khalagi et al
Poustchi et al
Attack Rate
1.0
0.5
0.0

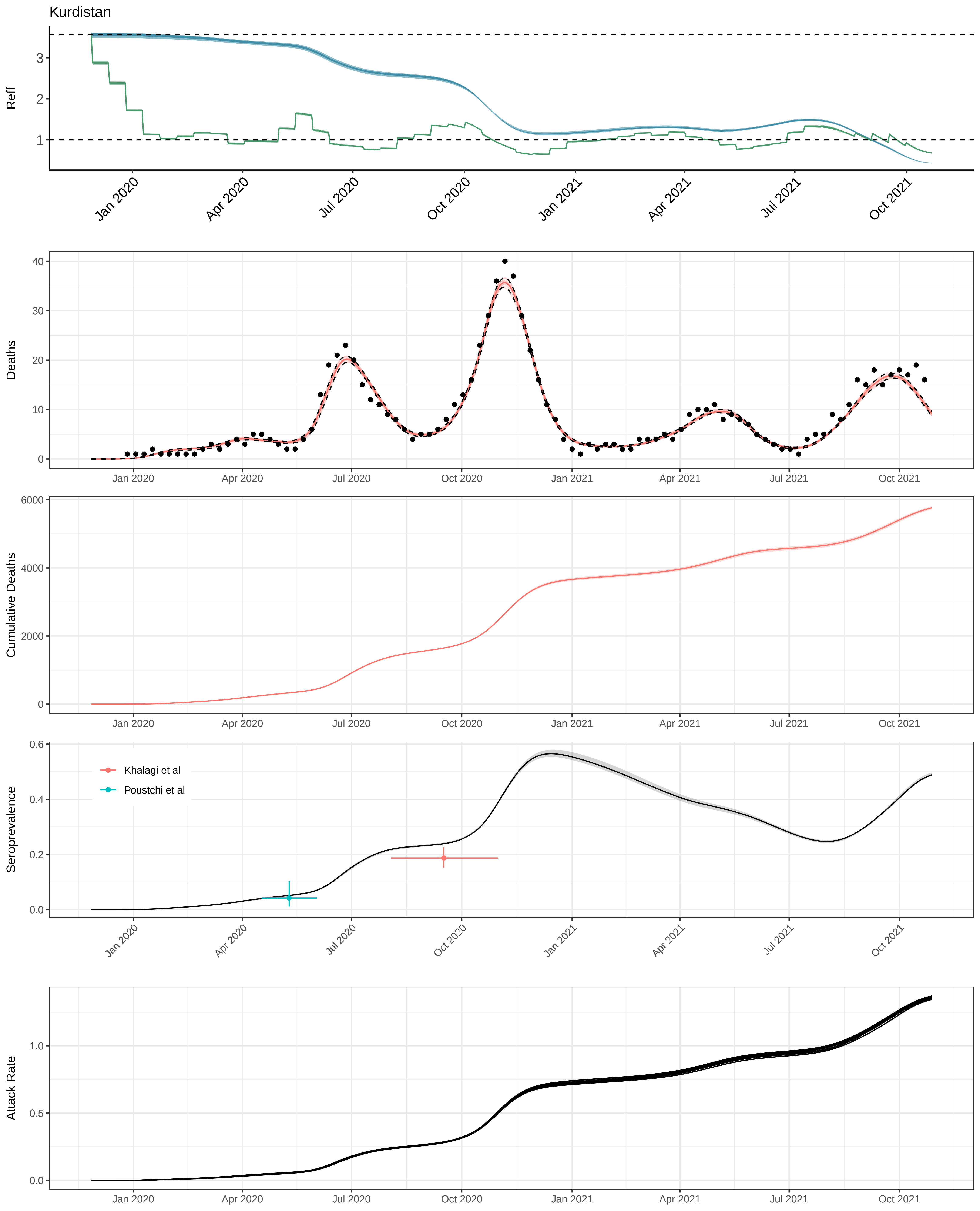

Kurdistan
Reff
Jan 2020
Apr 2020
Jul 2020
Oct 2020
Jan 2021
Apr 2021
Jul 2021
Oct 2021
Deaths
Cumulative Deaths
Seroprevalence
Khalagi et al
Poustchi et al
Attack Rate

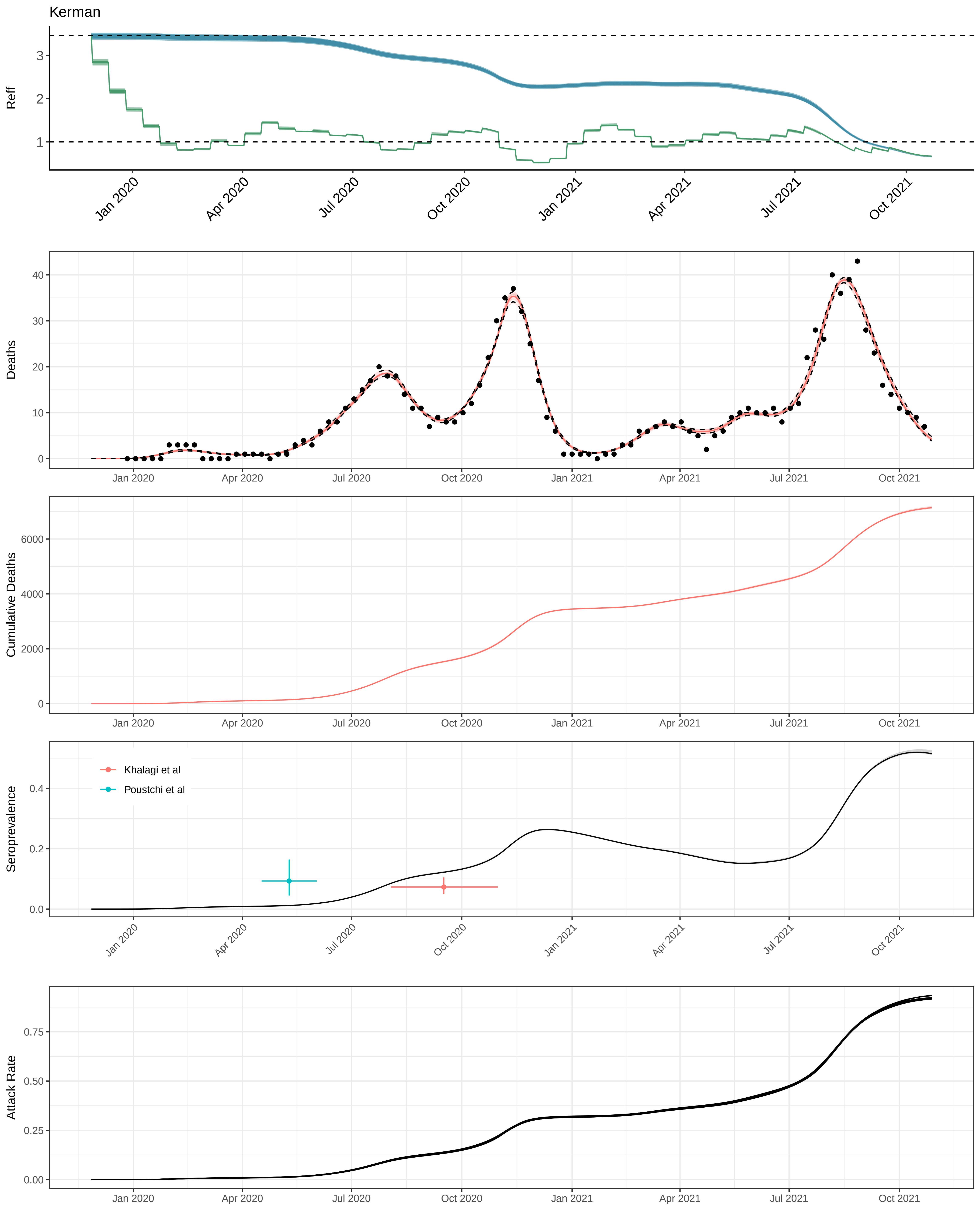

Kerman
Reff
Deaths
Cumulative Deaths
Seroprevalence
Khalagi et al
Poustchi et al
Attack Rate
Jan 2020
Apr 2020
Jul 2020
Oct 2020
Jan 2021
Apr 2021
Jul 2021
Oct 2021

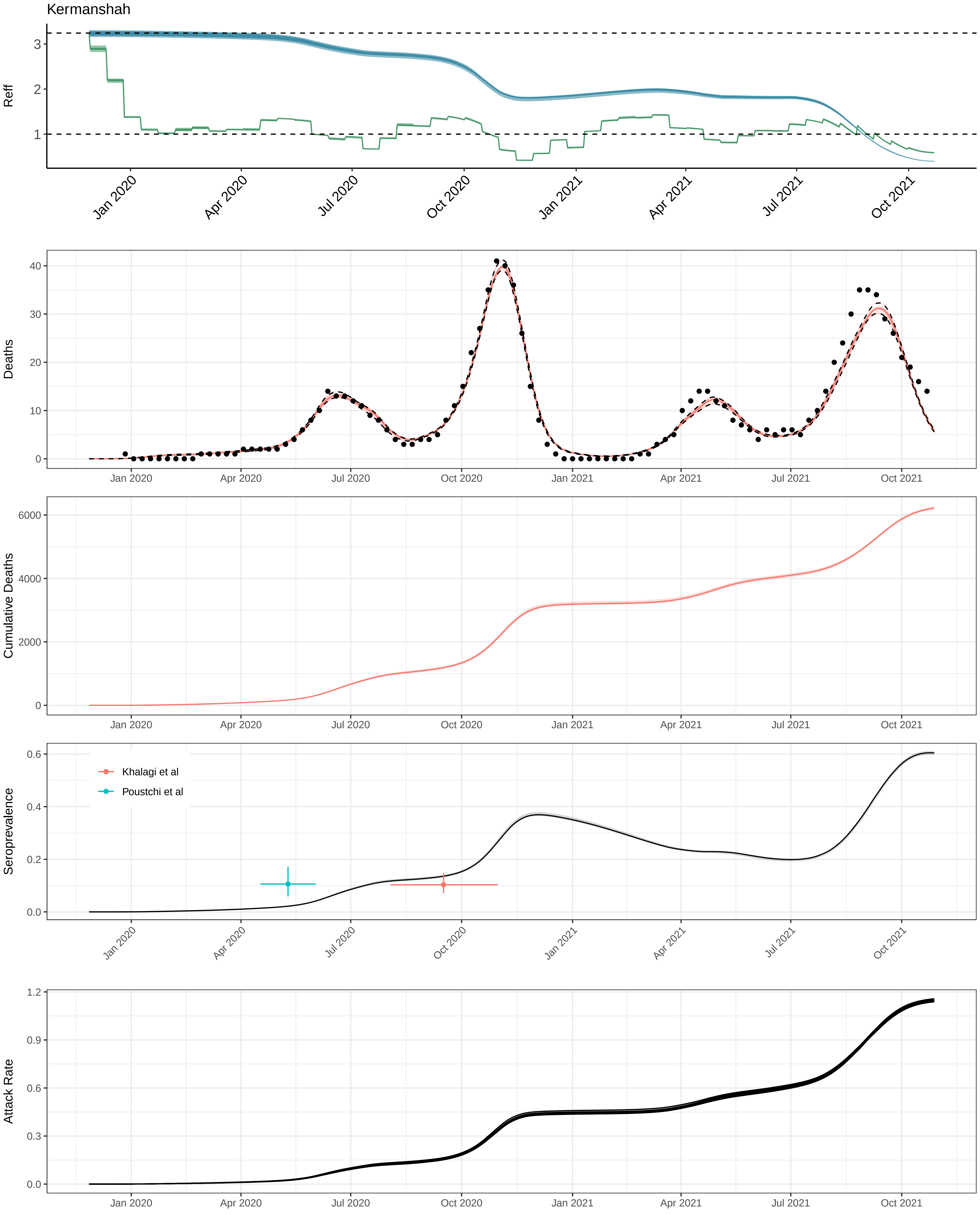
Kermanshah
Reff
3
2
1
Jan 2020
Apr 2020
Jul 2020
Oct 2020
Jan 2021
Apr 2021
Jul 2021
Oct 2021
Deaths
40
30
20
10
0
Cumulative Deaths
6000
4000
2000
0
Seroprevalence
0.6
0.4
0.2
0.0
Khalagi et al
Poustchi et al
Attack Rate
1.2
0.9
0.6
0.3
0.0

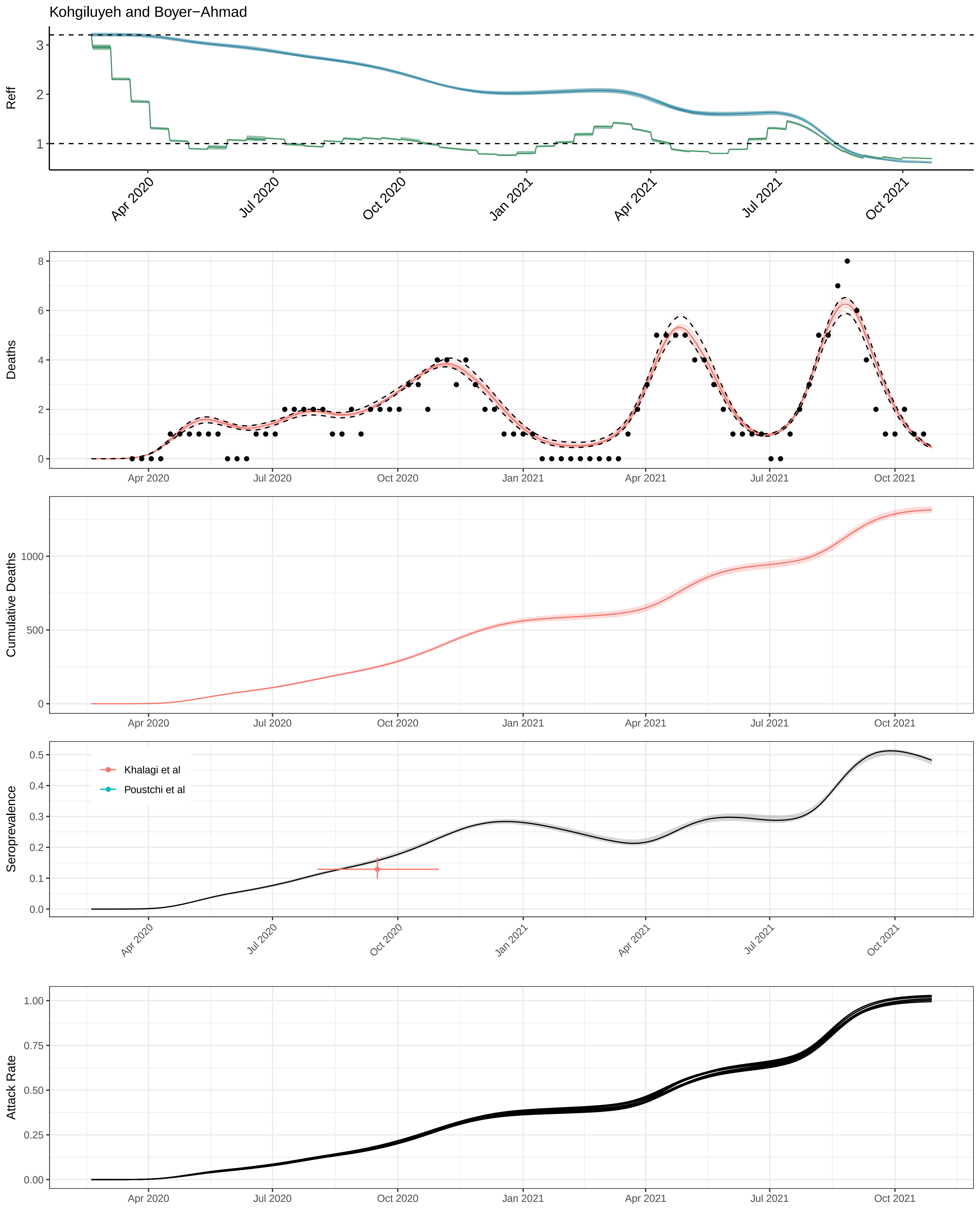
Kohgiluyeh and Boyer−Ahmad
Reff
Apr 2020
Jul 2020
Oct 2020
Jan 2021
Apr 2021
Jul 2021
Oct 2021
Deaths
Cumulative Deaths
Seroprevalence
Khalagi et al
Poustchi et al
Attack Rate

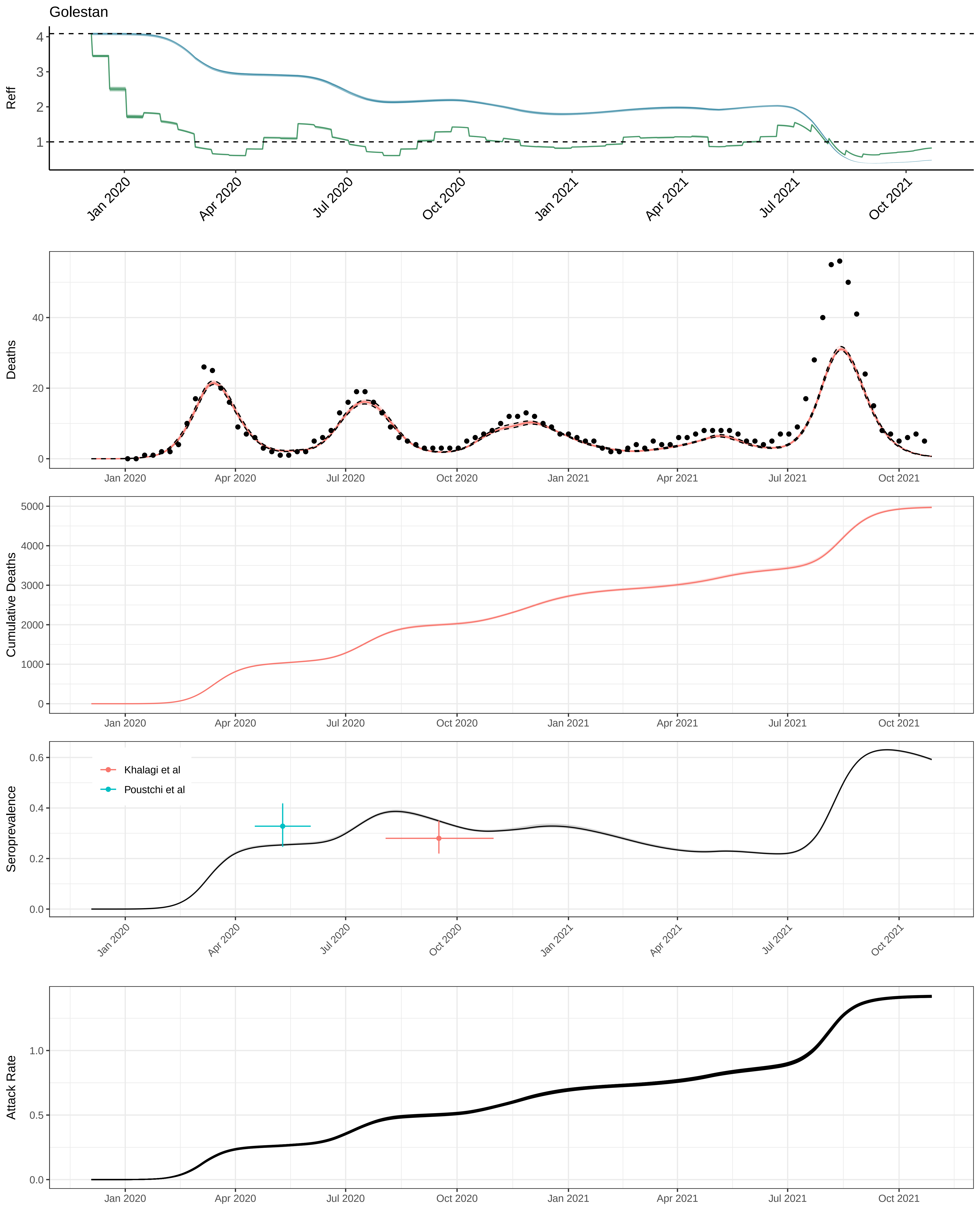

Golestan
Reff
4
3
2
1
Jan 2020
Apr 2020
Jul 2020
Oct 2020
Jan 2021
Apr 2021
Jul 2021
Oct 2021
Deaths
40
20
0
Cumulative Deaths
5000
4000
3000
2000
1000
0
Seroprevalence
0.6
0.4
0.2
0.0
Khalagi et al
Poustchi et al
Attack Rate
1.0
0.5
0.0

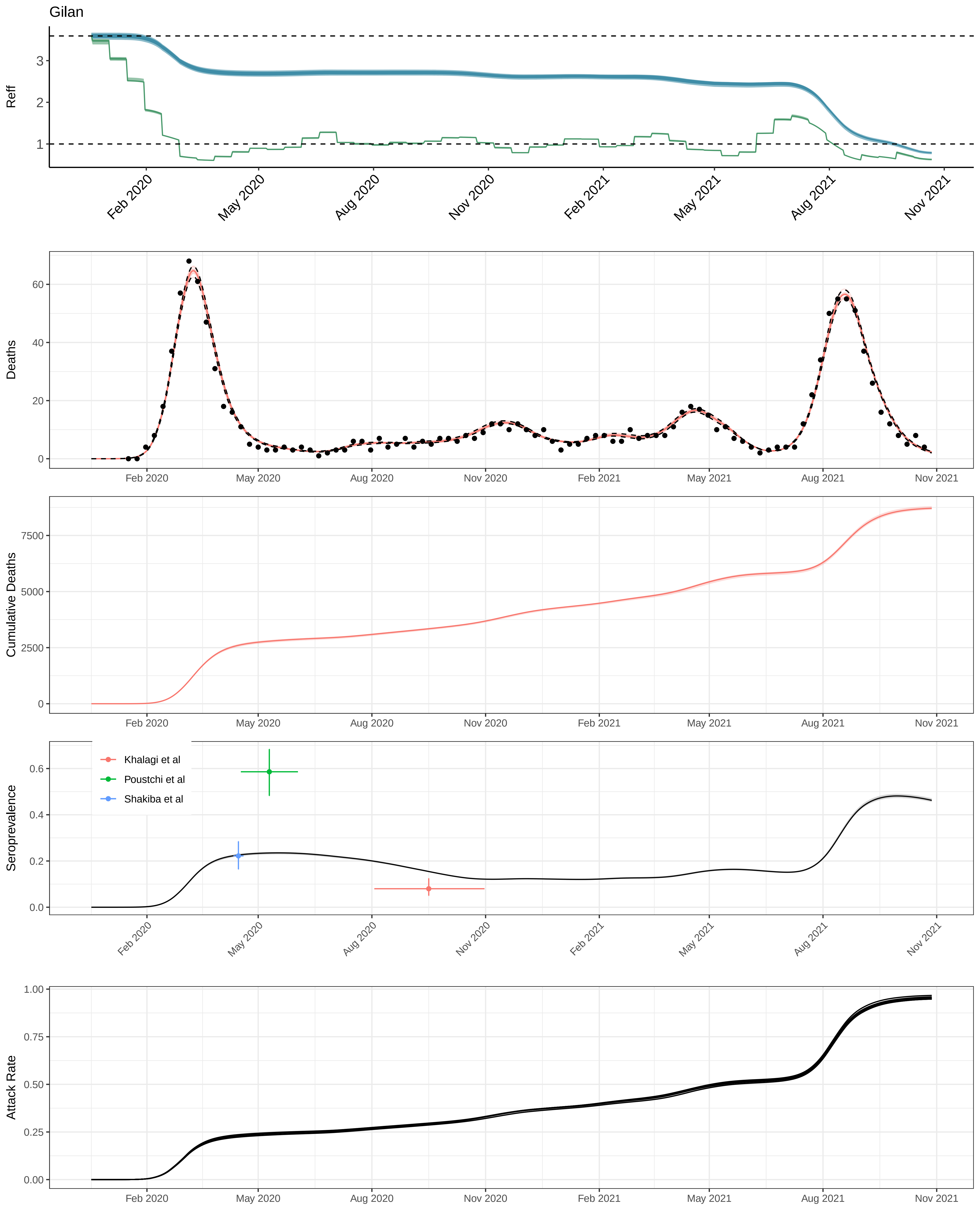

Gilan
Reff
Deaths
Cumulative Deaths
Seroprevalence
Khalagi et al
Poustchi et al
Shakiba et al
Attack Rate
Feb 2020
May 2020
Aug 2020
Nov 2020
Feb 2021
May 2021
Aug 2021
Nov 2021

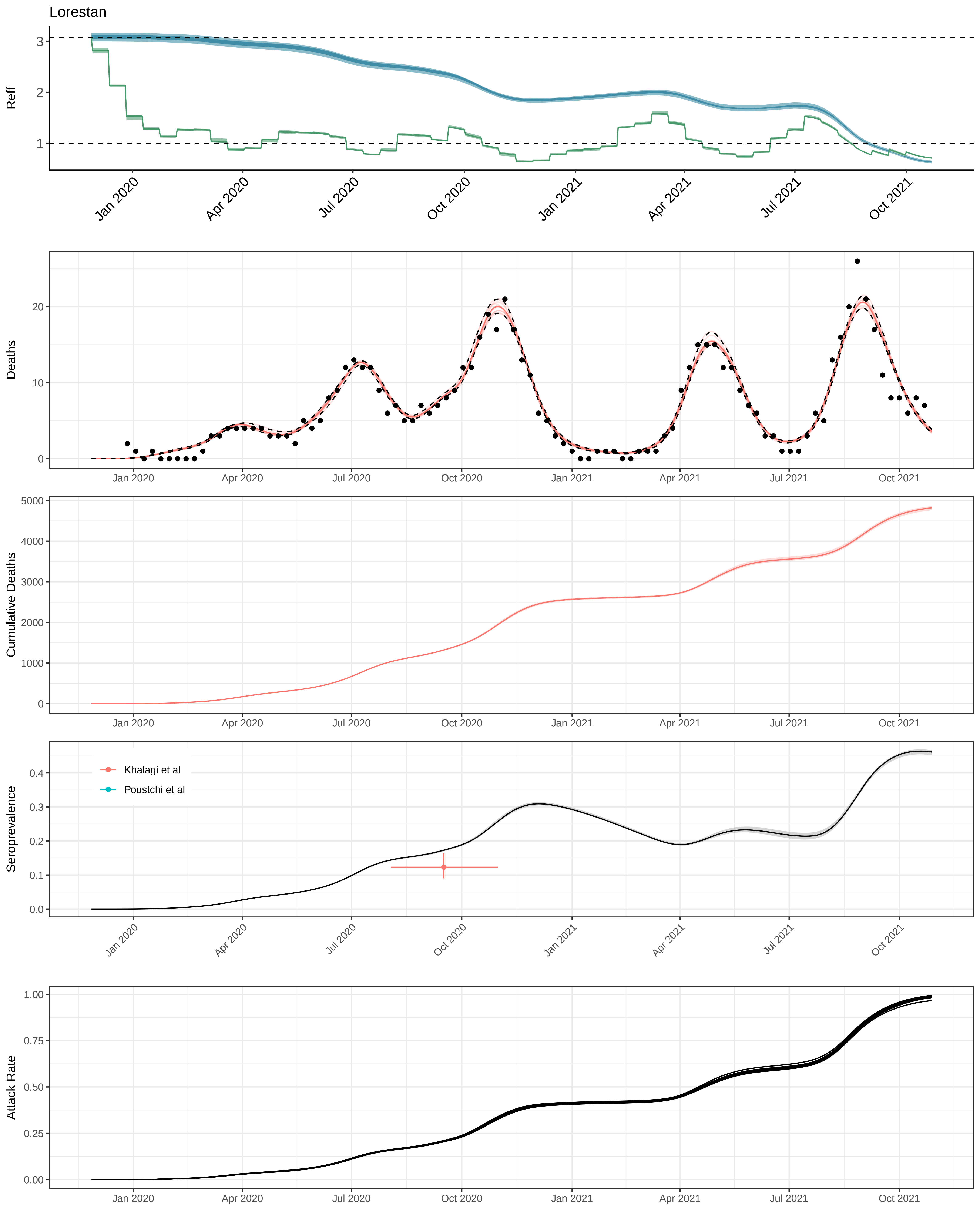

Lorestan
Reff
Deaths
Cumulative Deaths
Seroprevalence
Khalagi et al
Poustchi et al
Attack Rate
Jan 2020
Apr 2020
Jul 2020
Oct 2020
Jan 2021
Apr 2021
Jul 2021
Oct 2021

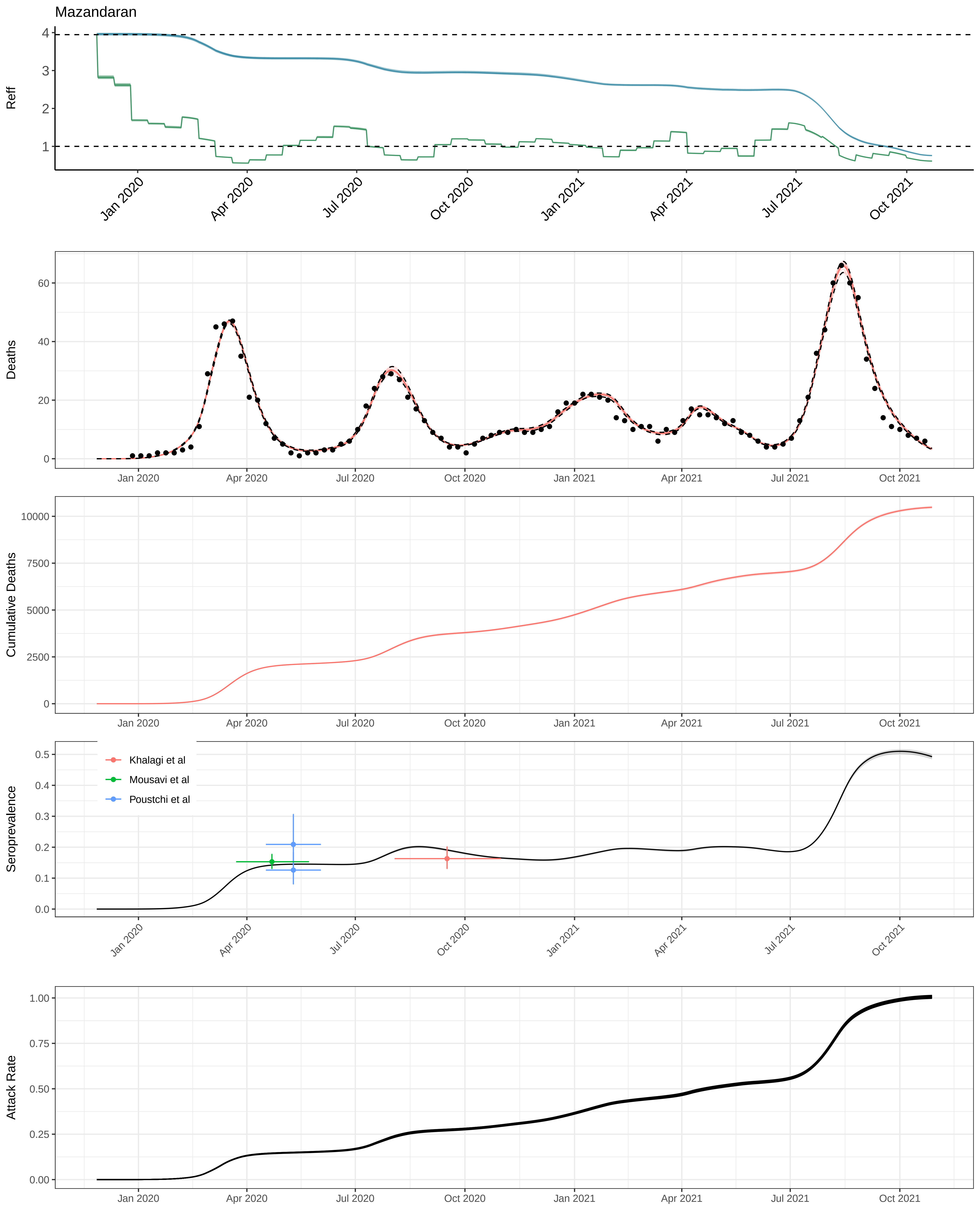
Mazandaran
Reff
4
3
2
1
Jan 2020
Apr 2020
Jul 2020
Oct 2020
Jan 2021
Apr 2021
Jul 2021
Oct 2021
Deaths
60
40
20
0
Cumulative Deaths
10000
7500
5000
2500
0
Seroprevalence
0.5
0.4
0.3
0.2
0.1
0.0
Khalagi et al
Mousavi et al
Poustchi et al
Attack Rate
1.00
0.75
0.50
0.25
0.00

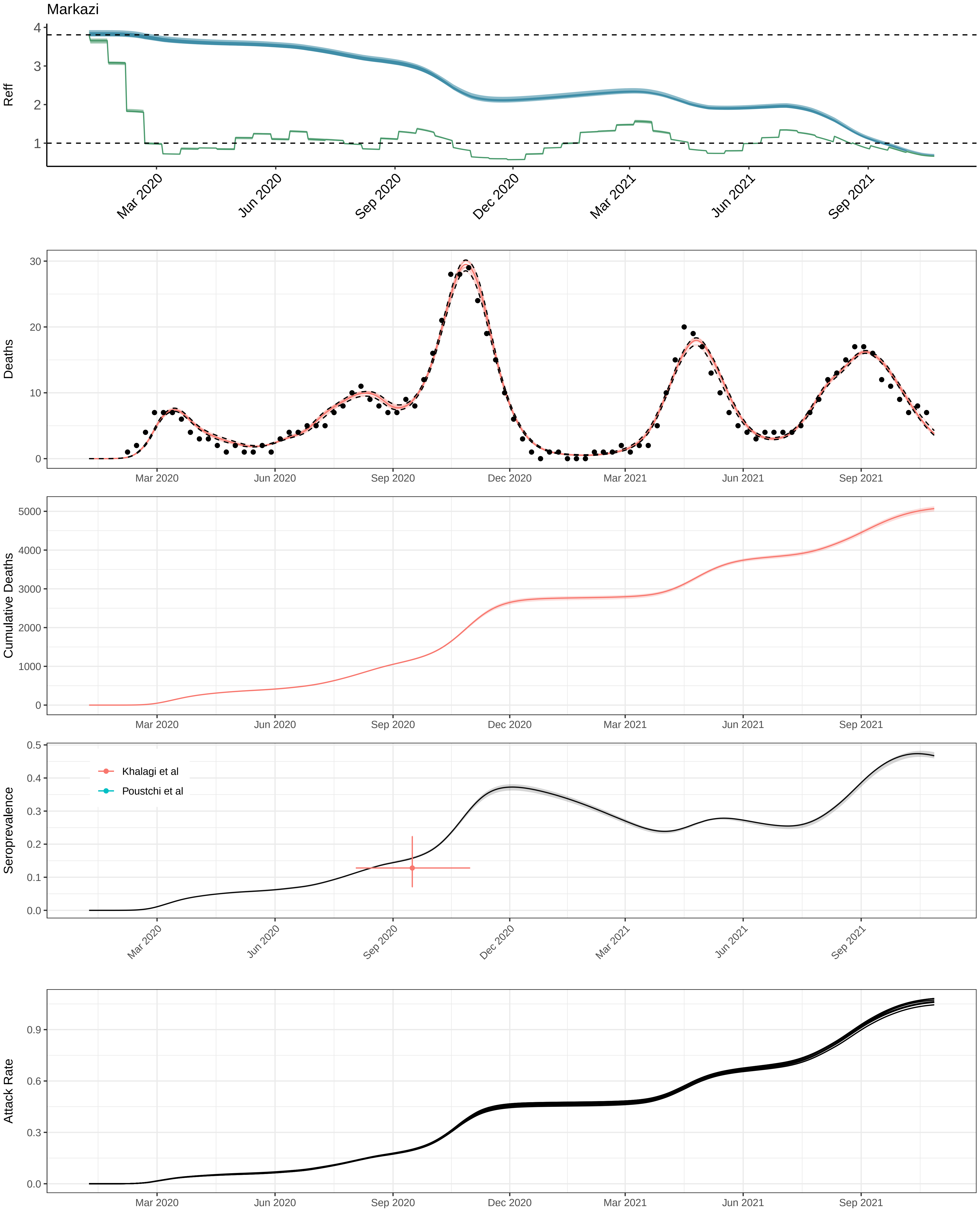
Markazi
Reff
Deaths
Cumulative Deaths
Seroprevalence
Khalagi et al
Poustchi et al
Attack Rate
Mar 2020
Jun 2020
Sep 2020
Dec 2020
Mar 2021
Jun 2021
Sep 2021

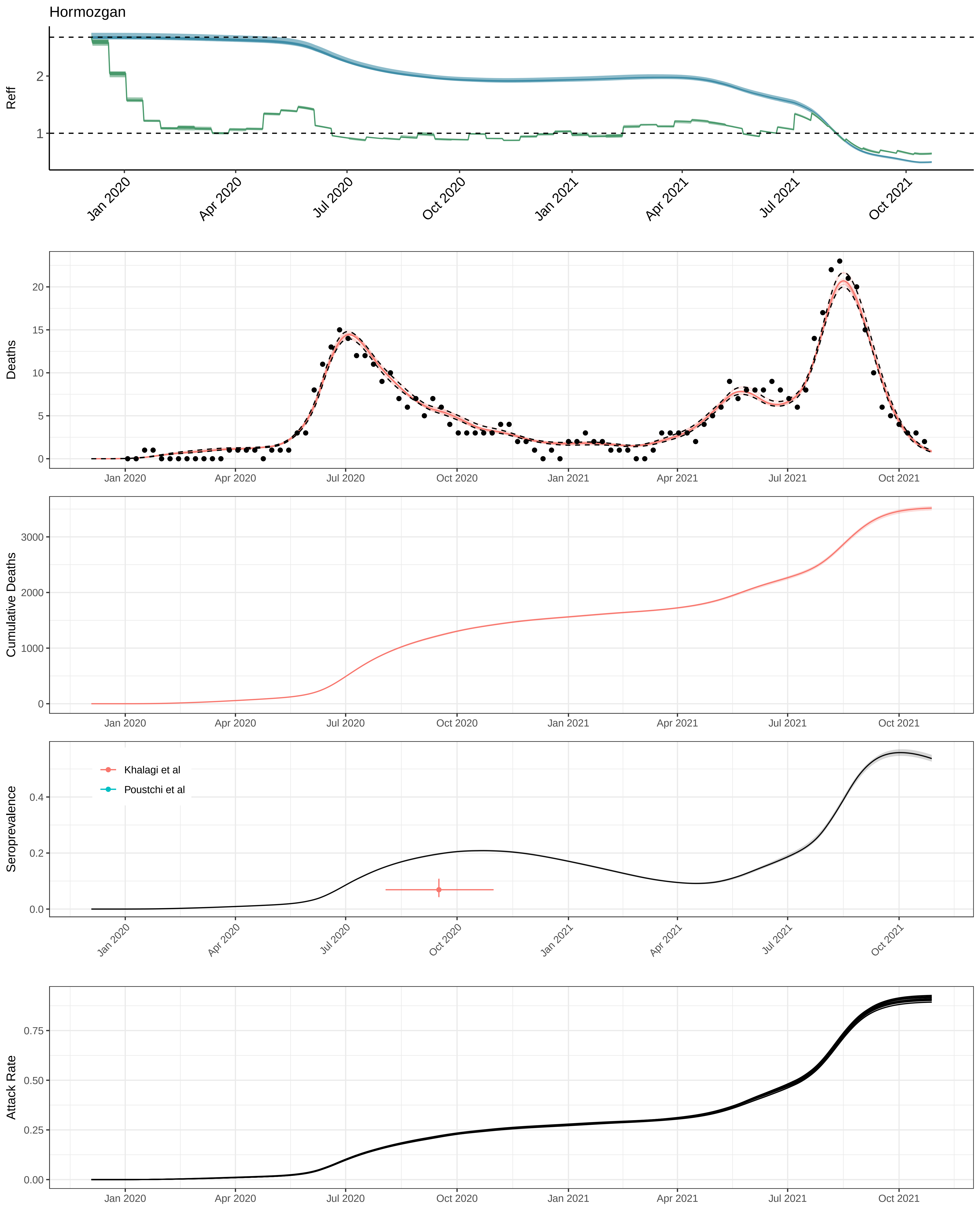

Hormozgan
Reff
2
1
Jan 2020
Apr 2020
Jul 2020
Oct 2020
Jan 2021
Apr 2021
Jul 2021
Oct 2021
Deaths
20
15
10
5
0
Cumulative Deaths
3000
2000
1000
0
Seroprevalence
Khalagi et al
Poustchi et al
0.4
0.2
0.0
Attack Rate
0.75
0.50
0.25
0.00

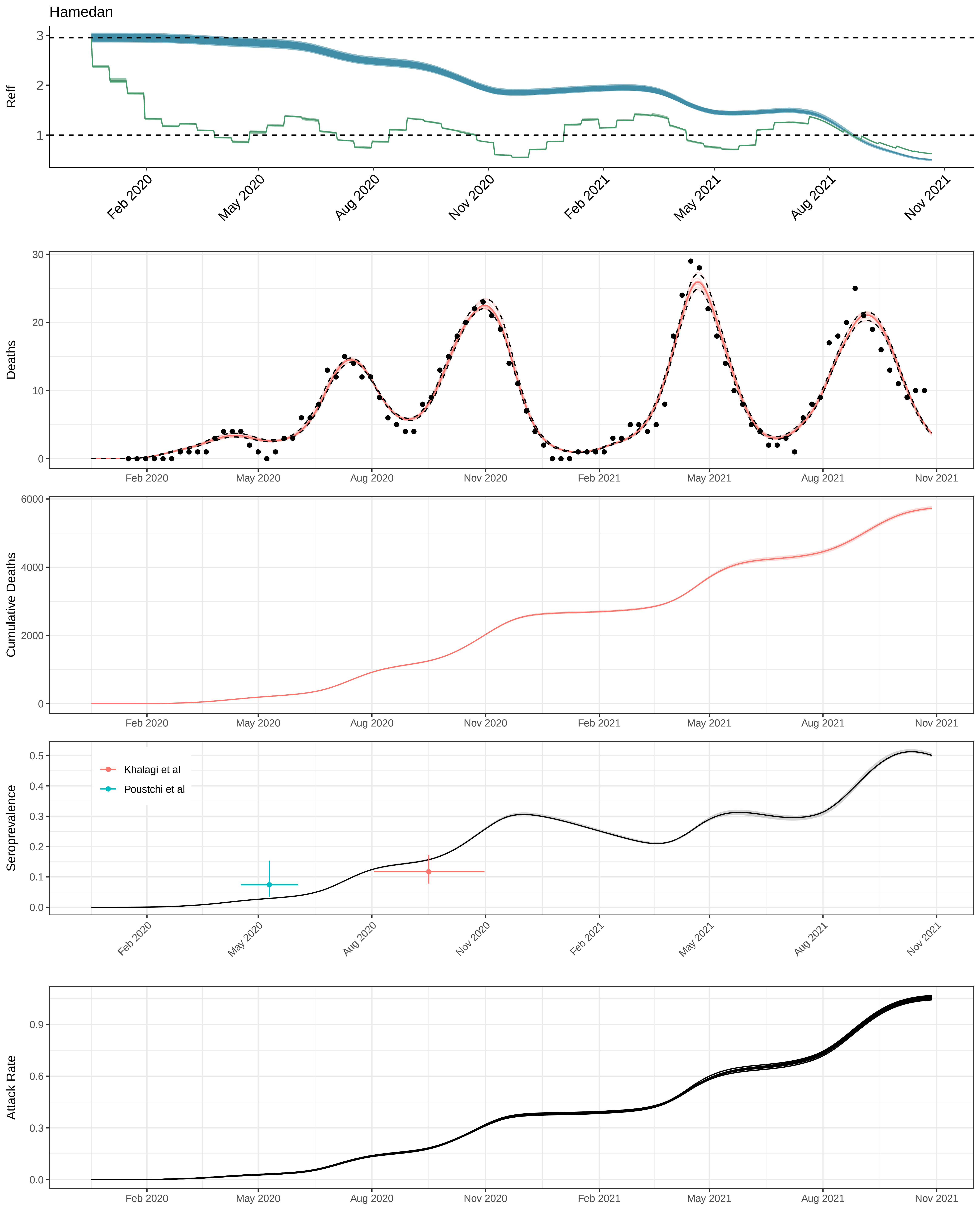

Hamedan
Reff
Deaths
Cumulative Deaths
Seroprevalence
Khalagi et al
Poustchi et al
Attack Rate
Feb 2020
May 2020
Aug 2020
Nov 2020
Feb 2021
May 2021
Aug 2021
Nov 2021

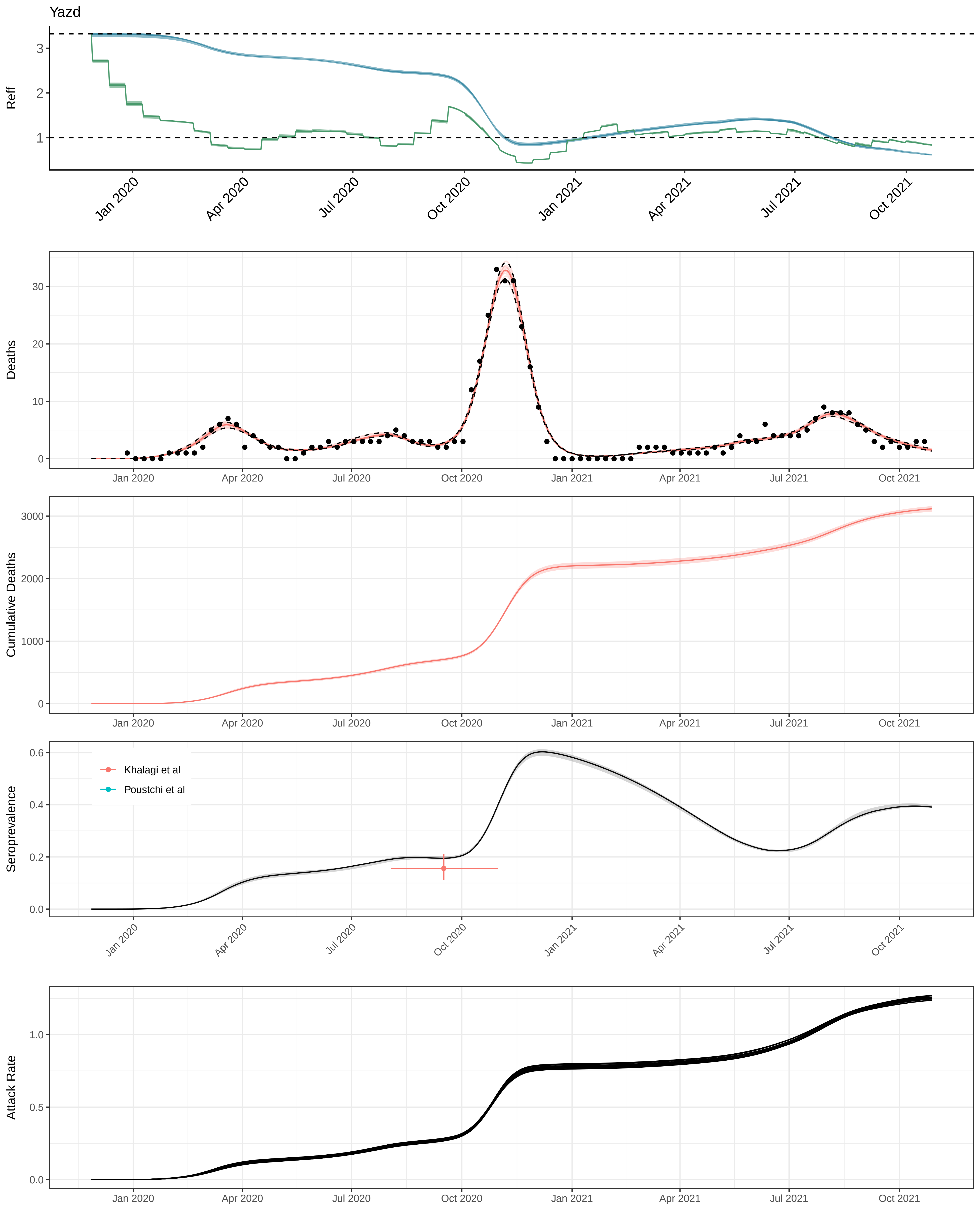

Yazd
Reff
Deaths
Cumulative Deaths
Seroprevalence
Khalagi et al
Poustchi et al
Attack Rate
Jan 2020
Apr 2020
Jul 2020
Oct 2020
Jan 2021
Apr 2021
Jul 2021
Oct 2021
